# *ShortKit-ML*: an open toolkit for measuring shortcut potential in medical imaging embeddings

**DOI:** 10.64898/2026.04.29.26352053

**Authors:** Sebastian Cajas, Aldo Marzullo, Sahil Kapadia, Filipe Santos, Felipe Ocampo Osorio, Qingpeng Kong, Alessandro Quarta, Po-Chih Kuo, Milit Patel, Raul Ignacio Rojas Sillery, Leo Anthony Celi

**Author notes:** Corresponding author *Email address:* (Sebastian Cajas). These authors contributed equally to this work (Sebastian Cajas, Aldo Marzullo, Sahil Kapadia).

## Abstract

**Background and Objective:** A classifier trained on frozen embeddings can take a shortcut through a sensitive attribute only if the embedding carries information about it beyond what the task label explains: its shortcut potential. We asked whether this can be measured on the embedding alone with a known false positive rate, and whether it predicts how strongly a classifier trained on confounded data relies on the attribute.

**Methods:** *ShortKit-ML* tests whether an embedding *E* and a sensitive attribute *A* are dependent given a task label *Y*. The primary test is a conditional probe: a logistic regression that predicts *A* from *E* and *Y*, scored by out-of-fold AUROC and calibrated by permuting *A* within levels of *Y*, between patients. Five secondary detectors share this null. We audited sex, age and race for seven frozen encoders on CheXpert and four on MIMIC-CXR, and trained heads on confounded CheXpert splits.

**Results:** Under the null the probe rejected in 0.02 of synthetic data sets. With *A* and *Y* strongly correlated, an unconditional probe rejected in 0.95 and the conditional probe in 0.03. No detector found signals that leave group means unchanged, unlike probes on squared or product features under the same null. The probe found attribute information in 21 of 21 CheXpert and 12 of 12 MIMIC-CXR cells. Across 42 CheXpert cells the probe score correlated with the true positive rate gap of a confounded head (Spearman 0.60, 95% CI 0.31 to 0.80 resampling encoders), 0.74 within sex and not significantly within age. Three embedding-level mitigations changed the gap by at most 0.01 relative to their controls. Refitting the head on a group-balanced subset closed it. A planted image marker was found in all 10 seeds.

**Conclusions:** One calibrated linear test measures shortcut potential on a frozen embedding, and on CheXpert its score correlated with how strongly a confounded logistic head relied on the attribute, clearly for sex. It does not show that a given head uses the attribute, and lowering the score did not remove reliance, so the head itself must be tested as well. Code is at https://github.com/criticaldata/ShortKit-ML.

## 1. Introduction

Deep learning models for medical imaging can reach expert-level accuracy on many diagnostic tasks, including skin cancer classification [1] and pneumonia detection on chest radiographs [2], as well as breast cancer screening [3]. High accuracy does not show that a model uses clinically meaningful evidence. Models often rely on shortcuts: signals that predict the label in the training data but are not part of the clinical mechanism, and that stop working when the data change [4]. In medical imaging, chest drains have served as proxies for pneumothorax [5], while pneumonia models have learned the hospital a radiograph came from [6] and hip fracture models have drawn on scanner and patient variables [7]. Patient race can be recognised from chest radiographs [8], and organ segmentation models show biases linked to sex, age and scanner manufacturer [9].

Discussions of shortcuts in pretrained representations often conflate encoding, use and harm. *Encoding* means that an embedding *E* carries information about a sensitive attribute *A*. A classifier trained on *E* for a task *Y uses* the attribute when it relies on that information, and the use becomes *harm* once it produces worse or unequal predictions, for example a gap in true positive rate between groups or a drop in accuracy under shift. Encoding is necessary for use and use is necessary for harm, but neither step follows from the one before. A model can encode sex and still not use it for a given diagnosis [10, 11], and a general-purpose encoder is expected to encode demographic attributes, because they correlate with much of what is visible in a radiograph [12, 13].

For a given task, what can be measured on the embedding alone is whether *E* carries information about *A* beyond what *Y* explains, the conditional dependence *I*(*E*; *A*| *Y*) *>* 0. We call this the *shortcut potential* of *E* for task *Y*. It is task-specific: the same embedding can have high potential for one label and none for another. When it is zero, a head trained on *E* has no attribute signal to exploit beyond *Y* itself. A positive value means only that a head could exploit it, and the only way to find out whether it does is to train the head and examine it [14, 13].

Can shortcut potential be measured on the embedding alone, with a test whose false positive rate is known? And when a head is trained on that embedding with confounded data, does the measured potential predict how strongly the head relies on the attribute? The second question matters because, as noted, general-purpose encoders are expected to encode demo-graphic attributes, so an audit that only reported their presence would flag almost every encoder. The audit is useful only if the size of its score tells the user something about the model that will be built on the embedding.

Methods that could estimate shortcut potential are spread across communities. Fairness toolkits such as Fairlearn [15] and AIF360 [16] report group disparities of trained classifiers. Probing studies train auxiliary classifiers on representations [17, 18]. Fay et al. [19] and Müller et al. [20] use dependence measures between features and a confounder as training penalties. These methods produce outputs on different scales with different thresholds, so their flags cannot be compared, and a detector that fires on pure noise looks the same in a table as one that has found a real signal.

This paper describes *ShortKit-ML*, a Python package that audits shortcut potential in frozen embeddings, and uses it to answer both questions. The contributions are:

1. A conditional probe for *I*(*E*; *A* |*Y*), calibrated by shuffling *A* within each level of *Y*, which is the single primary test (Section 3).
2. Five secondary detectors rewritten to score the attribute within task strata and calibrated by the same null, so that every reported *p*-value has a known false positive rate.
3. A synthetic benchmark in which *A* and *Y* are separate variables and the true answer is known, with linear, nonlinear and variance-shift signals (Section 4.1).
4. Audits of seven frozen encoders on CheXpert and four on MIMIC-CXR (Section 4.2), and a classification-head experiment on CheXpert that tests whether the measured potential predicts reliance on the attribute by a head trained on confounded data, and whether four mitigation methods reduce that reliance (Section 4.3).
5. An open package in which every table, figure and result number of this paper is regenerated by committed scripts from runs whose manifests record the code commit, the configuration and hashes of the inputs, with two integrity checks, a fake patient-level attribute and a row-alignment check between encoders, that caught faults in our own data (Appendix F).

The package also contains methods that need model training, gradients or concept examples. They are documented, but they are outside the audit score evaluated here (Section 3.5).

## 2. Related work

### Shortcut learning in medical imaging

Geirhos et al. defined shortcut learning as reliance on decision rules that work on the training distribution but fail to transfer [4]. Texture bias in ImageNet classifiers [21] and annotation artefacts in natural language inference [22] are early examples, and recent surveys give taxonomies of dataset bias [23] and of spurious correlations [24]. Medical examples include site-specific signal in pneumonia detection that degraded external performance [6] and scanner and hospital process variables in hip fracture prediction [7]. Other documented shortcuts are slide-origin signal in pathology [25], chest drains [26] and laterality markers [27] in radiography, and rulers in dermoscopy [28]. Reviews in radiology catalogue these routes [29, 30]. Hill et al. trained models on knee radiographs to predict targets that should not be visible, such as whether a patient drinks beer, and reached above-chance AUROC through sex, site and scanner signals that preprocessing did not remove [12].

### Encoding of protected attributes

Gichoya et al. showed that deep models recognise self-reported race from chest radiographs with high accuracy, and that the signal survives image degradation [8]. Glocker et al. found that chest X-ray disease detection models encode sex and race but appear to make little use of them for disease prediction [11], and that classifiers built on the frozen features of a chest radiography foundation model show subgroup performance gaps [31]. Jones et al. showed that subgroup separability, how well the group can be recovered from the image, governs how much a group’s performance degrades when its labels are biased [32], and argued that the right mitigation depends on the causal structure of the bias [33]. Yang et al. reported that models which encode more demographic information show larger fairness gaps, and that removing the information does not always help on external data [34].

Brown et al. addressed the step between encoding and use directly: shortcut testing varies how strongly a model encodes an attribute and measures the effect on fairness, which separates attributes that are used from attributes that are only present [10]. Petersen et al. argued that a demographically invariant representation is neither necessary nor sufficient for a fair model, and that a representation carrying only disease-relevant features will correlate with group whenever those features do [13]. Kina and Petersen trained pneumothorax classifiers with chest drains as a confounded shortcut, on fine-tuned and frozen foundation-model encoders, and found that recalibrating the head separately for each shortcut group, with the representation unchanged, recovered much of the performance on images where drain and label disagree [14]. On frozen encoders, then, much of shortcut use is decided in the head. Our audit sits at the encoding step, and the head experiment tests whether the encoded signal gets used.

### Probing and dependence measures

Probes were introduced to read what a layer represents [17] and are now a standard tool, with known caveats about probe capacity and what a high probe score means [18]. Bolukbasi et al. removed a gender direction from word embeddings [35]. Gonen and Gold-berg showed that the attribute stays recoverable after such removal [36], and Ravfogel et al. removed it iteratively by nullspace projection [37]. Image embeddings show measurable social bias [38]. Fay et al. split the encoder output into a task part and a confounder part and minimised the mutual information between them during training, so that the task prediction becomes independent of the confounder [19]. Müller et al. benchmarked such dependence penalties (mutual information estimated with MINE, and distance correlation) against adversarial training and rebalancing, on Morpho-MNIST and on CheXpert with sex as the shortcut for pleural effusion. They measured encoding with a *k*NN probe on each latent subspace and use through the drop on a test set with the correlation inverted [20]. Both use the dependence measure as a training penalty and need the confounder label and access to the encoder, and with a frozen encoder there is no training loop to apply them in [14]. We use a conditional dependence estimate as an audit statistic on a frozen encoder, and calibrate it with a conditional permutation test [39]. Two-sample tests such as MMD [40] and permutation-based fairness tests [41] are close relatives.

### Slice discovery

GEORGE [42] and Domino [43] cluster embeddings to find subpopulations on which a model errs, without attribute labels, and related work uses hierarchical clustering for the same purpose [44]. Our HBAC detector borrows the clustering step but scores agreement between clusters and a known attribute, so it needs attribute labels.

### Fairness toolkits and benchmarks

Fairlearn [15], AIF360 [16] and Aequitas [45] implement group fairness metrics and mitigation for trained classifiers, mostly on tabular data. MEDFAIR benchmarks bias mitigation algorithms across medical imaging datasets with a common training and model selection protocol, and found that few methods beat empirical risk minimisation once model selection is controlled [46]. FairMedFM extends this to medical foundation models and evaluates the fairness of zero-shot, linear-probe and fine-tuned use across many datasets [47]. Both evaluate the fairness of trained models, and neither tests the representation for attribute information conditional on the task. Table 1 compares these tools with *ShortKit-ML*.

**Table 1:** Comparison of existing bias and shortcut detection tools. Footnotes define the entries in the Embedding analysis and Multi-method columns.

| Tool | Level | Embedding analysis | Modality | Multi-method | Auto report | Medical focus |
|---|---|---|---|---|---|---|
| Fairlearn [15] | Prediction | No | Tabular | No | No | No |
| AIF360 [16] | Prediction | No | Tabular | Partial<sup>*a*</sup> | No | No |
| Aequitas [45] | Prediction | No | Tabular | Partial<sup>*a*</sup> | Yes | No |
| What-If Tool [48] | Prediction | No | Tabular | Yes | Yes | No |
| WEAT [49] | Embedding | Word only<sup>*b*</sup> | Text | No | No | No |
| Domino [43] | Embedding | Slice discovery<sup>*c*</sup> | Vision/Text | No | No | No |
| MEDFAIR [46] | Prediction | No | Vision (med.) | Partial<sup>*a*</sup> | No | Yes |
| FairMedFM [47] | Prediction | No | Vision (med. FMs) | Partial<sup>*a*</sup> | No | Yes |
| ***ShortKit-ML*** **(Ours)** | **Embedding** | **Yes (6 calibrated)**<sup>*d*</sup> | **Image embeddings**<sup>*e*</sup> | **1 primary, 5 secondary** | **Yes** | **Yes** |

<sup>*a*</sup> Partial: several fairness metrics or mitigation algorithms are offered, but they are computed or run separately and the toolkit does not combine their outputs into one assessment. <sup>*b*</sup> Word only: WEAT measures association between word embeddings and attribute word sets. It does not apply to image embeddings. <sup>*c*</sup> Slice discovery: Domino clusters embeddings to find subgroups with high model error, and it does not test a named sensitive attribute. <sup>*d*</sup> Six attribute-aware detectors calibrated by the same conditional permutation null. The package implements 20 methods in total (Section 3.5). <sup>*e*</sup> Validated on chest radiograph and face image embeddings. Any fixed-length vector can be passed, but other modalities are untested.

## 3. Methods

### 3.1. Problem formulation

Let *E ∈* ℝ^*n×d*^ be embeddings of *n* images from a frozen encoder, where *Y* denotes the task label and *A* is a sensitive attribute with *G* groups. The audit tests the null hypothesis

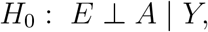

which holds exactly when the conditional mutual information *I*(*E*; *A* |*Y*) is zero. In words, the audit asks whether the embedding holds any information about the attribute that the task label does not already account for. If *H*_0_ holds, a classifier trained on *E* to predict *Y* cannot gain anything from *A* beyond its correlation with *Y*, so the shortcut potential is zero. Rejecting *H*_0_ shows only that the potential is positive. Whether a particular head uses it is tested separately in Section 4.3. Conditioning on *Y* matters: if *A* and *Y* are correlated, an embedding that encodes only *Y* will also predict *A*, and an unconditional test would flag it. The tests in this paper are linear, so they have power against dependence that shifts the mean of *E* between groups of *A* within *Y*. Section 5.1 shows that they have none against signals that leave those means unchanged.

A positive result reads differently when the attribute has physical correlates in the image. Sex, for example, is associated with chest anatomy, so a positive audit for sex is expected and may be clinically harmless. The audit reports only that the information is there, since an anatomical correlate and an unwanted one are both dependence between *E* and *A* given *Y*, and it leaves that judgement, and the head experiment, to the analyst.

### 3.2. Calibration by conditional permutation

Each detector *m* returns a continuous statistic *s*_*m*_(*E, A, Y*), where larger means more information about *A* in *E*. Calibration asks how large that statistic would be if the embedding carried no information about the attribute beyond the task label, and answers this by recomputing it on data where that is true by construction. To build such data, we permute *A* within each stratum of *Y*, which keeps the joint distribution of (*A, Y*) and of (*E, Y*) but breaks any dependence between *E* and *A* given *Y* [39].

When a cohort has several images per patient, the attribute is constant within a patient, so we permute it between patients within three strata (all of a patient’s images negative for the task, all positive, or mixed) and keep all images of a patient together in the cross-validation folds. On CheXpert with Pneumothorax as the task these strata hold 421, 12 and 44 of the 477 patients, so in the stratified permutation no patient is without a partner; in the labelled-only subsets of Section 4.3 and in the test halves audited after mitigation the smallest stratum held 4 patients. Patients in one stratum are treated as exchangeable, which assumes that their number of images and, in the mixed stratum, their share of positive images are unrelated to *A*. On CheXpert neither quantity differed by sex or by age (Mann–Whitney *p* from 0.55 to 0.83 over the 4 comparisons, 44 mixed patients). An attribute that can differ between images of one patient, such as the planted marker, is permuted in blocks: whole patients’ attribute vectors are exchanged between patients with the same task labels and number of images. In the block permutation of the planted-marker audit, 449 of the 477 patients had at least one such partner. The 28 without one keep their observed vector in every permutation, which leaves the test valid and only reduces the number of distinct permutations.

Repeating this *K* times gives null statistics 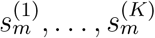 and the permutation *p*-value

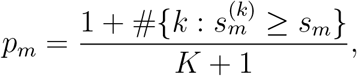

which is valid at any sample size [50]. Under *H*_0_, and when *A* is exchangeable within the strata, every detector then has false positive rate at most *α*, whatever its scale or dimensionality. The observed rates on synthetic null data are given in Table 2 and Section 5.1. We use *K* = 500 for the probe in the clinical audits and the head experiment, and *K* = 200 for the secondary statistics, the synthetic grid and the audits after mitigation (Appendix D). Within one audit cell (one embedding, attribute and task), the conditional probe is the single primary test and is judged at its own level, *p <* 0.05. The *p*-values of the five secondary detectors are adjusted by the Benjamini– Hochberg procedure [51] across those five, and a secondary detector is flagged when its adjusted value *q*_*m*_ *<* 0.05. Earlier versions of the package compared each statistic against a fixed threshold, such as probe accuracy above 70%. Those thresholds are no longer used for any decision.

**Table 2:** Detectors and their status in the revised audit. For every statistic *s*_*m*_, larger means more information about *A* in *E* given *Y*. Every scored detector takes only the embedding *E*, the attribute *A* and the task label *Y*, and is calibrated by the conditional permutation null of Section 3.2 (*A* shuffled within *Y* strata, *K* permutations; the probe judged at *p <* 0.05, Benjamini–Hochberg across the five secondary detectors of a cell). Null FPR: fraction of synthetic data sets with *δ* = 0 rejected at *p <* 0.05 before correction, pooled over all settings and 10 seeds. After BH across the five secondary detectors no secondary detector exceeded 0.03 (Table E.1).

| **Detector** | **Status** | **Statistic *s*<sub>*m*</sub>** | **Calibration** | **Null FPR** |
|---|---|---|---|---|
| Conditional probe | Primary | Out-of-fold AUROC of L2 logistic regression predicting *A* from [*E*, *Y*], 5-fold CV stratified on (*A*, *Y*) | Perm., *K* = 500 (synthetic: 200) | 0.02 |
| Statistical | Secondary | Fraction of dimensions with a BH-significant *A* difference within *Y* strata (Mann–Whitney or Kruskal–Wallis) | Perm., *K* = 200 | 0.07 |
| Geometric | Secondary | Between-group centroid distance over pooled within-group spread, averaged over *Y* strata | Perm., *K* = 200 | 0.07 |
| Bias direction PCA | Secondary | Standardised projection gap of group means on the first PC of group-mean differences, within *Y* | Perm., *K* = 200 | 0.08 |
| HBAC | Secondary | Normalised mutual information between *k*-means clusters of *E* and *A*, within *Y* | Perm., *K* = 200 | 0.04 |
| Frequency | Secondary | Conditional probe AUROC using only the top-ranked dimensions (ranked on training folds) | Perm., *K* = 200 | 0.07 |
| SIS | Removed from score | n/a: the implementation fits on *Y* with a fixed 15% subset and has no attribute-aware form yet | None | n/a |
| Equalized odds (TPR and FPR gaps) | Head experiment only | Gaps of a held-out head trained on *E* (Section 4.3), which are properties of a classifier; demographic parity and intersectional gaps are implemented but not reported | n/a | n/a |
| GroupDRO, GCE, SSA | Outside score | Need training dynamics or labelled and unlabelled splits that a frozen-embedding audit does not have | n/a | n/a |

### 3.3. Primary test: the conditional probe

The primary test is a conditional probe. The idea is that if a linear classifier predicts the attribute from the embedding and the task label better than it could from the task label alone, the embedding carries attribute information that the task label does not explain. An L2-regularised logistic regression predicts *A* from the standardised embedding concatenated with *Y*. Its score is the out-of-fold AUROC from 5-fold cross-validation stratified on (*A, Y*) and grouped by patient where patient identifiers exist. When *G >* 2 it is macro-averaged over one binary one-vs-rest regression per group, and in every case it is averaged over five random fold splits. Standardisation is fitted inside each training fold, and *Y* enters as one more standardised column under the same penalty (every task here is binary). Each permuted data set uses a single fold split, which makes the test slightly conservative (the permuted score has the same expectation as the observed five-split average but more variance, so the null is wider), and the fold splits come from a random stream separate from the permutations, so the observed score does not depend on *K*. Across fold splits the score had an SD of at most 0.018 on CheXpert and 0.013 on MIMIC-CXR. The regularisation strength is *C* = 1.

Because *Y* is an input, the probe can reach an AUROC above 0.5 from *Y* alone when *A* and *Y* are correlated. The permuted data keep that correlation, so the null distribution absorbs it and only the information that *E* adds is tested. For the same reason the AUROC is pooled over the *Y* strata rather than computed within each: pooling raises the null mean when *A* and *Y* are correlated, and under *H*_0_ it raises the observed score by the same amount. Along with the probe AUROC and its permutation *p*-value, we report a 95% confidence interval from 2000 bootstrap resamples of patients (identities on CelebA) applied to the out-of-fold predictions, which does not include the variation between fold splits reported above. We chose a linear probe so that a positive result means the attribute is linearly available to a linear head, which is how frozen embeddings are usually used.

### 3.4. Secondary detectors

Five secondary detectors are reported with their continuous scores and calibrated *p*-values. Each was rewritten so that it reads *A* and works within strata of *Y* (Table 2, formulas in Appendix A). Within each *Y* stratum, the statistical detector tests every dimension for a group difference with Mann– Whitney or Kruskal–Wallis tests, and its score is the fraction of dimensions that stay significant after Benjamini–Hochberg correction. The geometric score is the distance between group centroids relative to the pooled within-group spread, averaged over *Y* strata. For bias direction PCA [35], samples are projected on the first principal direction of the group-mean differences within each stratum, and the score is the standardised gap of the projections. HBAC [44, 52] runs *k*-means (*k* = 8) on *E* within each stratum and scores the normalised mutual information between clusters and *A*. The frequency detector keeps only the top 5% of dimensions, ranked by their group difference on the training folds, and reports the conditional probe AUROC on those [53].

For descriptive purposes we also count how many detectors are flagged in a cell,

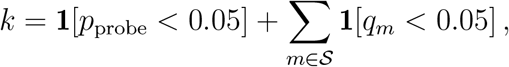

where *p*_probe_ is the permutation *p*-value of the conditional probe, *S* is the set of the five secondary detectors (|*S*|= 5), *q*_*m*_ is the Benjamini–Hochberg adjusted *p*-value of secondary detector *m* computed across those five, and 1[*·*] is the indicator function, equal to 1 when its argument is true and 0 otherwise. Decisions rest on the conditional probe alone, judged at the conventional level *p <* 0.05, and the count is reported as a description. The secondary detectors are adjusted among themselves so that their flags share one false discovery level without touching the primary decision. With *K* = 500 the smallest attainable *p*-value is 1/(*K* + 1) (and 1/(200 + 1) for the secondary statistics), and tables print the unadjusted *p*. Section 5.2 reports how often the probe was significant and the one cell on which adjusting it with the secondary detectors would change the decision. Because the detectors measure related properties of the same embedding, their flags are correlated, and Section 5.1 reports that correlation and the operating characteristics of every threshold *k* = 1, …, 6.

### 3.5. Package scope: what runs on embeddings alone

The package implements 20 methods. Six of them, the six calibrated detectors in Table 2, need only (*E, A, Y*) and are therefore usable when the encoder is a black box or an API that returns embeddings. The other 14 need something an embedding audit does not have: a trained classifier, training dynamics, concept examples, images and model gradients, or an auxiliary model. We kept them because they are useful when the user has the full model and training data, and because several are standard baselines. Appendix B lists each with what it needs, and none contributes to any result in this paper except the group gaps computed for the heads of Section 4.3.

### 3.6. Mitigation

The package includes six mitigation methods (Appendix C). Four of them work on frozen embeddings and are evaluated here with the package defaults. Adversarial and contrastive debiasing pass the embeddings through a small learned debiasing network, which can change the audit score by itself, so each is compared with a control: the same network trained the same way with the adversarial or contrastive term switched off (for the contrastive method the task loss then has weight 1 instead of 0.25). Shortcut masking sets to zero the 5% of embedding dimensions with the largest weight on the attribute direction, the difference between the group means of *A* after centring within *Y*, estimated on the training split. Its control sets to zero the same number of randomly chosen dimensions. Last-layer retraining (DFR) [54] leaves the embedding unchanged and refits the head on a training subset balanced over the four (*A, Y*) groups, so its audit score cannot change and only the head is compared. Each method is run with 3 seeds. For each method and control, the head is retrained on the transformed training embeddings and the audit is rerun on the transformed test-half embeddings, and both the audit score and the task AUROC are reported, so that removal of attribute information can be weighed against loss of task information.

### 3.7. Implementation

Each detector implements detect(*E, Y, A*) *→ s*_*m*_, and the calibration layer wraps any such function, so a new statistic inherits the permutation null without further work. The audit is embedding-agnostic in the sense that any fixed-length vector with task and attribute labels can be passed to it, but only chest radiograph and face image embeddings were tested here. Figure 1 shows the pipeline. *ShortKit-ML* is a Python package installed with pip from its repository. One command per dataset reruns the audit and the head experiment and then the table and figure scripts. Every run writes a manifest that records the code commit and configuration together with hashes of the data (Appendix D). The probe audit of 50,000 images with *K* = 500 permutations took 11 min on a laptop CPU.

**Figure 1:**
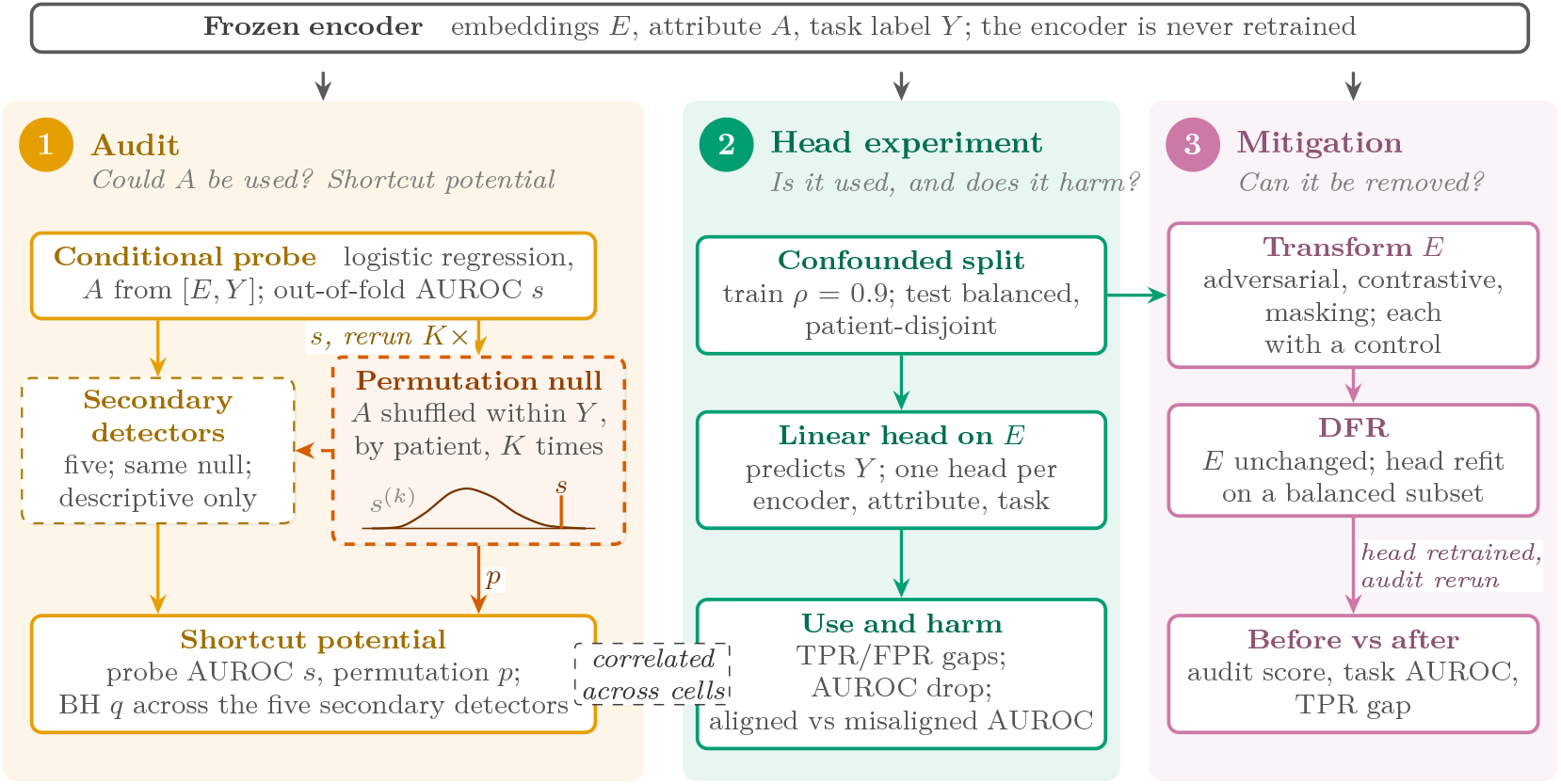
The *ShortKit-ML* pipeline in this paper. Left: the audit, which needs only embeddings, the task label and the attribute, and estimates shortcut potential. The permutation null that calibrates it is shown in the inset. Middle: the classification-head experiment, which trains a head on confounded data to measure whether the attribute is used and with what effect, and compares this with the audit. Right: the mitigation methods, applied to the confounded training embeddings and compared before and after. The audit addresses the first question of the Introduction, and the head experiment takes up the second.

## 4. Experiments

The synthetic benchmark addresses the first question of the Introduction on data where the truth is known: it checks that the probe and the secondary detectors hold their false positive rate, and it measures their power. The clinical audits then apply the test to common frozen encoders on two chest radiograph cohorts, for sex, age and race. The classification-head experiment addresses the second question: it trains heads on confounded CheX-pert data and asks whether the audit score predicts how much they rely on the attribute, and whether four mitigation methods reduce that reliance. A planted image marker supplies a shortcut with known ground truth, and CelebA is a non-medical check. Table 3 lists which method is used in which experiment. Seeds and run manifests are described in Appendix D.

**Table 3:** Which method is used in which experiment. •: scored and calibrated. *°*: computed on a trained head, not part of the audit. –: not run. The package implements 20 methods, and the 6 audit detectors are the only ones that need nothing beyond (*E, A, Y*).

| **Method** | **Synthetic** | **CheXpert** | **MIMIC-CXR** | **CelebA** | **Head exp.** |
|---|---|---|---|---|---|
| Conditional probe (primary) | ● | ● | ● | ● | ● |
| Statistical | ● | ● | ● | ● | – |
| Geometric | ● | ● | ● | ● | – |
| Bias direction PCA | ● | ● | ● | ● | – |
| HBAC | ● | ● | ● | ● | – |
| Frequency | ● | ● | ● | ● | – |
| Eq. odds (TPR, FPR gaps) | – | – | – | – | ○ |
| Four mitigation methods | – | – | – | – | ○ |
| Other 13 package methods | – | – | – | – | – |

### 4.1. Synthetic benchmark

The synthetic data make the answer known. The task label is *Y* ~ Bernoulli(0.5) and the attribute satisfies *P* (*A* = *Y*) = *ρ* for *ρ* equal to 0.5, 0.7 or 0.9, so *A* and *Y* are distinct variables whose correlation we control. Embeddings have *d* standard normal dimensions. The task signal sits in its own block of dimensions. The attribute signal sits in a separate block and comes in three forms. In the linear mean shift the two groups differ by exactly *δ* standard deviations (each shifted by *±δ*/2, so *δ* is Cohen’s *d* in each attribute dimension). The nonlinear XOR form leaves both group means at zero and carries the attribute in the sign of the product of two dimensions. Under the variance shift the groups have equal means and unequal variances (Appendix E). We varied *δ ∈ {*0, 0.2, 0.5, 0.8, 1.2, 2.0 *}, n ∈ {*200, 1000*}* and *d ∈ {*128, 512*}*, with 10 seeds per configuration. The ground truth is that *I*(*E*; *A* |*Y*) *>* 0 exactly when *δ >* 0. At *δ* = 0 with *ρ* = 0.9, the embedding predicts *A* through *Y* but carries no extra information, which is the case a conditional test has to get right.

A mean shift in a few dimensions is a simplification. Shortcut signal in real encoders is likely spread across many dimensions and may be nonlinear. The XOR and variance variants were added to test whether each detector can see signals that leave group means unchanged, and the planted image marker (Section 4.3) tests the pipeline on a shortcut that passes through a real encoder. For each detector and variant we report sensitivity (flag rate at *δ >* 0) and specificity (one minus flag rate at *δ* = 0), the operating characteristics of the descriptive count *k* for *k* = 1, …, 6, the pairwise correlation of detector flags, and the effective number of independent detectors computed from the eigenvalues of that correlation matrix [55, 56].

### 4.2. Clinical cohorts

#### CheXpert

We used the first 2,000 images streamed from a public Hugging Face mirror of the CheXpert training set [57]. These follow the order of the official `train.csv`, which is not random. They come from 477 patients, and 423 are lateral views. The mirror itself has no patient identifiers, so we joined each image by its path to the official `train.csv`, which gives the patient identifier, sex, age, view and findings, and took race per patient from the CheXpert demographics file. Images without a demographics record, or with race recorded as unknown or refused (217 images), are left out of the race analyses. Native Hawaiian or Pacific Islander (24 images) is its own group and counts as non-White in the binary split. All cross-validation and permutation on CheXpert is grouped by patient. The audit task is Pneumothorax, with *Y* = 1 for a positive label and *Y* = 0 otherwise (prevalence 10.9%). *Y* = 0 covers 584 negative, 23 uncertain and 1,175 blank labels, and Table F.7 repeats the audit on the 802 images with a definite label.

#### MIMIC-CXR

We used 1,491 subjects from MIMIC-CXR [58], one image per subject, so ordinary cross-validation is already patient-level. The images are a coreset of 1,999 images drawn for an earlier embedding project by stratified sampling on pathology labels (strategy “PathologyStratified-Clean”, seed 0, of the public embedding dataset https://huggingface.co/datasets/MITCriticalData/qml-mimic-cxr-embeddings, which also provides the coreset strata used in [59]). All are frontal (AP or PA) views. Rows without a linked hospital admission outcome were removed. The task is in-hospital mortality of the linked admission (hospital_expire_flag in MIMIC-IV [60]), prevalence 6.0%. Mortality is an outcome that depends on many factors outside the image, and no radiological finding defines it.

#### Encoders

All encoders were used frozen, with their published weights and no fine-tuning on either cohort. Table 4 separates architecture from pretraining. Some public encoders were pretrained on corpora that include CheXpert or MIMIC-CXR images. RAD-DINO [61] publishes its list of training images, and all 2000 CheXpert images in our sample (477 patients) and 1479 of the 1491 MIMIC-CXR images are on it. RAD-DINO is self-supervised, so it saw these images without their labels or attributes, and its scores here describe an encoder that has seen the audited images. We did not train any encoder, so no patient in our analysis was used in a training step we controlled. All seven CheXpert embeddings are extracted with code in the repository from pinned public checkpoints. In the first submission four of them (ViT-B/32, DINOv2, RAD-DINO, MedSigLIP) were arrays stored without their extraction code, which could not be reproduced exactly. We re-extracted those four from the same images, and every CheXpert result here uses the re-extracted arrays. The extraction code reproduces the stored ViT-B/16 array, whose code was kept, to cosine 1.000, and Table F.6 compares the old and new arrays for the other four. The MIMIC-CXR embeddings come from the same public dataset: the CLS token of ViT-Base-Patch16-224 and ViT-Base-Patch32-224, RAD-DINO, and MedSigLIP-448. The dataset’s documentation names google/siglip-so400m-patch14-384 for the last one, but re-embedding 50 of its images with google/medsiglip-448 reproduced the stored vectors (cosine 1.000 for every image), while the general-purpose SigLIP So400m checkpoint did not (median 0.15).

**Table 4:** Frozen encoders: architecture, pretraining and embedding dimension. CheXpert uses all seven, and MIMIC-CXR uses RAD-DINO, ViT-B/16, ViT-B/32 and MedSigLIP. ^*†*^CheXpert array re-extracted for this revision from the named checkpoint. The arrays used in the first submission had no extraction code (Table F.6).

| **Name** | **Architecture** | **Pretraining** | **Dim** |
|---|---|---|---|
| ResNet-50 | CNN | Supervised, ImageNet-1k (torchvision default weights) | 2048 |
| DenseNet-121 | CNN | Supervised, ImageNet-1k (torchvision default weights) | 1024 |
| ViT-B/16 | ViT-Base, patch 16 | Supervised, ImageNet-1k (torchvision default weights) | 768 |
| ViT-B/32<sup>†</sup> | ViT-Base, patch 32 | Supervised, ImageNet-1k (torchvision default weights) | 768 |
| DINOv2<sup>†</sup> | ViT-Base, patch 14 | Self-supervised, natural images (facebook/dinov2-base) | 768 |
| RAD-DINO<sup>†</sup> | ViT-Base, patch 14 | Self-supervised (DINOv2 recipe), chest radiographs (microsoft/rad-dino) | 768 |
| MedSigLIP<sup>†</sup> | SigLIP vision tower, 448 px | Image–text contrastive, medical images (google/medsiglip-448) | 1152 |

#### Attributes

Sex is binary as recorded. Age is binary, 60 or over against under 60. A four-bin version (under 40, 40 to 59, 60 to 79, 80 and over) is a sensitivity analysis. Race is White against non-White in the primary analysis, because several groups are small in these samples. A sensitivity analysis tests separately the race groups with at least 50 images (White, Black and Asian on CheXpert, and White and Black on MIMIC-CXR), and repeats this keeping only group and *Y* combinations with at least 50 images. On CheXpert only White patients reach 50 images with *Y* = 1, so that version compares groups among *Y* = 0 images only. Group sizes for every cohort and attribute are given in Table F.4.

### 4.3. Classification-head experiment

The audit measures potential, and to measure use and harm we trained classification heads on frozen CheXpert embeddings, for every encoder, with *A* equal to sex and then age, and three tasks: Pneumothorax, Pleural Effusion and Cardiomegaly. For these tasks, uncertain and blank labels are dropped per task. Train and test sets are disjoint at patient level (a two-fold split grouped by patient and stratified on (*A, Y*)), and each cell is repeated with 10 seeds.

#### Confounded training

An L2 logistic head on standardised embeddings is trained on a subsample with *P* (*Y* = 1 | *A* = 1) = 0.9 and *P* (*Y* = 1 | *A* = 0) = 0.1 (*ρ*_train_ = 0.9), and tested on a patient-disjoint subsample with all four (*A, Y*) groups of equal size (*ρ* = 0.5). Patients are first split in half, stratified on the image-level (*A, Y*) labels with patients as groups (scikit-learn’s StratifiedGroupKFold), so every image of a patient falls in one half, with a different split for each seed. The subsamples below draw images within each (*A, Y*) cell of a half. In the training half the (*A, Y*) groups (1, 1) and (0, 0) each hold 0.45*N* images and (1, 0) and (0, 1) each hold 0.05*N*, with *N* the largest size that the smallest group allows, so *N* changes with the split. Training sets held 42 to 272 images (24 to 124 patients), and test sets 60 to 404 images (40 to 153 patients).

Reliance on *A* is measured two ways: the gaps in true and false positive rate between groups on the test set, and the drop in test AUROC relative to a head trained on an unconfounded subsample of the same size. We also report the mean absolute change in the head’s output when the attribute direction is removed from the test embeddings (output shift). Appendix C explains why it is not used as a measure of reliance. That direction is the difference of the two group means after centring each *Y* stratum, estimated on the training pool. Each test embedding *x* is replaced by *x −* ((*− x m*)^*⊤*^*u*) *u*, with *u* the unit direction and *m* the training mean, and the confounded head is applied again. We also report the head’s AUROC separately on test images where *A* and *Y* agree with the training correlation (aligned) and where they disagree (misaligned), as in Kina and Petersen [14]. To check that the result does not depend on the linear head, the reliance measures are repeated on the same splits and seeds with a multilayer perceptron at the scikit-learn defaults (one hidden layer of 256 ReLU units, Adam with learning rate 10^*−*3^, L2 weight 10^*−*3^ and up to 500 iterations).

#### Audit score against reliance

Across encoder, attribute and task cells we compute the Spearman rank correlation between the conditional probe score, computed on all labelled images of the task, and each reliance measure, with a bootstrap confidence interval over cells. The cells are not independent, since the seven encoders embed the same images and the attributes and tasks reuse the same patients. We therefore also report an interval from resampling whole encoders and the partial correlation given attribute and task. With seven encoders this interval is itself imprecise.

#### Mitigation

The three embedding-level mitigations of Section 3.6 and their controls are fitted on the confounded training embeddings and applied to the training and test embeddings, after which the head is retrained and the audit rerun on the test half of the patients. For DFR only the head is refit. Because the test half is smaller, the probe score before mitigation is lower than in Table 8, which uses all labelled images. The TPR gap before mitigation is the mean over the first 3 of the 10 seeds. Before and after each method we report the audit score and the task AUROC, together with the TPR gap.

#### Planted marker

To test the pipeline on a shortcut with known ground truth that passes through a real encoder, we stamped a white square (27 by 27 pixels, intensity 255, at the same position on every image, its corner at 30% of the width and height) into the CheXpert images at 320 pixels on the shorter side, before the encoder’s resizing to 224, embedded the images with the torchvision ResNet-50, and audited with the marker as the attribute and Pneumothorax as the task. For the audit, images with *Y* = 1 received the marker at rate *r* = 0.5. For the head, the training images with *Y* = 1 received it at rate 0.9. Images with *Y* = 0 never carried the marker in training. Patients were split in half for the head, and test images received the marker on a random half regardless of *Y*, so a head that ignores the marker shows no gap. The experiment was repeated with 10 seeds.

### 4.4. CelebA

CelebA [62] was used as a non-medical check. We extracted ResNet-50 embeddings (2,048 dimensions) from 10,000 images drawn at random with a fixed seed from the 202,599 and audited *A* = Male given each of three tasks, *Y ∈ {*Blond hair, Heavy makeup, Attractive*}*, which are known to correlate with Male in this dataset. Images are grouped by celebrity identity for cross-validation and permutation.

## 5. Results

### 5.1. Synthetic benchmark

The conditional permutation null held its nominal level. Pooled over all null configurations, the detectors rejected in 0.02 (conditional probe) to 0.08 (bias direction) of runs at *p <* 0.05, the level at which the probe is judged, and after Benjamini–Hochberg across the five secondary detectors no secondary detector exceeded 0.03 (Table E.1). The grid has 40 null data sets for each *ρ*, and at *ρ* = 0.9 the geometric and statistical detectors rejected in 0.17 and 0.12 of them, and the bias-direction detector in 0.15. A permutation test is exact under *H*_0_ whatever the statistic, so we drew 500 further null data sets at *ρ* = 0.9, *n* = 1000 and *d* = 128, the grid setting with the most rejections. There the three rates were 0.05, 0.03 and 0.06 (HBAC 0.07, frequency 0.04, probe 0.03), so the higher grid values reflect how few grid data sets there are. Conditioning mattered most when *A* and *Y* were strongly correlated. At *ρ* = 0.9 and *δ* = 0, where the embedding predicts *A* only through *Y*, an unconditional probe rejected the null in 0.95 of data sets and the conditional probe in 0.03.

For the linear signal, the probe’s sensitivity was 0.16 at *δ* = 0.2, 0.62 at *δ* = 0.5 and 0.87 at *δ* = 0.8 (Table 5, Figure 2). The frequency, statistical and geometric detectors had similar power, and HBAC had much less.

**Table 5:** Synthetic benchmark: specificity at *δ* = 0 and sensitivity at each *δ >* 0 (Cohen’s *d*), per detector and signal variant. Pooled over *n ∈{* 200, 1000*}, d ∈{*128, 512*}, ρ ∈{*0.5, 0.7, 0.9, 10*}* seeds per configuration. Flag: probe at *p <* 0.05, secondary detectors at Benjamini–Hochberg *q <* 0.05 across the five.

| Variant | Detector | Spec. (δ = 0) | δ = 0.2 | δ = 0.5 | δ = 0.8 | δ = 1.2 | δ = 2 |
|---|---|---|---|---|---|---|---|
| linear | Probe | 0.98 | 0.16 | 0.62 | 0.87 | 0.99 | 1.00 |
|  | Stat. | 0.99 | 0.07 | 0.66 | 0.91 | 0.99 | 1.00 |
|  | Geom. | 0.99 | 0.14 | 0.57 | 0.82 | 0.96 | 0.99 |
|  | BiasDir | 0.97 | 0.12 | 0.48 | 0.68 | 0.80 | 0.97 |
|  | HBAC-NMI | 1.00 | 0.03 | 0.15 | 0.30 | 0.45 | 0.64 |
|  | Freq. | 0.97 | 0.12 | 0.70 | 0.93 | 0.99 | 1.00 |
| xor | Probe | 0.98 | 0.02 | 0.02 | 0.02 | 0.02 | 0.02 |
|  | Stat. | 0.99 | 0.01 | 0.01 | 0.01 | 0.01 | 0.03 |
|  | Geom. | 0.99 | 0.01 | 0.01 | 0.02 | 0.02 | 0.03 |
|  | BiasDir | 0.97 | 0.03 | 0.03 | 0.03 | 0.03 | 0.04 |
|  | HBAC-NMI | 1.00 | 0.01 | 0.00 | 0.00 | 0.00 | 0.02 |
|  | Freq. | 0.97 | 0.03 | 0.03 | 0.03 | 0.03 | 0.03 |
| variance | Probe | 0.98 | 0.02 | 0.02 | 0.02 | 0.02 | 0.02 |
|  | Stat. | 0.99 | 0.01 | 0.01 | 0.02 | 0.02 | 0.02 |
|  | Geom. | 0.99 | 0.01 | 0.01 | 0.03 | 0.03 | 0.04 |
|  | BiasDir | 0.97 | 0.03 | 0.03 | 0.04 | 0.06 | 0.09 |
|  | HBAC-NMI | 1.00 | 0.02 | 0.01 | 0.00 | 0.02 | 0.02 |
|  | Freq. | 0.97 | 0.03 | 0.03 | 0.03 | 0.03 | 0.03 |

**Figure 2:**
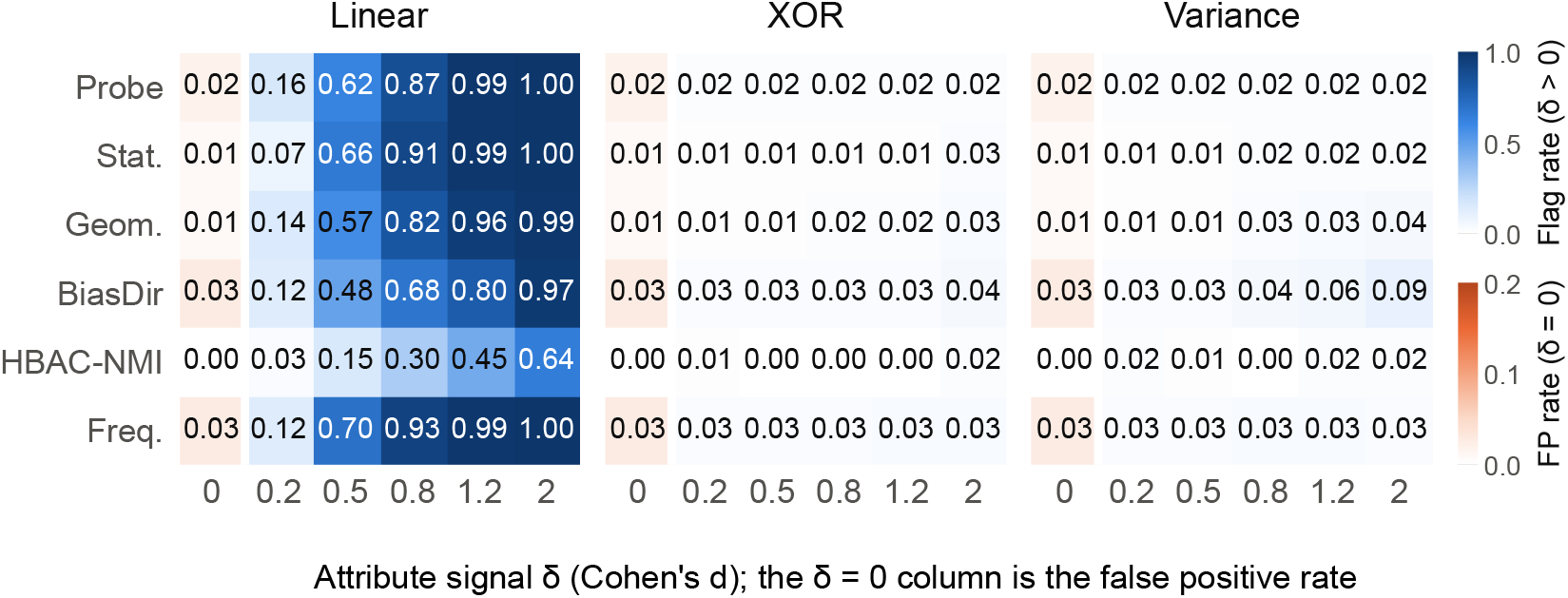
Flag rate per detector and effect size for the three signal variants, pooled over *n, d* and *ρ*, 10 seeds. The *δ* = 0 column uses a separate colour scale: any colour there is a false positive.

Every detector missed the nonlinear and variance-shift signals. At *δ* = 2.0 on the XOR variant, sensitivity was 0.02 for the probe, 0.02 for HBAC and 0.03 for the statistical detector, and on the variance-shift variant the same three were 0.02, 0.02 and 0.02. For every detector and every *δ*, sensitivity on these two variants stayed at or near the level of its false positive rate under the null. All six statistics respond to differences in group means, directly or through a linear model, and neither variant changes the means.

Over the whole grid the detectors’ flags were strongly correlated (mean pairwise correlation 0.76, Table E.2), with an effective number of independent detectors of 1.5 of 6. Most of this correlation comes from the effect size, since every detector flags more often as *δ* grows. For the linear signal at a fixed effect size the effective number was 3.0 at *δ* = 0.2, 2.2 at *δ* = 0.5 and 3.0 at *δ* = 0.8, and 3.7 under the null. Requiring more detectors to agree lowered sensitivity at every effect size without a useful gain in specificity, because the probe alone already had a false positive rate of 0.02 at *p <* 0.05 (Figure 3).

**Figure 3:**
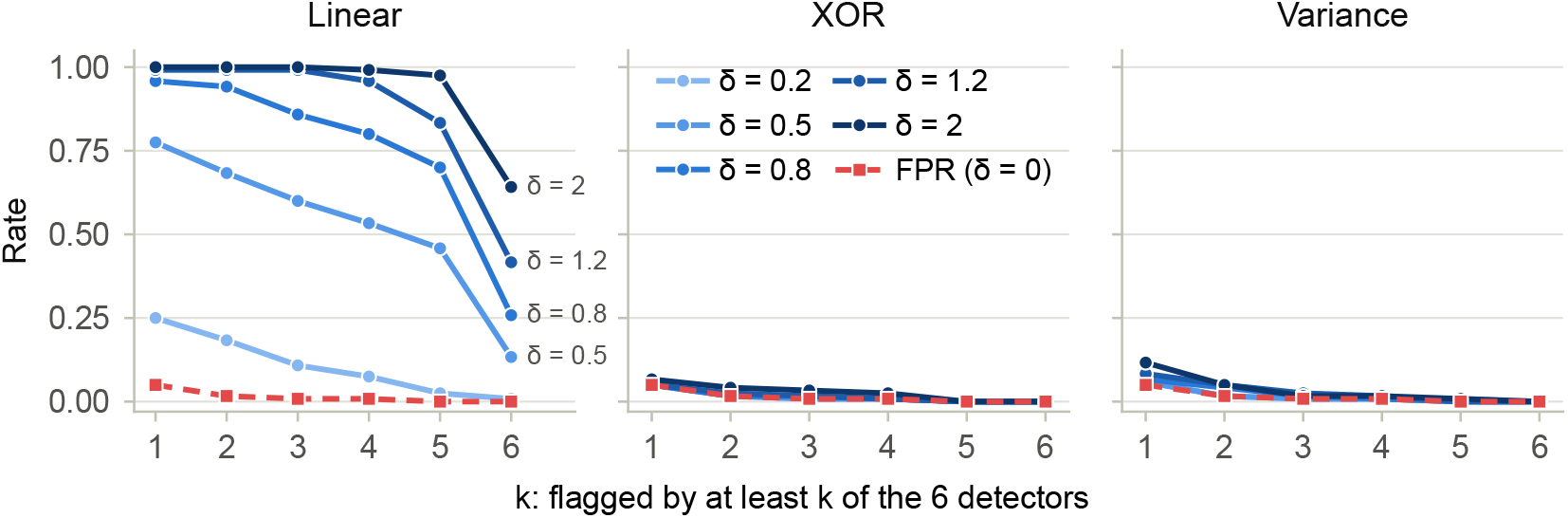
False positive rate at *δ* = 0 and sensitivity at each *δ* of the rule “at least *k* of the six detectors flagged”, for *k* = 1, …, 6, one panel per signal variant. The false positive rate is from the linear-variant null runs.

### 5.2. Clinical audit

*Patient-level calibration*. CheXpert has several images per patient. To see whether that matters, we drew a fake binary attribute at patient level, unrelated to the images, 30 times on the RAD-DINO embeddings (Table F.3). With image-level permutation and ordinary cross-validation, the conditional probe rejected the null in 1.00 of draws, and so did every secondary detector: images of the same patient look alike and share the fake label, so a probe can match them across folds. With patient-level permutation and folds grouped by patient, the probe rejected in 0.00 of draws (0 of 30; the 95% upper bound on its false positive rate is 0.11, so the control rules out the leakage rather than estimating that rate precisely). All CheXpert results below use the patient-level version. Grouping also lowered the observed probe scores. For sex on ResNet-50, for example, the AUROC was 0.841 grouped vs 0.887 plain.

#### Audit results

Every encoder carried attribute information beyond the task label, for every attribute. Tables 6 and 7 give the conditional probe AUROC with its interval and *p*-value and the continuous score of each secondary detector for every encoder and attribute, and Figure 4 shows the same scores. Tables F.11 to F.16 in Appendix F list, for every detector and cell, the statistic, the mean and SD of its permutation null, the raw *p*-value and, for the secondary detectors, the BH-adjusted *q*-value. The conditional probe rejected *H*_0_ in 21 of 21 CheXpert cells and in 12 of 12 MIMIC-CXR cells. Adjusting the probe’s *p*-values across the 21 CheXpert cells by Benjamini– Hochberg leaves all 21 below 0.05 (largest *q* = 0.022), and the same holds across the 12 MIMIC-CXR cells. The MIMIC-CXR ViT-B/32 row uses the file rebuilt from the source (Table F.5 and the comparison with the submitted pipeline below).

**Table 6:** CheXpert audit, task Pneumothorax, first 2,000 images (477 patients), seven frozen encoders (dimensions in Table 4). Conditional probe: out-of-fold AUROC with folds grouped by patient, 95% bootstrap CI over patients (2000 resamples), patient-level permutation *p* (*K* = 500). Secondary detectors: continuous score (*K* = 200). ^*\**^: probe *p <* 0.05, secondary detectors Benjamini–Hochberg *q <* 0.05 across the five within the cell. Flags: number of the six detectors flagged (the probe at *p <* 0.05 plus the secondary detectors at *q <* 0.05). Seed 42.

| Encoder | Attribute | Probe AUROC (95% CI) | *p* | Stat. | Geom. | BiasDir | HBAC-NMI | Freq. | Flags |
|---|---|---|---|---|---|---|---|---|---|
| ResNet-50 | Sex | 0.84 (0.81, 0.87)* | <0.01 | 0.300* | 0.15* | 1.01* | 0.008 | 0.80* | 5/6 |
|  | Age 60+ | 0.75 (0.71, 0.78)* | <0.01 | 0.392* | 0.19* | 0.84* | 0.017* | 0.74* | 6/6 |
|  | White vs non-White | 0.59 (0.54, 0.64)* | <0.01 | 0.282* | 0.15 | 0.60 | 0.012 | 0.61* | 3/6 |
| DenseNet-121 | Sex | 0.88 (0.85, 0.91)* | <0.01 | 0.551* | 0.16* | 1.16* | 0.014 | 0.85* | 5/6 |
|  | Age 60+ | 0.70 (0.65, 0.74)* | <0.01 | 0.627* | 0.22* | 0.84* | 0.039* | 0.76* | 6/6 |
|  | White vs non-White | 0.60 (0.54, 0.64)* | <0.01 | 0.502* | 0.19* | 0.65* | 0.011 | 0.60* | 5/6 |
| ViT-B/16 | Sex | 0.85 (0.82, 0.88)* | <0.01 | 0.462* | 0.14* | 0.99* | 0.011 | 0.72* | 5/6 |
|  | Age 60+ | 0.70 (0.65, 0.75)* | <0.01 | 0.642* | 0.18* | 0.73* | 0.019* | 0.69* | 6/6 |
|  | White vs non-White | 0.60 (0.55, 0.66)* | <0.01 | 0.467* | 0.16* | 0.61 | 0.018 | 0.57* | 4/6 |
| ViT-B/32 | Sex | 0.87 (0.84, 0.89)* | <0.01 | 0.634* | 0.19* | 1.22* | 0.021* | 0.78* | 6/6 |
|  | Age 60+ | 0.67 (0.61, 0.71)* | <0.01 | 0.452* | 0.15* | 0.82* | 0.013* | 0.67* | 6/6 |
|  | White vs non-White | 0.60 (0.55, 0.64)* | <0.01 | 0.362* | 0.14* | 0.69 | 0.009 | 0.60* | 4/6 |
| DINOv2 | Sex | 0.89 (0.86, 0.91)* | <0.01 | 0.581* | 0.15* | 1.40* | 0.011 | 0.80* | 5/6 |
|  | Age 60+ | 0.75 (0.70, 0.79)* | <0.01 | 0.530* | 0.15* | 0.85* | 0.017* | 0.71* | 6/6 |
|  | White vs non-White | 0.59 (0.54, 0.64)* | <0.01 | 0.333 | 0.13 | 0.43 | 0.018 | 0.61* | 2/6 |
| RAD-DINO | Sex | 0.99 (0.99, 1.00)* | <0.01 | 0.635* | 0.21* | 3.60* | 0.053* | 0.97* | 6/6 |
|  | Age 60+ | 0.83 (0.78, 0.87)* | <0.01 | 0.491* | 0.16* | 1.97* | 0.034* | 0.77* | 6/6 |
|  | White vs non-White | 0.61 (0.55, 0.66)* | <0.01 | 0.342 | 0.13* | 1.55* | 0.028 | 0.58 | 3/6 |
| MedSigLIP | Sex | 0.97 (0.95, 0.98)* | <0.01 | 0.792* | 0.31* | 2.68* | 0.021 | 0.97* | 5/6 |
|  | Age 60+ | 0.80 (0.74, 0.85)* | <0.01 | 0.766* | 0.22* | 0.87* | 0.026* | 0.77* | 6/6 |
|  | White vs non-White | 0.57 (0.50, 0.63)* | 0.02 | 0.495 | 0.18 | 0.54 | 0.017 | 0.58 | 1/6 |

**Table 7:**
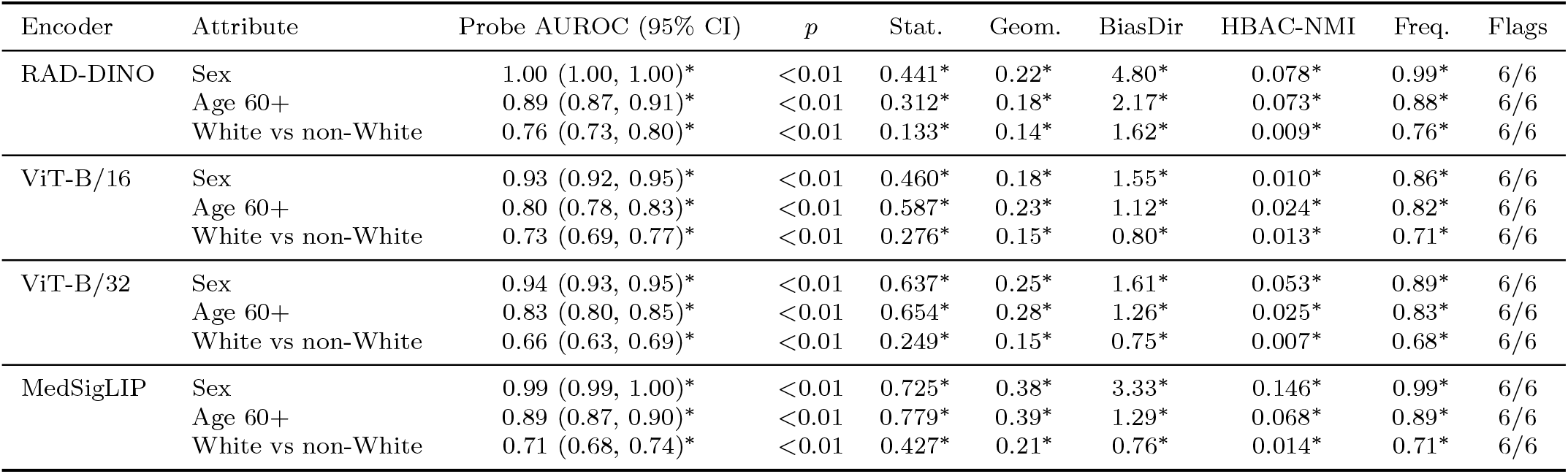
MIMIC-CXR audit, task in-hospital mortality, 1,491 subjects (one image each), four frozen encoders, seed 42. ViT-B/32 uses the file rebuilt from the source in metadata order (Table F.5). Columns as in Table 6.

| Encoder | Attribute | Probe AUROC (95% CI) | *p* | Stat. | Geom. | BiasDir | HBAC-NMI | Freq. | Flags |
|---|---|---|---|---|---|---|---|---|---|
| RAD-DINO | Sex | 1.00 (1.00, 1.00)* | <0.01 | 0.441* | 0.22* | 4.80* | 0.078* | 0.99* | 6/6 |
|  | Age 60+ | 0.89 (0.87, 0.91)* | <0.01 | 0.312* | 0.18* | 2.17* | 0.073* | 0.88* | 6/6 |
|  | White vs non-White | 0.76 (0.73, 0.80)* | <0.01 | 0.133* | 0.14* | 1.62* | 0.009* | 0.76* | 6/6 |
| ViT-B/16 | Sex | 0.93 (0.92, 0.95)* | <0.01 | 0.460* | 0.18* | 1.55* | 0.010* | 0.86* | 6/6 |
|  | Age 60+ | 0.80 (0.78, 0.83)* | <0.01 | 0.587* | 0.23* | 1.12* | 0.024* | 0.82* | 6/6 |
|  | White vs non-White | 0.73 (0.69, 0.77)* | <0.01 | 0.276* | 0.15* | 0.80* | 0.013* | 0.71* | 6/6 |
| ViT-B/32 | Sex | 0.94 (0.93, 0.95)* | <0.01 | 0.637* | 0.25* | 1.61* | 0.053* | 0.89* | 6/6 |
|  | Age 60+ | 0.83 (0.80, 0.85)* | <0.01 | 0.654* | 0.28* | 1.26* | 0.025* | 0.83* | 6/6 |
|  | White vs non-White | 0.66 (0.63, 0.69)* | <0.01 | 0.249* | 0.15* | 0.75* | 0.007* | 0.68* | 6/6 |
| MedSigLIP | Sex | 0.99 (0.99, 1.00)* | <0.01 | 0.725* | 0.38* | 3.33* | 0.146* | 0.99* | 6/6 |
|  | Age 60+ | 0.89 (0.87, 0.90)* | <0.01 | 0.779* | 0.39* | 1.29* | 0.068* | 0.89* | 6/6 |
|  | White vs non-White | 0.71 (0.68, 0.74)* | <0.01 | 0.427* | 0.21* | 0.76* | 0.014* | 0.71* | 6/6 |

**Table 8:** Classification-head experiment on frozen CheXpert embeddings, one row per attribute and task, mean over the seven encoders and 10 seeds. Head: L2 logistic regression on standardised embeddings, trained at *ρ*_train_ = 0.9 and tested at *ρ* = 0.5 on patient-disjoint halves. Probe: conditional probe AUROC on all labelled images of the task, one value per cell. TPR and FPR gap between groups (test). AUROC drop versus an unconfounded head of equal training size. Output shift: mean absolute change in head output when the attribute direction is removed. AUROC on aligned and misaligned test images. Expected calibration error (ECE) per group.

| *A* | *Y* | *n* train<br>(min-max) | *n* test<br>(min-max) | Probe | TPR<br>gap | FPR<br>gap | AUROC<br>drop | Output<br>shift | AUROC<br>aligned | AUROC<br>misaligned | ECE<br>*A*=1 | ECE<br>*A*=0 |
|---|---|---|---|---|---|---|---|---|---|---|---|---|
| Age 60+ | Cardiomegaly | 54-104 | 68-128 | 0.73 | 0.12 | 0.32 | -0.02 | 0.20 | 0.89 | 0.62 | 0.287 | 0.202 |
| Age 60+ | Pleural Effusion | 228-272 | 244-288 | 0.73 | 0.19 | 0.13 | 0.00 | 0.22 | 0.93 | 0.76 | 0.151 | 0.187 |
| Age 60+ | Pneumothorax | 42-132 | 60-220 | 0.76 | 0.13 | 0.16 | 0.02 | 0.25 | 0.71 | 0.49 | 0.332 | 0.329 |
| Sex | Cardiomegaly | 42-80 | 68-136 | 0.87 | 0.17 | 0.17 | 0.03 | 0.20 | 0.86 | 0.64 | 0.243 | 0.236 |
| Sex | Pleural Effusion | 134-222 | 244-404 | 0.90 | 0.21 | 0.23 | 0.04 | 0.20 | 0.93 | 0.67 | 0.190 | 0.197 |
| Sex | Pneumothorax | 86-168 | 64-276 | 0.89 | 0.29 | 0.24 | 0.02 | 0.27 | 0.82 | 0.46 | 0.286 | 0.318 |

**Figure 4:**
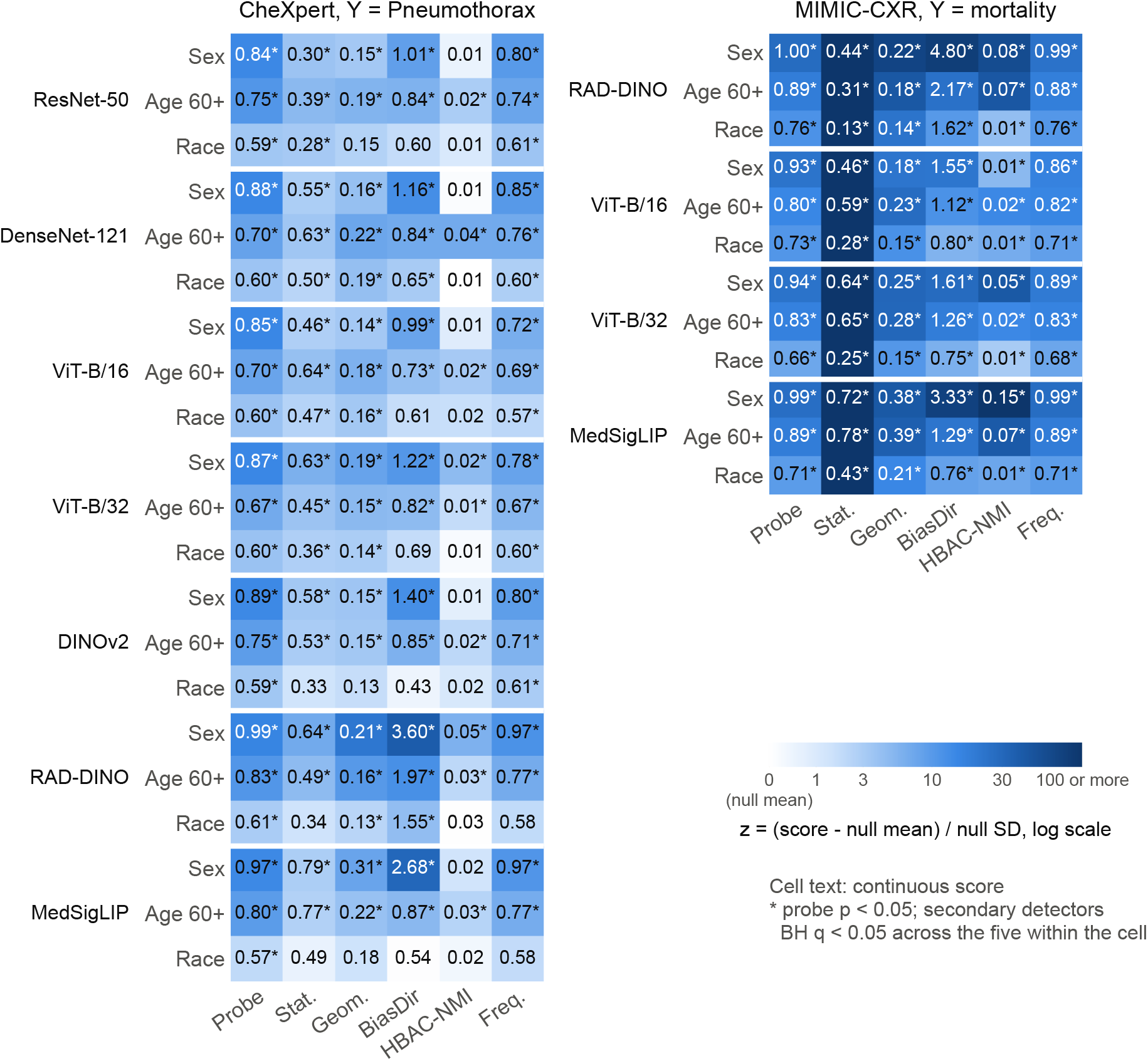
Detector scores for every encoder and attribute on CheXpert and MIMIC-CXR. Colour: *z*-score of the observed statistic against its own permutation null, (*s −* null mean)/null SD, on a log scale. Cell text: the continuous score. ^*\**^: probe *p <* 0.05, secondary detectors BH *q <* 0.05 across the five within the cell. Colour scale is colour-blind safe.

Sex had the largest potential, with a conditional probe AUROC of 0.84 to 0.99 on CheXpert and 0.93 to 1.00 on MIMIC-CXR, where RAD-DINO reached 1.00 (95% CI 1.00 to 1.00). For age the ranges were 0.67 to 0.83 on CheXpert and 0.80 to 0.89 on MIMIC-CXR. Race differed between the cohorts: 0.57 to 0.61 on CheXpert, where the MedSigLIP cell was the weakest and is flagged by the primary test alone (*p* = 0.022, none of its five secondary detectors flagged). Had the probe been adjusted together with the secondary detectors, it would not have been (*q* = 0.13), and this is the only cell on which the two rules differ. Race potential was higher at 0.66 to 0.76 on MIMIC-CXR. MedSigLIP-448, re-extracted for this revision, was among the strongest CheXpert encoders for sex and age (0.97 and 0.80). The unidentified array used under that name in the first submission had been the weakest for every attribute.

#### Stability of the race result

Table F.8 subsamples the non-White group to fixed sizes on RAD-DINO, ten draws per size. On CheXpert the probe rejected in 0.10 of draws with 25 non-White images and in 0.80 with 400, and its AUROC varied widely between draws (SD 0.16 at the smallest size). On MIMIC-CXR the rejection rate was 0.80 at 25 images and 1.00 at 400. The CheXpert race signal is weak: the probe rejected in 7 of 10 draws with 200 non-White images and in 8 of 10 with 400, but rarely with 100 or fewer, whereas the MIMIC-CXR signal is detected with 50. Keeping race groups with at least 50 images as separate groups (White, Black and Asian on CheX-pert, and White and Black on MIMIC-CXR, which has 38 Asian subjects) gave somewhat higher AUROCs than the binary split (Table F.1), and four age bins gave somewhat lower ones than the binary split (Table F.2).

#### Sensitivity of the audit

Table F.7 repeats the CheXpert probe with three changes. On the 1577 frontal images alone no AUROC moved by more than 0.03, and 21 of 21 cells were significant, against 21 in the main audit. Keeping only the 802 images with a definite Pneumothorax label, as in the head experiment, no AUROC moved by more than 0.06, and 20 of 21 cells had *p <* 0.05, with MedSigLIP race, the weakest cell of the main audit, the one exception. Setting the probe’s regularisation to *C* = 0.1 or *C* = 10 changed the significance decision in 0 of 42 cells. With the arrays used in the first submission the four re-extracted encoders gave AUROCs up to 0.29 away from the main result, almost all of it from the unidentified MedSigLIP array. Appending the squared coordinates to the probe’s features changed the decision in 0 of 33 clinical cells and raised the AUROC by at most 0.02 (Appendix E, Table E.4).

#### What the embeddings cluster on

To see whether the embeddings carry structure that the named attributes do not explain, we compared *k*-means clusters (*k* = 8) with all recorded metadata (Table F.9). On both cohorts the clusters followed image acquisition most closely: the strongest association on CheXpert was with AP/PA and on MIMIC-CXR with view (AP/PA), and on CheXpert the frontal-versus-lateral view had a mean normalised mutual information of 0.378 with the clusters. Support devices came next on CheX-pert (0.068, range 0.052 to 0.103). Sex, age and race were weakly associated with the clusters even where the probe recovered them well, so these attributes are encoded in directions that do not dominate the overall structure of the embedding.

#### Comparison with the submitted pipeline

Rerunning the submitted 13-method pipeline with only the flag reader fixed changed its clinical tables (Table F.10). The statistical, geometric and GCE detectors, which the submitted tables showed as never flagging, flagged in every CheXpert cell once their results were read correctly, and the mean agreement count on CheXpert rose from 7.0 to 9.4 of 13 (MIMIC-CXR: 5.2 to 7.5). The identical age and race rows of the submitted CheXpert table, present for 6 of 7 encoders, came from detectors that fitted on the task label and so returned the same output for every attribute. These comparisons describe the old pipeline only, and none of its outputs is used elsewhere in this paper. One input of the submitted pipeline was also replaced. The submitted MIMIC-CXR ViT-B/32 file failed two independent checks (Table F.5): it held the first 1,491 rows of its source, taken before rows without an outcome were removed. Its similarity structure matched no other encoder (largest Spearman correlation 0.006), and its sex probe was at chance (AUROC 0.50, against 0.93 for ViT-B/16 on the same images). The file rebuilt from the source passed both (0.499 and 0.94), and every MIMIC-CXR result in this paper uses it.

### 5.3. Classification-head experiment

Across the 42 encoder, attribute and task cells, heads trained on confounded data relied on the attribute (Table 8). The mean TPR gap between groups on the balanced test set was 0.19. On test images where attribute and label disagreed with the training correlation, the head’s AUROC was 0.61, against 0.85 where they agreed. A head trained on unconfounded data of the same size scored 0.75 and 0.77. The drop in overall test AUROC was smaller (mean 0.02), because the balanced test set mixes aligned and misaligned images. Calibration error was similar for the two groups (0.248 and 0.245).

The audit score correlated with the size of this reliance: across cells, the Spearman correlation between the conditional probe AUROC and the TPR gap was 0.60 (95% CI 0.33 to 0.79, or 0.31 to 0.80 when whole encoders are resampled, and partial correlation given attribute and task 0.63). With the FPR gap it was 0.28 (*p* = 0.072), and with the AUROC drop 0.72 (Figure 5). Within attributes the correlation with the TPR gap was 0.74 for sex (interval 0.19 to 0.94 when encoders are resampled, *p <* 0.001) and 0.19 for age (−0.29 to 0.57, *p* = 0.417), so the relation was clear across encoders for sex and uncertain within age. Leaving out any one encoder, the correlation ranged from 0.50 (without RAD-DINO) to 0.68, and leaving out any one task from 0.45 (without Pneumothorax) to 0.73. With the mean of the permutation null subtracted from each probe score, which removes the part a head could get from *Y* alone, the correlation was 0.57 (0.75 within sex, 0.09 within age). With a multilayer perceptron head (one hidden layer of 256 units) trained on the same splits the correlation was 0.53 (interval 0.20 to 0.76 when encoders are resampled), 0.70 within sex and 0.09 within age, and the two heads’ TPR gaps correlated at 0.96 across cells.

**Figure 5:**
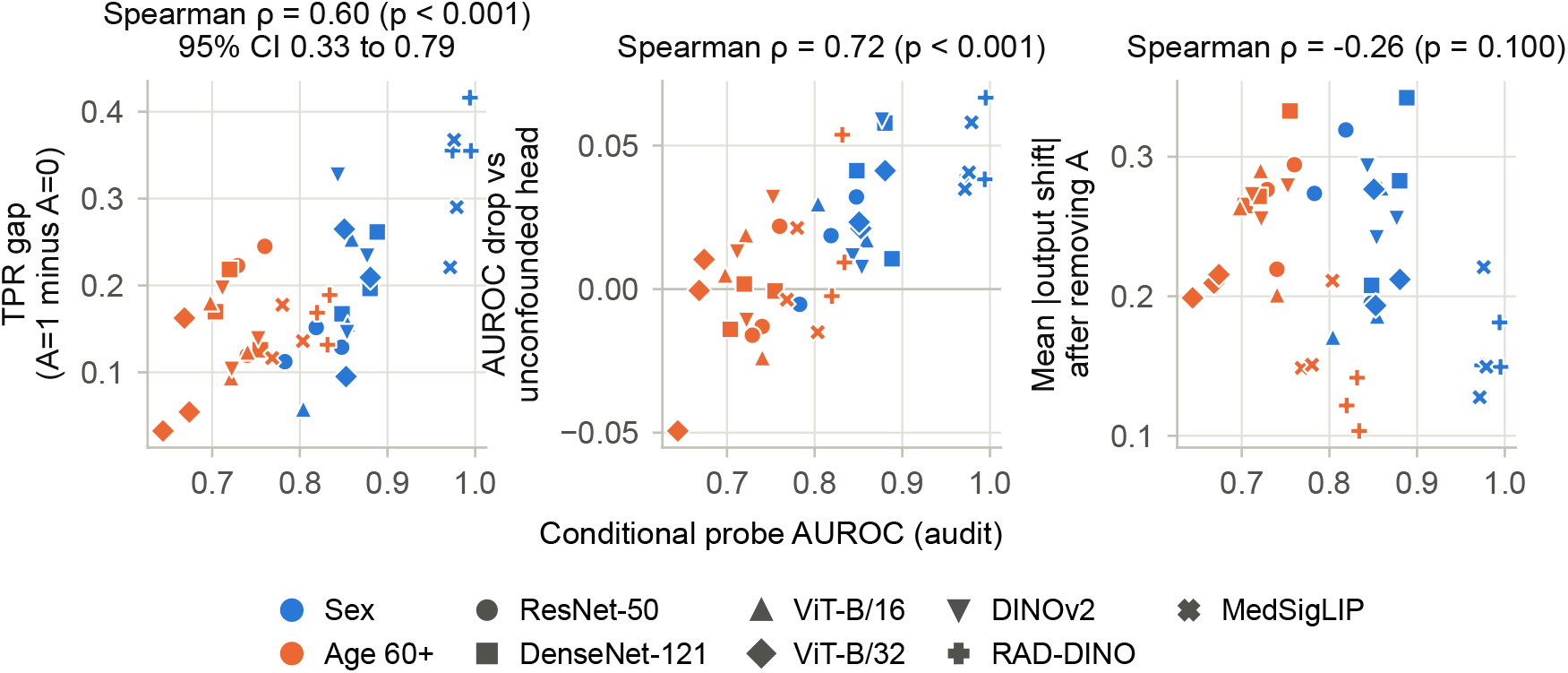
Conditional probe AUROC against the TPR gap, the AUROC drop and the output shift (not used as a measure of reliance, Appendix C), one point per encoder, attribute and task cell on CheXpert (42 cells; colour marks the attribute, marker shape the encoder), with Spearman correlations over all cells. Within attributes the correlation with the TPR gap is 0.74 for sex and 0.19 for age (Section 5.3).

With race as the attribute (White against non-White), the probe AUROCs spanned only 0.52 to 0.65, the confounded heads showed no consistent gap, and the correlation across the 21 cells was 0.03 (Table C.2 in Appendix C).

#### Mitigation

At the package defaults, none of the three mitigations that alter the embedding reduced the head’s TPR gap by more than 0.01 beyond its control, and only contrastive debiasing lowered the probe score beyond its control, by a small margin (Table 9). Paired by cell and seed, the method-minus-control differences in TPR gap were −0.01 for adversarial debiasing (−0.03 to 0.01 when whole encoders are resampled), 0.00 for contrastive debiasing (−0.03 to 0.02) and −0.01 for masking (−0.03 to −0.00); RAD-DINO was the one encoder on which all three lowered the gap, by about 0.06 relative to its controls. With adversarial debiasing the mean conditional probe AUROC changed by −0.09, and the number of audits with a significant probe went from 123 to 100 of 126. The control network without the adversary changed it by −0.09, which leaves −0.01 for the adversary itself. The contrastive debiasing method lowered the probe further, a change of −0.16 against −0.12 for its control, a difference of −0.04. Significant audits went from 123 to 70, and to 85 for its control. Relative to their controls, the adversarial and contrastive methods changed the task AUROC by 0.00 and 0.00 and the confounded head’s TPR gap by −0.01 and 0.00. Most of the lower audit score came from passing the embeddings through a small learned debiasing network, and in every case the confounded head found the attribute signal that remained.

**Table 9:** Conditional probe AUROC, number of audits with a significant probe (of 63 per attribute: seven encoders, three tasks, 3 seeds), task AUROC and TPR gap of the confounded head before and after each mitigation (package defaults) and its control. Adversarial and contrastive control: same debiasing network and training, mitigation term weight 0. Masking control: the same number of random dimensions set to zero. DFR refits the head only, so its probe columns repeat the raw embedding. Probe columns are computed on the test half of the patients. CheXpert, mean over cells and seeds.

| Method | *A* | Probe before | Probe after | Sig. before | Sig. after | Task AUROC before | Task AUROC after | TPR gap before | TPR gap after |
|---|---|---|---|---|---|---|---|---|---|
| Adversarial debiasing | Sex | 0.85 | 0.72 | 63/63 | 53/63 | 0.73 | 0.74 | 0.22 | 0.21 |
|  | Age 60+ | 0.71 | 0.66 | 60/63 | 47/63 | 0.73 | 0.73 | 0.14 | 0.15 |
| Same encoder, adversary weight 0 | Sex | 0.85 | 0.74 | 63/63 | 59/63 | 0.73 | 0.74 | 0.22 | 0.23 |
|  | Age 60+ | 0.71 | 0.65 | 60/63 | 44/63 | 0.73 | 0.73 | 0.14 | 0.15 |
| Contrastive debiasing | Sex | 0.85 | 0.64 | 63/63 | 43/63 | 0.73 | 0.72 | 0.22 | 0.22 |
|  | Age 60+ | 0.71 | 0.60 | 60/63 | 27/63 | 0.73 | 0.73 | 0.14 | 0.14 |
| Same encoder, contrastive weight 0 | Sex | 0.85 | 0.69 | 63/63 | 48/63 | 0.73 | 0.73 | 0.22 | 0.22 |
|  | Age 60+ | 0.71 | 0.62 | 60/63 | 37/63 | 0.73 | 0.72 | 0.14 | 0.14 |
| Shortcut masking | Sex | 0.85 | 0.84 | 63/63 | 63/63 | 0.73 | 0.74 | 0.22 | 0.20 |
|  | Age 60+ | 0.71 | 0.70 | 60/63 | 58/63 | 0.73 | 0.73 | 0.14 | 0.14 |
| Same number of random dimensions | Sex | 0.85 | 0.85 | 63/63 | 63/63 | 0.73 | 0.73 | 0.22 | 0.22 |
|  | Age 60+ | 0.71 | 0.71 | 60/63 | 59/63 | 0.73 | 0.73 | 0.14 | 0.15 |
| Last-layer retraining (DFR) | Sex | 0.85 | 0.85 | 63/63 | 63/63 | 0.73 | 0.66 | 0.22 | 0.01 |
|  | Age 60+ | 0.71 | 0.71 | 60/63 | 60/63 | 0.73 | 0.64 | 0.14 | 0.01 |

Masking the top 5% of attribute dimensions barely moved the probe (−0.01, against 0.00 for random dimensions), because the attribute is spread over many dimensions, and relative to its control it changed the TPR gap by −0.01. Refitting the head on a group-balanced subset was the only method that lowered the gap, from 0.18 to 0.01 on average and in 39 of 42 cells (Wilcoxon signed-rank test on cell means, *p <* 0.001). Task AUROC fell from 0.73 to 0.65. The balanced subset is limited by the rarest (*A, Y*) group of the confounded training set and held only 8 to 52 images. A control in Appendix C traces the loss in about equal parts to this small size and to the L1 penalty of the package’s DFR head, and none of it to balancing. Its audit score is unchanged by construction, since the embedding is the same. Table C.1 in Appendix C gives the changes in TPR gap and task AUROC of each method for each encoder.

#### Planted marker

With the marker on Pneumothorax-positive images at rate 0.5, the conditional probe AUROC for the marker was 0.999, and the probe flagged it in all 10 seeds (largest *p* = 0.005, Table 11). A head trained with the marker on 0.9 of positive training images used it: on test images, its TPR gap between marked and unmarked images was 0.94 and its FPR gap 0.90.

### 5.4. CelebA

For *A* = Male, the conditional probe AUROC was 0.98 given Blond hair, 0.99 given Heavy makeup and 0.98 given Attractive, and all six detectors flagged each task (Table 10).

**Table 10:** CelebA audit, ResNet-50 (2,048-dim), 10,000 images grouped by identity, *A* = Male, one row per task *Y*, seed 42. Columns as in Table 6.

| Task (*Y*) | *n* | *Y*=1 M/F | Probe AUROC (95% CI) | Stat. | Geom. | BiasDir | HBAC-NMI | Freq. | Flags |
|---|---|---|---|---|---|---|---|---|---|
| Blond Hair | 10000 | 76/1426 | 0.98 (0.98, 0.99)* | 0.952* | 0.73* | 2.76* | 0.243* | 0.98* | 6/6 |
| Heavy Makeup | 10000 | 12/3872 | 0.99 (0.98, 0.99)* | 0.901* | 0.57* | 2.41* | 0.119* | 0.99* | 6/6 |
| Attractive | 10000 | 1191/3904 | 0.98 (0.98, 0.98)* | 0.964* | 0.70* | 2.85* | 0.234* | 0.98* | 6/6 |

**Table 11:** Planted marker on CheXpert, ResNet-50 (2,048-dim), task Pneumothorax, 10 seeds. Detector rows: mean score, largest permutation *p* over seeds, and the number of seeds flagged (probe at *p <* 0.05, secondary detectors at BH *q <* 0.05 across the five). Head rows: TPR and FPR gap between marked and unmarked test images.

|  | Mean score | Max *p* | Flagged |
|---|---|---|---|
| Probe | 0.999 | 0.005 | 10/10 |
| Stat. | 0.344 | 0.005 | 10/10 |
| Geom. | 0.450 | 0.005 | 10/10 |
| BiasDir | 3.120 | 0.005 | 10/10 |
| HBAC-NMI | 0.361 | 0.010 | 10/10 |
| Freq. | 1.000 | 0.005 | 10/10 |
| Head TPR gap | 0.94 | n/a | n/a |
| Head FPR gap | 0.90 | n/a | n/a |

## 6. Discussion

### 6.1. What the audit measures, and how far it predicts use

Shortcut potential can be measured on the embedding alone, which answers the first question of the Introduction: on synthetic data the conditional probe held its stated error rate, including the case where *A* and *Y* are strongly correlated and an unconditional probe is wrong almost every time. That answer covers signals that shift group means. No detector saw the XOR or variance-shift signals (Section 5.1), although probes on squared or product features under the same null did (Appendix E). The answer to the second question is partial: on CheXpert, encoders with higher conditional probe scores produced heads with larger subgroup gaps when the training data were confounded, with a rank correlation of 0.60 (interval 0.31 to 0.80 when whole encoders are resampled). Within attributes the relation was strong for sex (0.74) and weak for age (0.19). One reason is range restriction: the mean TPR gap of the age cells varied little between cells (SD 0.05, against 0.10 for sex) while seeds within a cell varied by 0.14, so the ranking of age cells is largely seed noise.

For a linear head trained on confounded data, shortcut potential measured on the embedding was therefore informative about use, clearly for sex, not significantly for age. For race, whose potential on CheXpert was weak for every encoder, the heads showed no consistent gap and the relation was not detectable over that range (0.03, Appendix C). It does not establish use. The correlation is across cells, and it was measured for logistic and small MLP heads under one form of confounding, on training sets of 42 to 272 images drawn from a CheXpert sample that is not random. A head trained on unconfounded data showed almost no gap between aligned and misaligned images on the same embeddings. Whether an attribute is used depends on the training data and the head, as Brown et al. [10], Petersen et al. [13] and Kina and Petersen [14] argue. Our aligned and misaligned AUROCs repeat the pattern Kina and Petersen found for a chest-drain shortcut on frozen foundation models.

The mitigation results add a caution: at the package defaults, adversarial and contrastive debiasing lowered the conditional probe score, but the same debiasing networks trained without the mitigation term lowered it nearly as much (for contrastive debiasing the difference from its control was −0.04), and masking the top attribute dimensions did little more than masking random ones. None of the three moved the confounded head’s TPR gap by more than 0.01 beyond its control. A head trained on confounded data found the attribute signal that remained. The one method that closed the gap, refitting the head on group-balanced data, left the embedding and its audit score unchanged and cost task AUROC, for reasons unrelated to balancing (Appendix C). Both observations agree with the view that much of shortcut use on frozen encoders is decided in the head [14], and with MEDFAIR’s finding that mitigation often does not beat plain training [46]. An audit score can fall for reasons that have nothing to do with debiasing, so a mitigation needs a control, and the head has to be tested.

A positive audit for general-purpose encoders is unsurprising, since encoders pretrained on large image corpora are built to retain most of what is visible. Sex and age leave visible traces in chest radiographs, and earlier work recovered race from chest radiographs, which radiologists cannot do [8, 11], and sex, site and scanner from knee radiographs almost perfectly [12]. For sex in particular, part of the signal is likely anatomical. The audit cannot separate legitimate anatomical information from unwanted information, because both are information about *A* given *Y*. Subgroup separability sets an upper limit on harm without implying it [32], and the audit should be read in that sense.

### 6.2. Relation to earlier findings

Our probe results agree in direction with the encoding of sex, age and race reported by Gichoya et al. and Glocker et al. on full chest X-ray models [8, 11], though our numbers are not directly comparable, because we audit frozen embeddings on small samples and condition on a task. The correlation between audit score and head gap is a frozen-embedding counterpart of the relation Yang et al. reported between demographic encoding and fairness gaps [34]. Dependence measures have so far been used as training penalties [19, 20]. Calibrated by permutation, the same idea also works as a check on a frozen encoder.

### 6.3. What the secondary detectors add

On this evidence they add very little: their flags rose and fell with the probe’s as the effect size changed (effective number 1.5 of 6 over the whole grid, 2.2 at a fixed *δ* = 0.5), and requiring several of them to agree only cost sensitivity. None of them detected the XOR or variance-shift signals that the probe missed. In the submitted version we read disagreement between detectors as evidence about the geometry of the encoding. Most of that disagreement came from code faults, and the remainder is explained by differences in power. Combining detectors resembles ensembling [63] or a mixture of experts [64], but ensembles gain from roughly independent errors, which these detectors have only in part, and a mixture of experts would need a gate trained on labelled shortcuts. We keep the secondary detectors as diagnostics because their scores are on interpretable scales (fraction of dimensions, centroid distance), and they play no part in the decision.

### 6.4. Cohorts, acquisition and data handling

Race potential was weak on CheXpert and clearer on MIMIC-CXR. The cohorts differ in task and sampling, and in the make-up of the non-White group, so we do not read this as an institution effect. The clusters followed view and projection on both cohorts, consistent with earlier reports that hospital [6] and scanner [7] signals, and site signals in knee radiographs [12], dominate what image models pick up.

Several of the problems we found were data-handling faults. Repeated images of one patient made every detector reject a meaningless attribute until permutation and folds were grouped by patient. One MIMIC-CXR embedding file did not match its metadata, and the submitted tables reported shortcut flags for it. The CheXpert sex column was inverted in the submitted runs. The first two can be caught cheaply, with a fake-attribute control and a row-alignment check, and we recommend running them before any audit.

## 7. Limitations

The audit, and every secondary detector, missed the two attribute signals we tested that leave group means unchanged: on the XOR and variance-shift variants no detector did better than chance. The gap lies in the statistics, which are built around differences in location: under the same permutation null, probes on squared coordinates and on pairwise products detected both signals with their calibration intact (Appendix E), and on the clinical cohorts the squared-feature probe changed no decision (Table E.4). A linear probe was a deliberate choice, since it matches how frozen embeddings are usually used, but a nonlinear head could exploit signal that the audit reports as absent. A negative audit is therefore weaker evidence than a positive one.

The audit needs labelled sensitive attributes, which are often missing or recorded as imperfect proxies. It examines one embedding at a time, from one layer of one encoder, and says nothing about other layers or about the encoder after fine-tuning. For age the score was not significantly related to reliance, so on this cohort the audit met its own usefulness criterion clearly for sex only. The head experiment covers two head types (logistic regression and a small multilayer perceptron) and one form of confounding, with two attributes (race, whose signal on CheXpert is weak, only as an additional run in Table C.2) and three tasks on one cohort, and the correlation between audit and reliance is computed across only 42 cells that share encoders. With seven encoders, the interval that resamples them is coarse. The heads pool frontal and lateral views, and on frontal views alone the audit AUROCs moved by at most 0.03.

The CheXpert sample is the first 2,000 images in the order of the official training file, not a random sample, so the CheXpert results may not hold for the full training set and may carry whatever ordering the file was assembled with. Against the full training file the sample has a similar sex and race mix (43% against 41% female, and 64.5% against 63.8% White among images with a recorded race) and slightly younger patients (median age 59 against 62). It also has more lateral views (21% against 14%), which the frontal-only analysis bounds.

The MIMIC-CXR task, in-hospital mortality, is an outcome with many causes outside the image. MIMIC-CXR also has radiological labels, which we did not use as tasks, and the audit’s result is specific to the task it conditions on. Both samples are small, which limits power for small groups. The CheX-pert race result in particular is rarely detected with 100 or fewer non-White images. Collapsing race into White against non-White hides differences between groups. Records with race unknown, declined or not obtained are left out on both cohorts (217 CheXpert images, 71 MIMIC-CXR subjects). The head-experiment cells are not independent and some are small (42 to 272 training and 60 to 404 test images). The identity of the MIMIC-CXR encoders could not be verified from code. On CheXpert the four re-extracted encoders replaced arrays whose origin is unknown, and the old and new arrays differ (Table F.6). RAD-DINO was pretrained on nearly all the images audited here, and for the other encoders the pretraining data could not be checked image by image. MedSigLIP was trained on medical images paired with reports, MIMIC-CXR among them, so its text side could have taught it findings and demographics that reports mention, and its MIMIC-CXR scores, like RAD-DINO’s, may describe an encoder that saw the audited images in pretraining. CheXpert is not among the public data sets its model card lists, and the card does not itemise its licensed data sets.

The Benjamini–Hochberg correction is applied across the five secondary detectors within each cell, and the primary probe is judged at *p <* 0.05 in each cell, with no adjustment across cells for either. SIS was removed from the score, and methods that need training access were not evaluated. We compared embedding clusters with recorded metadata but did not run an error-aware slice discovery method such as GEORGE or Domino [42, 43], so subgroups on which a head fails without a recorded attribute were not searched for. The mitigations were run only on CheXpert and only at the package defaults, without tuning, so tuned settings could change the result, and the planted marker is a single, simple artefact on one encoder.

## 8. Conclusion

We asked whether shortcut potential can be measured on a frozen embedding, and whether the measurement predicts how strongly a head trained on that embedding with confounded data relies on the attribute. *ShortKit-ML* measures the shortcut potential of a frozen embedding for a task with one primary test, a conditional probe calibrated by permuting the attribute within task strata and, where a patient has several images, between patients. The test held its error rate on synthetic data and found a planted marker in every seed, but like every detector here it missed signals that leave group means unchanged, so a negative audit does not rule out a shortcut that a nonlinear head could use.

On CheXpert, its score correlated with how strongly a logistic head trained on confounded data relied on the attribute (Spearman 0.60, 0.74 within sex, not significant within age), but at the package defaults three embedding-level mitigations did no better than their controls at reducing that reliance. Refitting the head on a group-balanced subset closed that gap, in 39 of 42 cells, without changing the audit score. Task AUROC fell because that subset was small and because the package’s DFR head uses an L1 penalty. The audit is a first check on an encoder, and the head trained on it has to be tested as well. Two cheap checks caught our own data faults and belong before any audit: a fake patient-level attribute that no detector should find, and a row-alignment check between encoders.

Three extensions would make the audit more useful. Probes on squared and product features found the XOR and variance-shift signals that no detector here found (Appendix E). Making such tests part of the audit, with product features at encoder dimension, is the next step, since on the clinical cohorts the squared probe changed no decision. The link between audit score and reliance should be checked on other cohorts and with larger heads, and under confounding other than a single correlated attribute. Auditing an encoder on images it was pretrained on and on images it never saw would show how much of the potential comes from the pretraining data. The audit is intended as one of the embedding-level checks in DOJO, a community-driven platform for adversarial evaluation of health AI [65].

The wider lesson is about what can be measured. A shortcut can only be audited through an attribute someone recorded, and most of what an encoder has learned from its training data was never recorded. This toolkit is a first step for the case where the attribute is known and the input is a frozen embedding. The harder cases, confounders with no label, signals that no linear test sees, and inputs beyond embeddings (the image itself, text and multimodal encoders) need tools that do not yet exist, and we see this work as a map of which ones to build next.

## Data Availability

All code, configuration and the scripts that generate every table and figure are at https://github.com/criticaldata/ShortKit-ML. CheXpert and MIMIC-CXR must be obtained from their providers under their data use agreements; CelebA is public. The embeddings and metadata needed to rerun the analyses, including the MIMIC-CXR subset and the files it was drawn from, are on a gated Hugging Face dataset (https://huggingface.co/datasets/MITCriticalData/ShortKit-ML-data), released to credentialed PhysioNet accounts with MIMIC-CXR access; no images are shared. CheXpert and CelebA embeddings are extracted from pinned public checkpoints by scripts in the repository. No new patient data were collected.

https://huggingface.co/datasets/MITCriticalData/ShortKit-ML-data

https://physionet.org/content/mimic-cxr/

https://stanfordmlgroup.github.io/competitions/chexpert/

https://mmlab.ie.cuhk.edu.hk/projects/CelebA.html

## Ethics Statement

This study used three publicly available datasets: CheXpert, MIMIC-CXR, and CelebA. CheXpert was released by the Stanford Machine Learning Group under a research data use agreement and consists of deidentified chest radiographs. MIMIC-CXR was released by the Massachusetts Institute of Technology Laboratory for Computational Physiology under the PhysioNet Credentialed Health Data License 1.5.0. Access to MIMIC-CXR was obtained through completion of the required CITI Data or Specimens Only Research training (PhysioNet credentialed user). CelebA is a publicly available facial attribute dataset released for academic research. No new human subjects data were collected for this study. As all data used had been collected and deidentified before their release through established public repositories under their respective data use agreements, this work did not require additional institutional review board approval. The study was conducted in accordance with the principles of the Declaration of Helsinki.

## Data and code availability

Code, configuration and the scripts that generate every table and figure are at https://github.com/criticaldata/ShortKit-ML. CheXpert and MIMIC-CXR must be obtained from their providers under their data use agreements. The embeddings and metadata needed to rerun the analyses, including the MIMIC-CXR subset and the files it was drawn from, are on a gated Hugging Face dataset (https://huggingface.co/datasets/MITCriticalData/ShortKIT-ML-data), released to credentialed PhysioNet accounts with MIMIC-CXR access; no images are shared. CheXpert embeddings are extracted from pinned public checkpoints by a script in the repository (Table F.6), and the CelebA embeddings by another script there from the public images. The RAD-DINO training image list is published with its model card (https://huggingface.co/microsoft/rad-dino).

## Acknowledgments

The authors thank the MIT Critical Data community for support and discussion. Computational resources for embedding extraction were provided by the MIT Office of Research Computing and Data (ORCD) through A100 and H200 GPU allocations, and the audits and head experiments ran on a laptop CPU. The revision was supported in part by Anthropic’s AI for Science program.

## Declaration of generative AI and AI-assisted technologies in the manuscript preparation process

During the preparation of this work the authors used Claude (Anthropic) in order to draft the first version of the manuscript and to review the code. MIMIC-CXR data stayed on institutional machines and were not given to Claude or any other AI service, and model training on the authors’ conversations was disabled. After using this tool, the authors reviewed and edited the content as needed and take full responsibility for the content of the published article.

## Appendix A. Detector definitions

Notation: for each image *i, E*_*i*_ *∈* ℝ^*d*^ is its standardised embedding, with attribute group *a*_*i*_ and task label *y*_*i*_. *S*_*y*_ = *{i* : *y*_*i*_ = *y}* is a task stratum and *S*_*y,a*_ = *{i* : *y*_*i*_ = *y, a*_*i*_ = *a}* a cell. Statistics averaged over strata are weighted by |*S*_*y*_|. All six statistics are calibrated by the conditional permutation null of Section 3.2. The null recomputes the whole statistic, including any fitted direction, ranking or clustering, on each permuted data set. **Conditional probe**. L2-regularised logistic regression on [*E*_*i*_, *y*_*i*_], regularisation strength 1, 5-fold CV stratified on (*a, y*) and grouped by patient (CheXpert) or identity (CelebA), averaged over five fold splits. Its statistic *s* is the out-of-fold AUROC for *A* (macro one-vs-rest when *G >* 2) [17, 18].

**Statistical**. For each dimension *j* and stratum *y*, a Mann–Whitney *U* test (*G* = 2) or Kruskal–Wallis test (*G >* 2) of *E*_*ij*_ across groups within *S*_*y*_. All *d ×* |*Y*| *p*-values are adjusted by Benjamini–Hochberg at 0.05 [51], and *s* is the fraction of dimensions significant in at least one stratum.

**Geometric**. Within stratum *y*, with group centroids *µ*_*y,a*_,

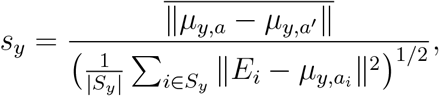

the mean pairwise centroid distance over the pooled within-group spread. The statistic *s* is the weighted mean of *s*_*y*_.

**Bias direction PCA**. Within stratum *y, v*_*y*_ is the first principal component of the group-mean differences (for *G* = 2, the normalised difference of the two means) [35]. With projections 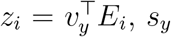 is the largest absolute difference of group means of *z* divided by the pooled standard deviation of *z*. Because *v*_*y*_ is fitted on the same data, *s*_*y*_ is positive even under the null. The permutation null accounts for this, which a fixed threshold did not.

**HBAC**. Within stratum *y, k*-means with *k* = 8 clusters on *E*, with *s*_*y*_ the normalised mutual information between cluster labels and *A* [44, 52].

**Frequency**. Within each training fold, dimensions are ranked by their absolute within-stratum standardised group difference (one ranking per one-vs-rest target when *G >* 2, as for the probe) and the top 5% are kept; *s* is the out-of-fold AUROC of the conditional probe restricted to those dimensions [53]. The values of *k*, the top fraction and the probe’s *C* are package defaults fixed before any result was seen and were not tuned. Table F.7 reports the probe’s sensitivity to *C*.

## Appendix B. Package methods outside the audit score

The package implements 20 methods. Table 2 covers the six scored detectors and Table 3 shows where each method is used. The remaining 14, and what each needs beyond (*E, A, Y*):

- **Demographic parity, equalized odds, intersectional analysis**: a trained classifier’s predictions. Demographic parity is the difference in positive prediction rate between groups, equalized odds the largest difference in TPR or FPR, and intersectional analysis the same gaps over combinations of attributes. They are properties of a trained classifier; of the three, only the equalized-odds gaps (TPR and FPR) are reported in this paper, for the heads of Section 4.3.
- **GroupDRO** [66], **GCE** [67], **early-epoch clustering (SPARE)**: training a model and access to its loss or representations during training.
- **SSA**: a split into attribute-labelled and unlabelled data.
- **CAV**: user-defined concept examples. **SpRAy** [68] and **GradCAM mask overlap** [69]: images and model gradients.
- **Causal effect estimation** [70, 71], **generative CVAE, VAE disentanglement**: training an auxiliary model.
- **SIS**: the current implementation searches for sufficient subsets for the task label, with a fixed subset fraction of 15%. It has no attribute-aware form and was removed from the score.

## Appendix C. Mitigation method details

1. **Shortcut masking:** The embedding dimensions with the largest loading on the attribute direction (the top 5% here) are set to zero, randomised or inpainted. The method works on the embeddings themselves and needs no retraining.
2. **Background randomisation:** For image-level pipelines, foreground objects are swapped across backgrounds so that background and label are no longer correlated. It needs segmentation masks or a foreground extractor.
3. **Adversarial debiasing:** Adversarial training with a gradient reversal layer removes group information from the learned representation [72, 73]. An adversary tries to predict *A* from *E*, and the debiasing network is trained to raise the adversary’s loss while keeping task performance.
4. **Explanation regularisation (RRR):** A Right for the Right Reasons regularisation term, added to the training loss, penalises model attention on detected shortcut regions.
5. **Last-layer retraining (DFR):** Only the final classification layer is retrained, on a group-balanced subset of the data [54]. The method is cheap to run and suited to the case where the attribute is encoded in the representation and the head gives it too much weight. It leaves the embedding unchanged and, in Section 4.3, lowered the TPR gap at a cost in task AUROC.
6. **Contrastive debiasing:** A contrastive loss pulls together representations from different demographic groups within the same task class [74], so that the groups become less separable while the task classes stay apart.

Of these, shortcut masking, adversarial debiasing, last-layer retraining and contrastive debiasing work on frozen embeddings. They were run with the package defaults and are evaluated in Section 4.3. Background randomisation and explanation regularisation need images or model gradients and were not evaluated.

### Settings used in the head experiment

Adversarial debiasing uses a debiasing network (one linear layer to 64 units, with ReLU and dropout 0.1) followed by a linear task head and, behind a gradient-reversal layer (reversal weight 0.5), a linear adversary. The loss is the sum of the task and adversary cross-entropies, trained with Adam (learning rate 10^*−*3^, batch 64) for 50 epochs. Contrastive debiasing uses the same network. For each anchor it draws up to 32 positives (same *Y*, other *A*) and 32 negatives (other *Y*), and minimises 0.75 times an InfoNCE loss (temperature 0.05, one term per positive against the negatives) plus 0.25 times the task cross-entropy, with Adam (learning rate 10^*−*3^) for 50 epochs. Their controls are the same networks trained on the task loss alone. The contrastive loss in the released package was corrected during the revision: it had treated additional positives of an anchor as negatives. The embedding passed on is the 64-unit output of that network. Shortcut masking and its control are as defined in Section 3.6, applied to the training, test and audit embeddings.

DFR standardises the embedding and refits an L1 logistic head (*C* = 1, balanced class weights, and the L1 penalty follows the DFR reference implementation [54]) on a subset of the confounded training set with equal numbers of images in each of the four (*A, Y*) groups, so it uses 4*×* the size of the smallest group. To separate the effect of size from that of balancing, we refit the same L1 head on a random subset of the confounded training set of the same size, and an L2 head (as for the confounded head) on the balanced subset, over the same cells and seeds. Task AUROC was 0.65 for DFR, 0.65 for the random subset of the same size, which kept a TPR gap of 0.10 (so the small subset alone removed about half of the confounded head’s gap of 0.18), and 0.69 for the balanced subset with the L2 head, against 0.73 for the confounded head. Balancing itself cost no AUROC. The loss came from the small subset and the L1 penalty in about equal parts.

### Output shift

The output shift of Section 4.3 correlated negatively with the probe score (−0.26). Projecting out *u* moves each test embedding by its whole coordinate along *u*, most of which is spread within each attribute group rather than the gap between groups. The measure therefore tracks how much the head responds to that spread, which is largest for encoders that encode the attribute weakly, so it is reported in Table 8 but not used as a measure of reliance.

Table C.1 breaks the head-experiment results of Section 4.3 down by encoder.

**Table C.1:** Mitigation results per CheXpert encoder. Adversarial debiasing, contrastive debiasing and shortcut masking: change in the confounded head’s TPR gap and in task AUROC against the method’s control on the same cell and seed (method minus control). DFR: after minus before. Mean over the two attributes, three tasks and 3 seeds. The last row is the mean over all encoders, as in the text. A negative Δ TPR gap means a smaller gap.

|  | Adversarial |  | Contrastive |  | Masking |  | DFR |  |
|---|---|---|---|---|---|---|---|---|
| Encoder | Δ TPR gap | Δ AUROC | Δ TPR gap | Δ AUROC | Δ TPR gap | Δ AUROC | Δ TPR gap | Δ AUROC |
| ResNet-50 | 0.01 | 0.00 | 0.02 | 0.00 | -0.01 | 0.00 | -0.16 | -0.09 |
| DenseNet-121 | -0.01 | 0.00 | 0.06 | 0.00 | 0.00 | 0.00 | -0.23 | -0.07 |
| ViT-B/16 | 0.00 | 0.00 | 0.00 | 0.00 | 0.00 | 0.01 | -0.10 | -0.06 |
| ViT-B/32 | 0.03 | 0.00 | 0.02 | 0.00 | 0.00 | 0.01 | -0.09 | -0.07 |
| DINOv2 | -0.03 | 0.01 | -0.05 | 0.00 | 0.00 | 0.01 | -0.18 | -0.09 |
| RAD-DINO | -0.06 | 0.00 | -0.06 | -0.01 | -0.06 | 0.01 | -0.26 | -0.14 |
| MedSigLIP | -0.01 | 0.00 | -0.01 | 0.01 | 0.00 | 0.00 | -0.18 | -0.06 |
| All encoders | -0.01 | 0.00 | 0.00 | 0.00 | -0.01 | 0.00 | -0.17 | -0.08 |

### Race in the head experiment

The head experiment of Section 4.3 was repeated with race as the attribute, White against non-White as in the clinical audit, with images without a recorded race dropped and the same splits, seeds, logistic head and probe audit per cell. The mitigations were not rerun (Table C.2). Across the 21 encoder and task cells the Spearman correlation between the conditional probe AUROC and the TPR gap was 0.03 (interval −0.40 to 0.51 when whole encoders are resampled), the mean TPR gap was −0.01, and the probe AUROCs spanned 0.52 to 0.65. The race signal on CheXpert is weak for every encoder, below the lowest age cell of Table 8, and the confounded heads showed no consistent gap, so over this range the relation between audit and reliance was not detectable. Of the seed draws, 7 were skipped because the smallest (*A, Y*) group of the training half did not allow *N ≥* 40 at *ρ*_train_ = 0.9.

**Table C.2:** Classification-head experiment with race as the attribute (White against non-White, images without a recorded race dropped), one row per CheXpert encoder and task, 10 seeds. Head, splits and test set as in Table 8. Probe: conditional probe AUROC and permutation *p* on all labelled images of the task with a recorded race. *n* train: size of the confounded training set over the feasible seeds. TPR gap between groups on the balanced test set, mean (SD over seeds). AUROC drop versus an unconfounded head of equal training size.

| Encoder | *Y* | *n* train (min-max) | Probe | *p* | TPR gap (SD) | AUROC drop |
|---|---|---|---|---|---|---|
| ResNet-50 | Pneumothorax | 48-106 | 0.58 | 0.02 | 0.05 (0.13) | 0.05 |
| ResNet-50 | Pleural Effusion | 124-252 | 0.59 | <0.01 | 0.05 (0.05) | 0.02 |
| ResNet-50 | Cardiomegaly | 62-92 | 0.57 | 0.07 | 0.03 (0.13) | 0.05 |
| DenseNet-121 | Pneumothorax | 48-106 | 0.61 | <0.01 | -0.11 (0.09) | 0.06 |
| DenseNet-121 | Pleural Effusion | 124-252 | 0.60 | <0.01 | 0.02 (0.12) | 0.01 |
| DenseNet-121 | Cardiomegaly | 62-92 | 0.61 | 0.02 | 0.00 (0.12) | 0.04 |
| ViT-B/16 | Pneumothorax | 48-106 | 0.65 | <0.01 | -0.10 (0.19) | 0.04 |
| ViT-B/16 | Pleural Effusion | 124-252 | 0.58 | 0.01 | -0.03 (0.12) | 0.03 |
| ViT-B/16 | Cardiomegaly | 62-92 | 0.54 | 0.17 | 0.00 (0.11) | 0.00 |
| ViT-B/32 | Pneumothorax | 48-106 | 0.61 | <0.01 | -0.02 (0.18) | 0.04 |
| ViT-B/32 | Pleural Effusion | 124-252 | 0.56 | 0.04 | -0.05 (0.06) | 0.01 |
| ViT-B/32 | Cardiomegaly | 62-92 | 0.56 | 0.08 | 0.00 (0.12) | 0.05 |
| DINOv2 | Pneumothorax | 48-106 | 0.61 | <0.01 | 0.01 (0.23) | 0.01 |
| DINOv2 | Pleural Effusion | 124-252 | 0.59 | <0.01 | -0.03 (0.08) | 0.02 |
| DINOv2 | Cardiomegaly | 62-92 | 0.57 | 0.07 | -0.07 (0.17) | -0.01 |
| RAD-DINO | Pneumothorax | 48-106 | 0.61 | 0.02 | 0.16 (0.08) | 0.04 |
| RAD-DINO | Pleural Effusion | 124-252 | 0.56 | 0.06 | -0.04 (0.12) | 0.02 |
| RAD-DINO | Cardiomegaly | 62-92 | 0.58 | 0.07 | 0.07 (0.16) | 0.03 |
| MedSigLIP | Pneumothorax | 48-106 | 0.57 | 0.05 | -0.05 (0.18) | 0.03 |
| MedSigLIP | Pleural Effusion | 124-252 | 0.52 | 0.34 | -0.07 (0.07) | 0.01 |
| MedSigLIP | Cardiomegaly | 62-92 | 0.52 | 0.34 | 0.07 (0.12) | 0.05 |

## Appendix D. Implementation and reproducibility

Seeds, permutation counts, run times and the configuration of every stage are documented in the repository, with the run manifests from which each table and figure was regenerated.

## Appendix E. Synthetic benchmark detail

The generator draws *y*_*i*_ *~* Bernoulli(0.5) and sets *a*_*i*_ = *y*_*i*_ with probability *ρ* and *a*_*i*_ = 1 *− y*_*i*_ otherwise. The embedding has *d* coordinates drawn from *N* (0, 1). A block of *k*_*y*_ = 4 task dimensions is shifted by *µ*_*y*_/2 according to *y*_*i*_, with *± µ*_*y*_ = 1.0. A disjoint block of *k*_*a*_ = 4 attribute dimensions carries the attribute signal:

- **Linear**: each attribute dimension is shifted by +*δ*/2 for *a*_*i*_ = 1 and *−δ*/2 for, *a*_*i*_ = 0 so the standardised mean difference is exactly *δ* in each dimension and the Mahalanobis distance over the block is 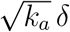.
- **Nonlinear (XOR)**: each pair of attribute dimensions is shifted by (*s*_1_, *s*_2_) *δ*/2, where *s*_1_ is a random sign drawn per image, independently for each pair, and *s*_2_ = *−s*_1_ for *a*_*i*_ = 1 and *s*_2_ = *s*_1_ for *a*_*i*_ = 0. Both groups have zero mean in every dimension. Given the partner coordinate, they differ by *δ* standard deviations.
- **Variance shift**: attribute dimensions have standard deviation 1 for *a*_*i*_ = 0 and 1 + *δ* for *a*_*i*_ = 1, with equal means.

In the submitted version the generator passed the task label as the group, so the synthetic benchmark measured task signal, and the shift was *±δ*, which made the effect 2*δ*. Both are corrected here. Figure E.1 shows the perdimension false positive rate of the statistical detector on null data with and without multiple testing correction.

**Figure E.1.**
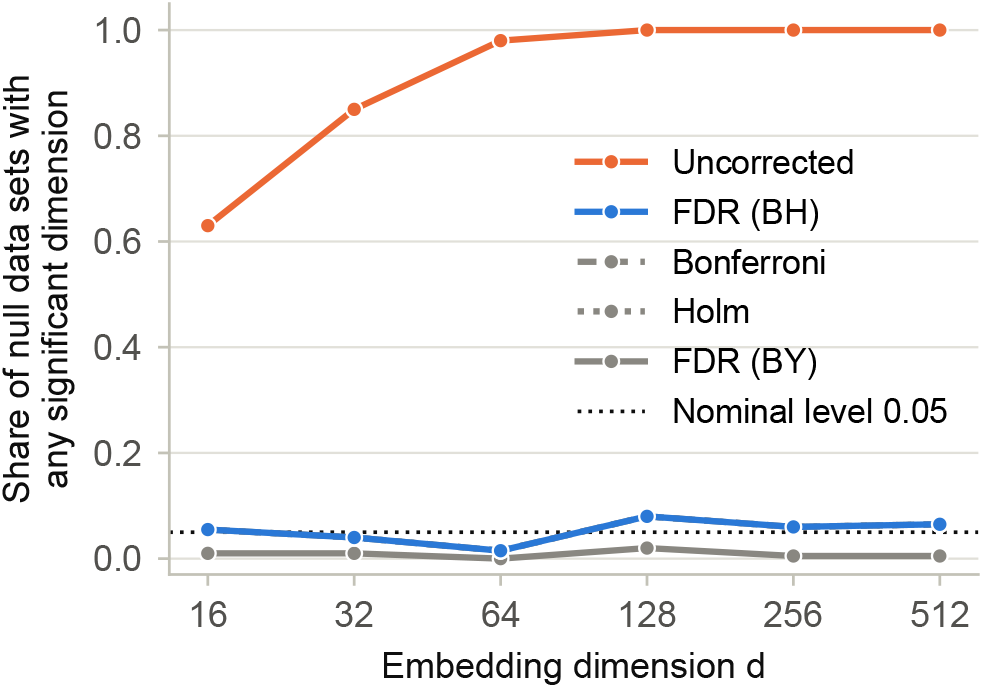
Share of synthetic null data sets (*δ* = 0) in which the statistical detector found at least one significant dimension, against embedding dimension, without correction and with Benjamini–Hochberg, Bonferroni, Holm and Benjamini–Yekutieli correction, with *n* = 1000 and 10 seeds. The Bonferroni, Holm and Benjamini–Yekutieli curves coincide.

**Table E.2:** Correlation of detector flags across all synthetic runs (linear variant).

|  | Probe | Stat. | Geom. | BiasDir | HBAC-NMI | Freq. |
|---|---|---|---|---|---|---|
| Probe | 1.0 | 0.86 | 0.9 | 0.8 | 0.57 | 0.91 |
| Stat. | 0.86 | 1.0 | 0.87 | 0.77 | 0.58 | 0.87 |
| Geom. | 0.9 | 0.87 | 1.0 | 0.88 | 0.6 | 0.89 |
| BiasDir | 0.8 | 0.77 | 0.88 | 1.0 | 0.58 | 0.8 |
| HBAC-NMI | 0.57 | 0.58 | 0.6 | 0.58 | 1.0 | 0.54 |
| Freq. | 0.91 | 0.87 | 0.89 | 0.8 | 0.54 | 1.0 |

**Table E.1:**
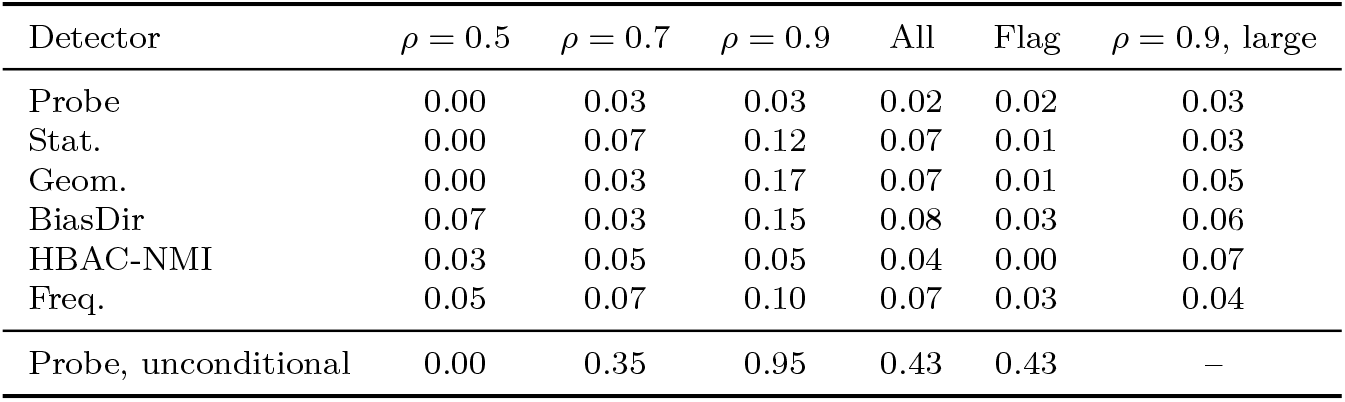
Synthetic null (*δ* = 0): rejection rate per detector at *p <* 0.05 before correction, by *ρ* and pooled (All), and flag rate under the decision rule (Flag: probe at *p <* 0.05, secondary detectors at Benjamini–Hochberg *q <* 0.05 across the five), pooled over *n, d* and the three variants, 10 seeds. Last row: an unconditional probe (predicting *A* from *E* alone, *A* permuted without stratification). Last column: 500 further null data sets at *ρ* = 0.9, *n* = 1000 and *d* = 128 (Section 5.1); –: not run.

| Detector | ρ = 0.5 | ρ = 0.7 | ρ = 0.9 | All | Flag | ρ = 0.9, large |
|---|---|---|---|---|---|---|
| Probe | 0.00 | 0.03 | 0.03 | 0.02 | 0.02 | 0.03 |
| Stat. | 0.00 | 0.07 | 0.12 | 0.07 | 0.01 | 0.03 |
| Geom. | 0.00 | 0.03 | 0.17 | 0.07 | 0.01 | 0.05 |
| BiasDir | 0.07 | 0.03 | 0.15 | 0.08 | 0.03 | 0.06 |
| HBAC-NMI | 0.03 | 0.05 | 0.05 | 0.04 | 0.00 | 0.07 |
| Freq. | 0.05 | 0.07 | 0.10 | 0.07 | 0.03 | 0.04 |
| Probe, unconditional | 0.00 | 0.35 | 0.95 | 0.43 | 0.43 | – |

### Nonlinear probes

The blind spot named in Section 7 was checked directly at one small setting of the benchmark (*n* = 1000, *d* = 64, *ρ* = 0.5, *K* = 200, 50 seeds), without rerunning the grid. The conditional probe was fitted three times on each data set, with the same code and in-fold standardisation, the same folds and the same null (*A* permuted within *Y*). Only the design changed: *E* (the audit’s probe), [*E, E*^2^] with the squared coordinates appended, and [*E, E*^2^, *E*_*i*_*E*_*j*_] with all *d*(*d −* 1)/2 pairwise products appended as well (2144 features). Table E.3 gives the rejection rate at *p <* 0.05 and the mean AUROC. At *δ* = 2.0 the squared-coordinate probe rejected the variance shift in 1.00 of runs and the product probe rejected the XOR signal in 1.00 of runs (0.52 at *δ* = 1.2), while the linear probe stayed at 0.06 on XOR and 0.06 on the variance shift. On null data the three probes rejected in 0.06, 0.04 and 0.06 of runs, so the permutation null calibrates the nonlinear designs as it does the linear one.

**Table E.3:** Conditional probe on linear and nonlinear features of the synthetic embeddings, all three under the same permutation null (*A* permuted within *Y*): rejection rate at *p <* 0.05 (Rej.) and mean AUROC over 50 seeds, at *n* = 1000, *d* = 64, *ρ* = 0.5, *K* = 200. [*E*] is the audit’s probe, [*E, E*^2^] appends the squared coordinates, and [*E, E*^2^, *E*_*i*_*E*_*j*_] also appends all pairwise products. The null row is the same data for the three variants (*δ* = 0).

|  |  | [*E*] |  | [*E*, *E*<sup>2</sup>] |  | [*E*, *E*<sup>2</sup>, *E*<sub>*i*</sub>*E*<sub>*j*</sub>] |  |
|---|---|---|---|---|---|---|---|
| Variant | *δ* | Rej. | AUROC | Rej. | AUROC | Rej. | AUROC |
| Null | 0 | 0.06 | 0.50 | 0.04 | 0.50 | 0.06 | 0.50 |
| Linear | 1.2 | 1.00 | 0.94 | 1.00 | 0.92 | 1.00 | 0.78 |
| Linear | 2 | 1.00 | 0.99 | 1.00 | 0.99 | 1.00 | 0.93 |
| XOR | 1.2 | 0.06 | 0.50 | 0.06 | 0.50 | 0.52 | 0.54 |
| XOR | 2 | 0.06 | 0.50 | 0.04 | 0.50 | 1.00 | 0.65 |
| Variance | 1.2 | 0.06 | 0.50 | 1.00 | 0.88 | 1.00 | 0.65 |
| Variance | 2 | 0.06 | 0.50 | 1.00 | 0.94 | 1.00 | 0.69 |

### Squared-feature probe on the clinical cohorts

The [*E, E*^2^] probe was then fitted on every primary cell of the clinical audit (21 CheXpert and 12 MIMIC-CXR cells) with the audit’s seed and patient-grouped folds, and with *K* = 500 permutation draws. The product design was not run, since its *d*(*d −* 1)/2 columns are too many at the encoders’ dimensions. Table E.4 lists the result beside the linear probe: the squared probe flagged 21 of 21 CheXpert cells and 12 of 12 MIMIC-CXR cells, and the decision changed in 0 cells against the linear probe. The largest AUROC gain over the linear probe was 0.02, and no AUROC moved by more than 0.03 in either direction, so on these cohorts the squared coordinates carry no attribute signal the linear probe had missed.

**Table E.4:** Conditional probe on [*E*] (the audit’s probe) and on [*E, E*^2^] for the primary clinical cells, under the audit’s permutation null (*A* permuted within *Y* between patients, *K* = 500) and patient-grouped folds. *p* and the flag (*p <* 0.05) are those of the squared probe. The linear probe’s decision is given in parentheses where it differs.

| Dataset | Encoder | Attribute | AUROC [*E*] | AUROC [*E*, *E*<sup>2</sup>] | *p* | Flag |
|---|---|---|---|---|---|---|
| CheXpert | ResNet-50 | Sex | 0.84 | 0.82 | <0.01 | yes |
|  |  | Age 60+ | 0.75 | 0.74 | <0.01 | yes |
|  |  | White vs non-White | 0.59 | 0.60 | <0.01 | yes |
|  | DenseNet-121 | Sex | 0.88 | 0.88 | <0.01 | yes |
|  |  | Age 60+ | 0.70 | 0.71 | <0.01 | yes |
|  |  | White vs non-White | 0.60 | 0.61 | <0.01 | yes |
|  | ViT-B/16 | Sex | 0.85 | 0.84 | <0.01 | yes |
|  |  | Age 60+ | 0.70 | 0.71 | <0.01 | yes |
|  |  | White vs non-White | 0.60 | 0.59 | <0.01 | yes |
|  | ViT-B/32 | Sex | 0.87 | 0.87 | <0.01 | yes |
|  |  | Age 60+ | 0.67 | 0.65 | <0.01 | yes |
|  |  | White vs non-White | 0.60 | 0.61 | <0.01 | yes |
|  | DINOv2 | Sex | 0.89 | 0.89 | <0.01 | yes |
|  |  | Age 60+ | 0.75 | 0.74 | <0.01 | yes |
|  |  | White vs non-White | 0.59 | 0.58 | <0.01 | yes |
|  | RAD-DINO | Sex | 0.99 | 0.99 | <0.01 | yes |
|  |  | Age 60+ | 0.83 | 0.80 | <0.01 | yes |
|  |  | White vs non-White | 0.61 | 0.61 | <0.01 | yes |
|  | MedSigLIP | Sex | 0.97 | 0.97 | <0.01 | yes |
|  |  | Age 60+ | 0.80 | 0.79 | <0.01 | yes |
|  |  | White vs non-White | 0.57 | 0.56 | 0.046 | yes |
| MIMIC-CXR | RAD-DINO | Sex | 1.00 | 1.00 | <0.01 | yes |
|  |  | Age 60+ | 0.89 | 0.89 | <0.01 | yes |
|  |  | White vs non-White | 0.76 | 0.76 | <0.01 | yes |
|  | ViT-B/16 | Sex | 0.93 | 0.93 | <0.01 | yes |
|  |  | Age 60+ | 0.80 | 0.80 | <0.01 | yes |
|  |  | White vs non-White | 0.73 | 0.72 | <0.01 | yes |
|  | ViT-B/32 | Sex | 0.94 | 0.94 | <0.01 | yes |
|  |  | Age 60+ | 0.83 | 0.82 | <0.01 | yes |
|  |  | White vs non-White | 0.66 | 0.67 | <0.01 | yes |
|  | MedSigLIP | Sex | 0.99 | 0.99 | <0.01 | yes |
|  |  | Age 60+ | 0.89 | 0.89 | <0.01 | yes |
|  |  | White vs non-White | 0.71 | 0.71 | <0.01 | yes |

## Appendix F. Statistical detail for the clinical cohorts

Cohort composition is in Table F.4, and Table F.3 is the fake-attribute control that motivates patient-level calibration. Two integrity checks follow: the row-alignment check (Table F.5) and the re-extraction of the CheXpert encoders whose code had not been kept (Table F.6). The sensitivity analyses for race, age and group size are Tables F.1, F.2 and F.8, with the comparison of clusters against metadata in Table F.9. The raw statistics of every detector in every cell are in Tables F.11 to F.16, and Table F.10 reruns the submitted pipeline with its flag reader fixed. The CheXpert sex labels in the submitted runs were inverted (the mirror codes 0 as male). All results here use the sex recorded in the official `train.csv`.

**Table F.1:** Race sensitivity analysis: conditional probe AUROC and permutation *p* for White against non-White, for the race groups with at least 50 images kept separate (White, Black and Asian on CheXpert, White and Black on MIMIC-CXR), and for the same groups restricted to at least 50 images in every task stratum (on CheXpert only White patients reach 50 images with *Y* = 1, so there the last column compares groups among *Y* = 0 images only). One row per cohort and frozen encoder, seed 42.

| Dataset | Encoder | White vs non-White | Groups with 50+ (W/B/A) | Groups with 50+ per *Y* |
|---|---|---|---|---|
| CheXpert | ResNet-50 | 0.59 (*p* <0.01) | 0.63 (*p* <0.01) | 0.63 (*p* <0.01) |
| CheXpert | MedSigLIP | 0.57 (*p* 0.02) | 0.62 (*p* <0.01) | 0.67 (*p* <0.01) |
| CheXpert | DenseNet-121 | 0.60 (*p* <0.01) | 0.66 (*p* <0.01) | 0.68 (*p* <0.01) |
| CheXpert | ViT-B/16 | 0.60 (*p* <0.01) | 0.65 (*p* <0.01) | 0.67 (*p* <0.01) |
| CheXpert | ViT-B/32 | 0.60 (*p* <0.01) | 0.63 (*p* <0.01) | 0.64 (*p* <0.01) |
| CheXpert | DINOv2 | 0.59 (*p* <0.01) | 0.63 (*p* <0.01) | 0.66 (*p* <0.01) |
| CheXpert | RAD-DINO | 0.61 (*p* <0.01) | 0.63 (*p* <0.01) | 0.64 (*p* <0.01) |
| MIMIC-CXR | MedSigLIP | 0.71 (*p* <0.01) | 0.77 (*p* <0.01) | 0.78 (*p* <0.01) |
| MIMIC-CXR | RAD-DINO | 0.76 (*p* <0.01) | 0.87 (*p* <0.01) | 0.87 (*p* <0.01) |
| MIMIC-CXR | ViT-B/16 | 0.73 (*p* <0.01) | 0.78 (*p* <0.01) | 0.78 (*p* <0.01) |
| MIMIC-CXR | ViT-B/32 | 0.66 (*p* <0.01) | 0.74 (*p* <0.01) | 0.75 (*p* <0.01) |

**Table F.2:** Age sensitivity analysis: conditional probe AUROC and permutation *p* for age 60 or over against under 60, and for four age bins (under 40, 40 to 59, 60 to 79, 80 and over, with macro one-vs-rest AUROC). One row per cohort and frozen encoder, seed 42.

| Dataset | Encoder | Age 60+ (binary) | Age, 4 bins |
|---|---|---|---|
| CheXpert | ResNet-50 | 0.75 (*p* <0.01) | 0.65 (*p* <0.01) |
| CheXpert | MedSigLIP | 0.80 (*p* <0.01) | 0.70 (*p* <0.01) |
| CheXpert | DenseNet-121 | 0.70 (*p* <0.01) | 0.66 (*p* <0.01) |
| CheXpert | ViT-B/16 | 0.70 (*p* <0.01) | 0.66 (*p* <0.01) |
| CheXpert | ViT-B/32 | 0.67 (*p* <0.01) | 0.63 (*p* <0.01) |
| CheXpert | DINOv2 | 0.75 (*p* <0.01) | 0.66 (*p* <0.01) |
| CheXpert | RAD-DINO | 0.83 (*p* <0.01) | 0.76 (*p* <0.01) |
| MIMIC-CXR | MedSigLIP | 0.89 (*p* <0.01) | 0.80 (*p* <0.01) |
| MIMIC-CXR | RAD-DINO | 0.89 (*p* <0.01) | 0.79 (*p* <0.01) |
| MIMIC-CXR | ViT-B/16 | 0.80 (*p* <0.01) | 0.72 (*p* <0.01) |
| MIMIC-CXR | ViT-B/32 | 0.83 (*p* <0.01) | 0.73 (*p* <0.01) |

**Table F.3:** Fake-attribute negative control, CheXpert RAD-DINO: fraction of 30 draws of a random patient-level binary attribute in which each detector rejected at *p <* 0.05, with patient-level permutation and folds grouped by patient, and with image-level permutation and ordinary folds.

| Detector | Patient-level permutation, grouped CV | Image-level permutation, plain CV |
|---|---|---|
| Probe | 0.00 | 1.00 |
| Stat. | 0.00 | 1.00 |
| Geom. | 0.07 | 1.00 |
| BiasDir | 0.00 | 1.00 |
| HBAC-NMI | 0.03 | 1.00 |
| Freq. | 0.03 | 1.00 |

**Table F.4:** Cohort composition: images, patients, sex, age, race groups and audit task for CheXpert (first 2,000 images of the official training file) and MIMIC-CXR (one image per subject).

| Cohort | Images | Patients | Male | Age 60+ | White | Black | Asian | NHPI | Other | Missing | Task (*Y*) | *Y*=1 |
|---|---|---|---|---|---|---|---|---|---|---|---|---|
| CheXpert | 2000 | 477 | 1149 (57%) | 961 (48%) | 1150 | 140 | 217 | 24 | 252 | 217 | Pneumothorax | 218 (10.9%) |
| MIMIC-CXR | 1491 | 1491 | 724 (49%) | 877 (59%) | 1027 | 208 | 38 | 0 | 147 | 71 | In-hospital mortality | 90 (6.0%) |

**Table F.5:** Row-alignment check: largest Spearman correlation between the pairwise cosine-similarity structure of one encoder’s embeddings and that of the other encoders on the same images (the first 400 rows of each file), against a control with rows shuffled. Low agreement with every other encoder means the rows do not match the metadata. The row “stored file, replaced” is the submitted MIMIC-CXR ViT-B/32 file, and the ViT-B/32 row above it is the file rebuilt from the source and used in every result.

| Dataset | Encoder | Max Spearman | Shuffled control | Sex probe AUROC | Similarity fails | Probe at chance | Replaced |
|---|---|---|---|---|---|---|---|
| CheXpert | DenseNet-121 | 0.780 | -0.001 | 0.88 | no | no | no |
| CheXpert | DINOv2 | 0.701 | 0.004 | 0.89 | no | no | no |
| CheXpert | MedSigLIP | 0.608 | -0.003 | 0.97 | no | no | no |
| CheXpert | RAD-DINO | 0.311 | -0.003 | 0.99 | no | no | no |
| CheXpert | ResNet-50 | 0.780 | -0.002 | 0.84 | no | no | no |
| CheXpert | ViT-B/16 | 0.753 | -0.001 | 0.85 | no | no | no |
| CheXpert | ViT-B/32 | 0.662 | 0.001 | 0.87 | no | no | no |
| MIMIC-CXR | MedSigLIP | 0.414 | -0.002 | 0.99 | no | no | no |
| MIMIC-CXR | RAD-DINO | 0.387 | -0.002 | 1.00 | no | no | no |
| MIMIC-CXR | ViT-B/16 | 0.499 | 0.003 | 0.93 | no | no | no |
| MIMIC-CXR | ViT-B/32 | 0.499 | -0.004 | 0.94 | no | no | no |
| MIMIC-CXR | ViT-B/32 (stored file, replaced) | 0.006 | 0.000 | 0.50 | yes | yes | yes |

**Table F.6:**
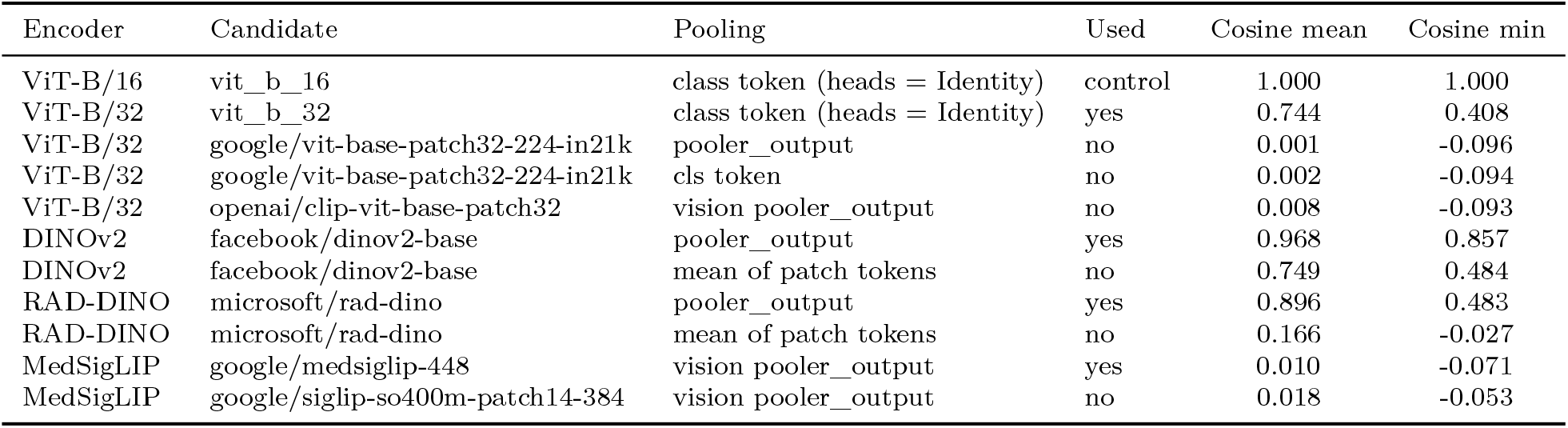
Re-extraction of the four CheXpert encoders whose arrays in the first submission had no extraction code: candidate public checkpoint and pooling, and cosine similarity between re-extracted and stored embeddings of the same images (mean and minimum over images). Used: the re-extraction that all CheXpert results in this paper use. ViT-B/16, whose extraction code was kept, is re-extracted with the same code as a control. The stored MedSigLIP array matched neither google/medsiglip-448 nor SigLIP So400m, so its source is unknown.

| Encoder | Candidate | Pooling | Used | Cosine mean | Cosine min |
|---|---|---|---|---|---|
| ViT-B/16 | vit_b_16 | class token (heads = Identity) | control | 1.000 | 1.000 |
| ViT-B/32 | vit_b_32 | class token (heads = Identity) | yes | 0.744 | 0.408 |
| ViT-B/32 | google/vit-base-patch32-224-in21k | pooler_output | no | 0.001 | -0.096 |
| ViT-B/32 | google/vit-base-patch32-224-in21k | cls token | no | 0.002 | -0.094 |
| ViT-B/32 | openai/clip-vit-base-patch32 | vision pooler_output | no | 0.008 | -0.093 |
| DINOv2 | facebook/dinov2-base | pooler_output | yes | 0.968 | 0.857 |
| DINOv2 | facebook/dinov2-base | mean of patch tokens | no | 0.749 | 0.484 |
| RAD-DINO | microsoft/rad-dino | pooler_output | yes | 0.896 | 0.483 |
| RAD-DINO | microsoft/rad-dino | mean of patch tokens | no | 0.166 | -0.027 |
| MedSigLIP | google/medsiglip-448 | vision pooler_output | yes | 0.010 | -0.071 |
| MedSigLIP | google/siglip-so400m-patch14-384 | vision pooler_output | no | 0.018 | -0.053 |

**Table F.7:** Sensitivity of the CheXpert conditional probe AUROC (task Pneumothorax). Main: the audit reported in the paper. Stored array: the arrays used in the first sub-mission, for the four encoders that were re-extracted (same: unchanged encoder). Frontal only: the 1577 frontal images. Labelled only: the 802 images with a definite Pneumothorax label (uncertain and blank dropped, as in the head experiment). *C*: the probe’s regularisation, 1 in the main audit. *: probe *p <* 0.05, the decision rule of the main audit, in every column.

| Encoder | *A* | Main | Stored array | Frontal only | Labelled only | *C* = 0.1 | *C* = 10 |
|---|---|---|---|---|---|---|---|
| ResNet-50 | Sex | 0.84* | same | 0.85* | 0.82* | 0.84* | 0.84* |
| ResNet-50 | Age | 0.75* | same | 0.74* | 0.76* | 0.75* | 0.74* |
| ResNet-50 | Race | 0.59* | same | 0.59* | 0.58* | 0.60* | 0.59* |
| MedSigLIP | Sex | 0.97* | 0.68* | 0.97* | 0.98* | 0.97* | 0.97* |
| MedSigLIP | Age | 0.80* | 0.57* | 0.81* | 0.80* | 0.81* | 0.79* |
| MedSigLIP | Race | 0.57* | 0.53 | 0.57* | 0.57 | 0.57* | 0.56* |
| DenseNet-121 | Sex | 0.88* | same | 0.90* | 0.89* | 0.89* | 0.88* |
| DenseNet-121 | Age | 0.70* | same | 0.72* | 0.75* | 0.72* | 0.69* |
| DenseNet-121 | Race | 0.60* | same | 0.62* | 0.61* | 0.61* | 0.59* |
| ViT-B/16 | Sex | 0.85* | same | 0.86* | 0.86* | 0.87* | 0.85* |
| ViT-B/16 | Age | 0.70* | same | 0.71* | 0.72* | 0.73* | 0.69* |
| ViT-B/16 | Race | 0.60* | same | 0.62* | 0.65* | 0.63* | 0.60* |
| ViT-B/32 | Sex | 0.87* | 0.88* | 0.88* | 0.85* | 0.89* | 0.87* |
| ViT-B/32 | Age | 0.67* | 0.68* | 0.66* | 0.67* | 0.69* | 0.66* |
| ViT-B/32 | Race | 0.60* | 0.59* | 0.58* | 0.61* | 0.63* | 0.59* |
| DINOv2 | Sex | 0.89* | 0.92* | 0.92* | 0.84* | 0.90* | 0.88* |
| DINOv2 | Age | 0.75* | 0.75* | 0.75* | 0.75* | 0.76* | 0.74* |
| DINOv2 | Race | 0.59* | 0.61* | 0.61* | 0.61* | 0.62* | 0.59* |
| RAD-DINO | Sex | 0.99* | 0.99* | 1.00* | 0.99* | 0.99* | 0.99* |
| RAD-DINO | Age | 0.83* | 0.80* | 0.84* | 0.83* | 0.84* | 0.82* |
| RAD-DINO | Race | 0.61* | 0.62* | 0.62* | 0.61* | 0.62* | 0.61* |

**Table F.8:** Group-size stability of the race result, RAD-DINO: the non-White group sub-sampled to a target size *m*, ten draws per size, the White group kept whole. Conditional probe AUROC (mean and SD) and the number of draws with *p <* 0.05. MIMIC-CXR has 393 non-White images with a recorded race, so at *m* = 400 every draw is the whole group and the SD is zero. On CheXpert whole patients are drawn until at least *m* images are reached, so the counts exceed *m* slightly.

| Dataset | Target *m* | Non-White images | Non-White *Y*=1 | Probe AUROC (mean ± SD) | *p* < 0.05 |
|---|---|---|---|---|---|
| CheXpert | 25 | 31 | 5.5 | 0.48 ± 0.16 | 1/10 |
| CheXpert | 50 | 52 | 3.6 | 0.53 ± 0.08 | 2/10 |
| CheXpert | 100 | 111 | 12.9 | 0.56 ± 0.04 | 0/10 |
| CheXpert | 200 | 203 | 21.3 | 0.60 ± 0.03 | 7/10 |
| CheXpert | 400 | 406 | 42.3 | 0.58 ± 0.02 | 8/10 |
| MIMIC-CXR | 25 | 25 | 0.8 | 0.70 ± 0.05 | 8/10 |
| MIMIC-CXR | 50 | 50 | 1.4 | 0.71 ± 0.06 | 10/10 |
| MIMIC-CXR | 100 | 100 | 3.5 | 0.74 ± 0.03 | 10/10 |
| MIMIC-CXR | 200 | 200 | 6.2 | 0.75 ± 0.02 | 10/10 |
| MIMIC-CXR | 400 | 393 | 12.0 | 0.76 ± 0.00 | 10/10 |

**Table F.9:** Association between *k*-means clusters (*k* = 8) of the embeddings and recorded metadata: normalised mutual information (mean and range over encoders), Cramér’s *V*, and the number of encoders with a *χ*^2^ test at *p <* 0.05.

| Dataset | Variable | NMI (mean) | NMI (range over encoders) | Cramér's V (mean) | *χ*<sup>2</sup> *p* < 0.05 |
|---|---|---|---|---|---|
| CheXpert | view (frontal/lateral) | 0.378 | 0.260 to 0.431 | 0.96 | 7/7 |
| CheXpert | AP/PA | 0.484 | 0.416 to 0.550 | 0.87 | 7/7 |
| CheXpert | Support Devices | 0.068 | 0.052 to 0.103 | 0.41 | 7/7 |
| CheXpert | sex | 0.015 | 0.005 to 0.050 | 0.18 | 7/7 |
| CheXpert | age 60+ | 0.019 | 0.009 to 0.036 | 0.21 | 7/7 |
| CheXpert | race White | 0.013 | 0.009 to 0.022 | 0.13 | 7/7 |
| CheXpert | Pneumothorax | 0.019 | 0.010 to 0.032 | 0.21 | 7/7 |
| CheXpert | Pleural Effusion | 0.034 | 0.020 to 0.078 | 0.29 | 7/7 |
| CheXpert | No Finding | 0.029 | 0.021 to 0.044 | 0.26 | 7/7 |
| MIMIC-CXR | view (AP/PA) | 0.232 | 0.141 to 0.399 | 0.70 | 4/4 |
| MIMIC-CXR | sex | 0.027 | 0.006 to 0.043 | 0.25 | 4/4 |
| MIMIC-CXR | age 60+ | 0.051 | 0.030 to 0.071 | 0.36 | 4/4 |
| MIMIC-CXR | race White | 0.007 | 0.002 to 0.012 | 0.13 | 3/4 |
| MIMIC-CXR | mortality | 0.037 | 0.026 to 0.055 | 0.29 | 4/4 |

**Table F.10:** The submitted 13-method pipeline rerun with only the flag reader fixed: number of cells flagged by each method as read in the submitted tables and as read correctly, and the number of cells in which the method ran. SSA failed in every run. This table documents the old pipeline, and its outputs are not used elsewhere.

| Dataset | Method | Submitted reading | Fixed reading | Evaluated |
|---|---|---|---|---|
| CheXpert | HBAC | 21/21 | 21/21 | 21/21 |
|  | Probe | 6/21 | 6/21 | 21/21 |
|  | Stat. | 0/21 | 21/21 | 21/21 |
|  | Geom. | 0/21 | 21/21 | 21/21 |
|  | Freq. | 21/21 | 21/21 | 21/21 |
|  | BiasDir | 21/21 | 21/21 | 21/21 |
|  | SIS | 21/21 | 21/21 | 21/21 |
|  | DP | 21/21 | 14/21 | 21/21 |
|  | EO | 15/21 | 9/21 | 21/21 |
|  | Inter. | 21/21 | 21/21 | 21/21 |
|  | GDRO | 0/21 | 0/21 | 21/21 |
|  | GCE | 0/21 | 21/21 | 21/21 |
|  | SSA | 0/21 | n/a | 0/21 |
| MIMIC-CXR | HBAC | 12/12 | 12/12 | 12/12 |
|  | Probe | 3/12 | 3/12 | 12/12 |
|  | Stat. | 0/12 | 9/12 | 12/12 |
|  | Geom. | 0/12 | 12/12 | 12/12 |
|  | Freq. | 9/12 | 9/12 | 12/12 |
|  | BiasDir | 12/12 | 12/12 | 12/12 |
|  | SIS | 12/12 | 12/12 | 12/12 |
|  | DP | 9/12 | 3/12 | 12/12 |
|  | EO | 3/12 | 3/12 | 12/12 |
|  | Inter. | 3/12 | 3/12 | 12/12 |
|  | GDRO | 0/12 | 0/12 | 12/12 |
|  | GCE | 0/12 | 12/12 | 12/12 |
|  | SSA | 0/12 | n/a | 0/12 |
| CelebA | HBAC | 0/3 | 0/3 | 3/3 |
|  | Probe | 3/3 | 3/3 | 3/3 |
|  | Stat. | 0/3 | 3/3 | 3/3 |
|  | Geom. | 0/3 | 3/3 | 3/3 |
|  | Freq. | 3/3 | 3/3 | 3/3 |
|  | BiasDir | 3/3 | 3/3 | 3/3 |
|  | SIS | 3/3 | 3/3 | 3/3 |
|  | DP | 3/3 | 3/3 | 3/3 |
|  | EO | 3/3 | 3/3 | 3/3 |
|  | Inter. | 3/3 | 3/3 | 3/3 |
|  | GDRO | 0/3 | 0/3 | 3/3 |
|  | GCE | 0/3 | 3/3 | 3/3 |
|  | SSA | 0/3 | n/a | 0/3 |

**Table F.11:** Raw statistics of the six detectors in every CheXpert cell, attribute sex (task Pneumothorax, primary audit, seed 42): statistic *s* as in Table 6, mean and SD of its conditional permutation null, raw permutation *p*-value, and, for the five secondary detectors, the Benjamini–Hochberg *q*-value across those five (the probe is judged on its raw *p*, so its *q* column shows –). The smallest attainable *p* is 1/(*K* + 1), with *K* = 500 permutations for the probe and 200 for the other detectors.

| Encoder | Detector | *s* | Null mean | Null SD | *p* | *q* |
|---|---|---|---|---|---|---|
| ResNet-50 | Probe | 0.84 | 0.496 | 0.024 | 0.002 | – |
|  | Stat. | 0.300 | 0.063 | 0.063 | 0.005 | 0.006 |
|  | Geom. | 0.15 | 0.093 | 0.018 | 0.005 | 0.006 |
|  | BiasDir | 1.01 | 0.430 | 0.066 | 0.005 | 0.006 |
|  | HBAC-NMI | 0.008 | 0.007 | 0.003 | 0.274 | 0.274 |
|  | Freq. | 0.80 | 0.493 | 0.031 | 0.005 | 0.006 |
| DenseNet-121 | Probe | 0.88 | 0.496 | 0.027 | 0.002 | – |
|  | Stat. | 0.551 | 0.202 | 0.124 | 0.005 | 0.006 |
|  | Geom. | 0.16 | 0.094 | 0.017 | 0.005 | 0.006 |
|  | BiasDir | 1.16 | 0.420 | 0.057 | 0.005 | 0.006 |
|  | HBAC-NMI | 0.014 | 0.013 | 0.004 | 0.428 | 0.428 |
|  | Freq. | 0.85 | 0.490 | 0.036 | 0.005 | 0.006 |
| ViT-B/16 | Probe | 0.85 | 0.495 | 0.024 | 0.002 | – |
|  | Stat. | 0.462 | 0.158 | 0.112 | 0.010 | 0.012 |
|  | Geom. | 0.14 | 0.092 | 0.015 | 0.010 | 0.012 |
|  | BiasDir | 0.99 | 0.450 | 0.068 | 0.005 | 0.012 |
|  | HBAC-NMI | 0.011 | 0.009 | 0.003 | 0.189 | 0.189 |
|  | Freq. | 0.72 | 0.488 | 0.036 | 0.005 | 0.012 |
| ViT-B/32 | Probe | 0.87 | 0.495 | 0.026 | 0.002 | – |
|  | Stat. | 0.634 | 0.126 | 0.084 | 0.005 | 0.005 |
|  | Geom. | 0.19 | 0.089 | 0.012 | 0.005 | 0.005 |
|  | BiasDir | 1.22 | 0.527 | 0.066 | 0.005 | 0.005 |
|  | HBAC-NMI | 0.021 | 0.007 | 0.003 | 0.005 | 0.005 |
|  | Freq. | 0.78 | 0.486 | 0.037 | 0.005 | 0.005 |
| DINOv2 | Probe | 0.89 | 0.495 | 0.028 | 0.002 | – |
|  | Stat. | 0.581 | 0.184 | 0.118 | 0.005 | 0.006 |
|  | Geom. | 0.15 | 0.092 | 0.017 | 0.005 | 0.006 |
|  | BiasDir | 1.40 | 0.368 | 0.082 | 0.005 | 0.006 |
|  | HBAC-NMI | 0.011 | 0.009 | 0.003 | 0.224 | 0.224 |
|  | Freq. | 0.80 | 0.490 | 0.034 | 0.005 | 0.006 |
| RAD-DINO | Probe | 0.99 | 0.494 | 0.032 | 0.002 | – |
|  | Stat. | 0.635 | 0.251 | 0.055 | 0.005 | 0.005 |
|  | Geom. | 0.21 | 0.103 | 0.005 | 0.005 | 0.005 |
|  | BiasDir | 3.60 | 1.206 | 0.062 | 0.005 | 0.005 |
|  | HBAC-NMI | 0.053 | 0.023 | 0.005 | 0.005 | 0.005 |
|  | Freq. | 0.97 | 0.490 | 0.041 | 0.005 | 0.005 |
| MedSigLIP | Probe | 0.97 | 0.495 | 0.034 | 0.002 | – |
|  | Stat. | 0.792 | 0.436 | 0.127 | 0.005 | 0.006 |
|  | Geom. | 0.31 | 0.131 | 0.023 | 0.005 | 0.006 |
|  | BiasDir | 2.68 | 0.497 | 0.073 | 0.005 | 0.006 |
|  | HBAC-NMI | 0.021 | 0.015 | 0.005 | 0.109 | 0.109 |
|  | Freq. | 0.97 | 0.491 | 0.044 | 0.005 | 0.006 |

**Table F.12:** Raw statistics of the six detectors in every CheXpert cell, attribute age 60+. Columns as in Table F.11.

| Encoder | Detector | *s* | Null mean | Null SD | *p* | *q* |
|---|---|---|---|---|---|---|
| ResNet-50 | Probe | 0.75 | 0.497 | 0.023 | 0.002 | – |
|  | Stat. | 0.392 | 0.062 | 0.068 | 0.005 | 0.006 |
|  | Geom. | 0.19 | 0.091 | 0.018 | 0.005 | 0.006 |
|  | BiasDir | 0.84 | 0.425 | 0.070 | 0.005 | 0.006 |
|  | HBAC-NMI | 0.017 | 0.007 | 0.003 | 0.015 | 0.015 |
|  | Freq. | 0.74 | 0.491 | 0.032 | 0.005 | 0.006 |
| DenseNet-121 | Probe | 0.70 | 0.495 | 0.025 | 0.002 | – |
|  | Stat. | 0.627 | 0.205 | 0.121 | 0.005 | 0.005 |
|  | Geom. | 0.22 | 0.092 | 0.016 | 0.005 | 0.005 |
|  | BiasDir | 0.84 | 0.416 | 0.061 | 0.005 | 0.005 |
|  | HBAC-NMI | 0.039 | 0.013 | 0.004 | 0.005 | 0.005 |
|  | Freq. | 0.76 | 0.490 | 0.038 | 0.005 | 0.005 |
| ViT-B/16 | Probe | 0.70 | 0.497 | 0.026 | 0.002 | – |
|  | Stat. | 0.642 | 0.161 | 0.116 | 0.005 | 0.005 |
|  | Geom. | 0.18 | 0.090 | 0.015 | 0.005 | 0.005 |
|  | BiasDir | 0.73 | 0.441 | 0.067 | 0.005 | 0.005 |
|  | HBAC-NMI | 0.019 | 0.009 | 0.003 | 0.005 | 0.005 |
|  | Freq. | 0.69 | 0.489 | 0.039 | 0.005 | 0.005 |
| ViT-B/32 | Probe | 0.67 | 0.496 | 0.025 | 0.002 | – |
|  | Stat. | 0.452 | 0.126 | 0.087 | 0.005 | 0.006 |
|  | Geom. | 0.15 | 0.088 | 0.012 | 0.005 | 0.006 |
|  | BiasDir | 0.82 | 0.514 | 0.065 | 0.005 | 0.006 |
|  | HBAC-NMI | 0.013 | 0.008 | 0.003 | 0.050 | 0.050 |
|  | Freq. | 0.67 | 0.489 | 0.036 | 0.005 | 0.006 |
| DINOv2 | Probe | 0.75 | 0.496 | 0.027 | 0.002 | – |
|  | Stat. | 0.530 | 0.179 | 0.119 | 0.015 | 0.019 |
|  | Geom. | 0.15 | 0.090 | 0.018 | 0.015 | 0.019 |
|  | BiasDir | 0.85 | 0.359 | 0.081 | 0.005 | 0.012 |
|  | HBAC-NMI | 0.017 | 0.009 | 0.004 | 0.045 | 0.045 |
|  | Freq. | 0.71 | 0.487 | 0.038 | 0.005 | 0.012 |
| RAD-DINO | Probe | 0.83 | 0.494 | 0.031 | 0.002 | – |
|  | Stat. | 0.491 | 0.253 | 0.050 | 0.005 | 0.006 |
|  | Geom. | 0.16 | 0.102 | 0.005 | 0.005 | 0.006 |
|  | BiasDir | 1.97 | 1.189 | 0.064 | 0.005 | 0.006 |
|  | HBAC-NMI | 0.034 | 0.024 | 0.005 | 0.015 | 0.015 |
|  | Freq. | 0.77 | 0.489 | 0.043 | 0.005 | 0.006 |
| MedSigLIP | Probe | 0.80 | 0.497 | 0.032 | 0.002 | – |
|  | Stat. | 0.766 | 0.438 | 0.127 | 0.005 | 0.008 |
|  | Geom. | 0.22 | 0.129 | 0.024 | 0.010 | 0.012 |
|  | BiasDir | 0.87 | 0.501 | 0.074 | 0.005 | 0.008 |
|  | HBAC-NMI | 0.026 | 0.015 | 0.005 | 0.030 | 0.030 |
|  | Freq. | 0.77 | 0.493 | 0.043 | 0.005 | 0.008 |

**Table F.13:** Raw statistics of the six detectors in every CheXpert cell, attribute White versus non-White. Columns as in Table F.11.

| Encoder | Detector | *s* | Null mean | Null SD | *p* | *q* |
|---|---|---|---|---|---|---|
| ResNet-50 | Probe | 0.59 | 0.497 | 0.028 | 0.002 | – |
|  | Stat. | 0.282 | 0.065 | 0.068 | 0.015 | 0.037 |
|  | Geom. | 0.15 | 0.101 | 0.020 | 0.035 | 0.058 |
|  | BiasDir | 0.60 | 0.467 | 0.073 | 0.060 | 0.075 |
|  | HBAC-NMI | 0.012 | 0.009 | 0.003 | 0.149 | 0.149 |
|  | Freq. | 0.61 | 0.489 | 0.033 | 0.005 | 0.025 |
| DenseNet-121 | Probe | 0.60 | 0.497 | 0.027 | 0.002 | – |
|  | Stat. | 0.502 | 0.203 | 0.133 | 0.035 | 0.044 |
|  | Geom. | 0.19 | 0.102 | 0.020 | 0.005 | 0.012 |
|  | BiasDir | 0.65 | 0.459 | 0.062 | 0.010 | 0.017 |
|  | HBAC-NMI | 0.011 | 0.012 | 0.005 | 0.552 | 0.552 |
|  | Freq. | 0.60 | 0.485 | 0.041 | 0.005 | 0.012 |
| ViT-B/16 | Probe | 0.60 | 0.497 | 0.027 | 0.002 | – |
|  | Stat. | 0.467 | 0.160 | 0.126 | 0.020 | 0.033 |
|  | Geom. | 0.16 | 0.100 | 0.017 | 0.010 | 0.033 |
|  | BiasDir | 0.61 | 0.490 | 0.071 | 0.065 | 0.065 |
|  | HBAC-NMI | 0.018 | 0.012 | 0.004 | 0.065 | 0.065 |
|  | Freq. | 0.57 | 0.484 | 0.041 | 0.015 | 0.033 |
| ViT-B/32 | Probe | 0.60 | 0.497 | 0.026 | 0.002 | – |
|  | Stat. | 0.362 | 0.122 | 0.088 | 0.020 | 0.033 |
|  | Geom. | 0.14 | 0.096 | 0.013 | 0.010 | 0.025 |
|  | BiasDir | 0.69 | 0.568 | 0.067 | 0.060 | 0.075 |
|  | HBAC-NMI | 0.009 | 0.009 | 0.003 | 0.393 | 0.393 |
|  | Freq. | 0.60 | 0.485 | 0.042 | 0.010 | 0.025 |
| DINOv2 | Probe | 0.59 | 0.496 | 0.029 | 0.002 | – |
|  | Stat. | 0.333 | 0.175 | 0.118 | 0.114 | 0.155 |
|  | Geom. | 0.13 | 0.098 | 0.019 | 0.085 | 0.155 |
|  | BiasDir | 0.43 | 0.393 | 0.091 | 0.289 | 0.289 |
|  | HBAC-NMI | 0.018 | 0.014 | 0.004 | 0.124 | 0.155 |
|  | Freq. | 0.61 | 0.483 | 0.042 | 0.005 | 0.025 |
| RAD-DINO | Probe | 0.61 | 0.499 | 0.037 | 0.004 | – |
|  | Stat. | 0.342 | 0.251 | 0.065 | 0.104 | 0.131 |
|  | Geom. | 0.13 | 0.112 | 0.007 | 0.015 | 0.037 |
|  | BiasDir | 1.55 | 1.299 | 0.059 | 0.005 | 0.025 |
|  | HBAC-NMI | 0.028 | 0.028 | 0.006 | 0.532 | 0.532 |
|  | Freq. | 0.58 | 0.485 | 0.050 | 0.050 | 0.083 |
| MedSigLIP | Probe | 0.57 | 0.496 | 0.036 | 0.022 | – |
|  | Stat. | 0.495 | 0.432 | 0.126 | 0.338 | 0.423 |
|  | Geom. | 0.18 | 0.141 | 0.025 | 0.090 | 0.224 |
|  | BiasDir | 0.54 | 0.540 | 0.075 | 0.433 | 0.433 |
|  | HBAC-NMI | 0.017 | 0.016 | 0.005 | 0.323 | 0.423 |
|  | Freq. | 0.58 | 0.489 | 0.048 | 0.045 | 0.224 |

**Table F.14:**
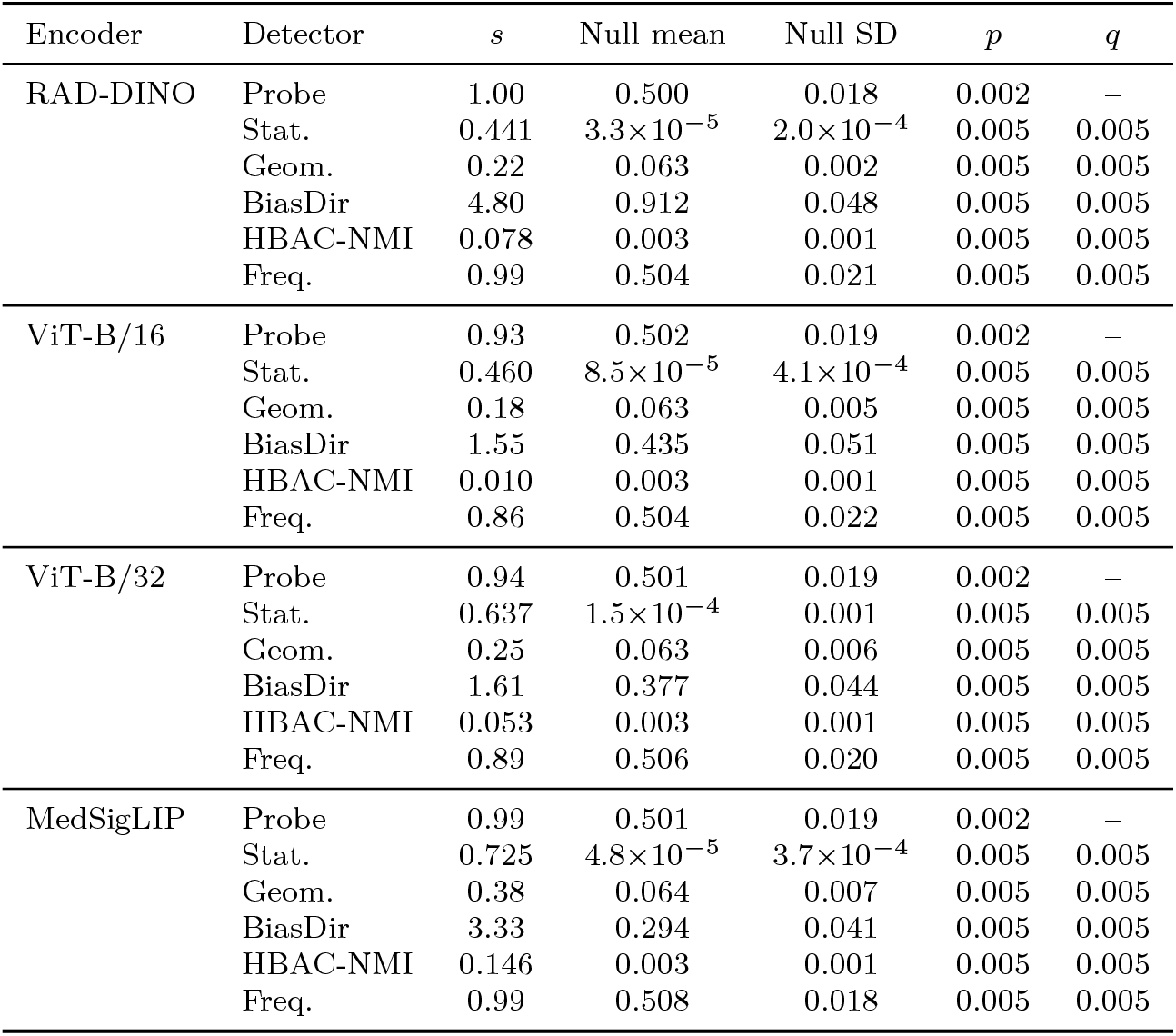
Raw statistics of the six detectors in every MIMIC-CXR cell, attribute sex (task in-hospital mortality, primary audit, seed 42). ViT-B/32 uses the file rebuilt from the source (Table F.5). Columns as in Table F.11.

| Encoder | Detector | *s* | Null mean | Null SD | *p* | *q* |
|---|---|---|---|---|---|---|
| RAD-DINO | Probe | 1.00 | 0.500 | 0.018 | 0.002 | – |
|  | Stat. | 0.441 | 3.3×10<sup>−5</sup> | 2.0×10<sup>−4</sup> | 0.005 | 0.005 |
|  | Geom. | 0.22 | 0.063 | 0.002 | 0.005 | 0.005 |
|  | BiasDir | 4.80 | 0.912 | 0.048 | 0.005 | 0.005 |
|  | HBAC-NMI | 0.078 | 0.003 | 0.001 | 0.005 | 0.005 |
|  | Freq. | 0.99 | 0.504 | 0.021 | 0.005 | 0.005 |
| ViT-B/16 | Probe | 0.93 | 0.502 | 0.019 | 0.002 | – |
|  | Stat. | 0.460 | 8.5×10<sup>−5</sup> | 4.1×10<sup>−4</sup> | 0.005 | 0.005 |
|  | Geom. | 0.18 | 0.063 | 0.005 | 0.005 | 0.005 |
|  | BiasDir | 1.55 | 0.435 | 0.051 | 0.005 | 0.005 |
|  | HBAC-NMI | 0.010 | 0.003 | 0.001 | 0.005 | 0.005 |
|  | Freq. | 0.86 | 0.504 | 0.022 | 0.005 | 0.005 |
| ViT-B/32 | Probe | 0.94 | 0.501 | 0.019 | 0.002 | – |
|  | Stat. | 0.637 | 1.5×10<sup>−4</sup> | 0.001 | 0.005 | 0.005 |
|  | Geom. | 0.25 | 0.063 | 0.006 | 0.005 | 0.005 |
|  | BiasDir | 1.61 | 0.377 | 0.044 | 0.005 | 0.005 |
|  | HBAC-NMI | 0.053 | 0.003 | 0.001 | 0.005 | 0.005 |
|  | Freq. | 0.89 | 0.506 | 0.020 | 0.005 | 0.005 |
| MedSigLIP | Probe | 0.99 | 0.501 | 0.019 | 0.002 | – |
|  | Stat. | 0.725 | 4.8×10<sup>−5</sup> | 3.7×10<sup>−4</sup> | 0.005 | 0.005 |
|  | Geom. | 0.38 | 0.064 | 0.007 | 0.005 | 0.005 |
|  | BiasDir | 3.33 | 0.294 | 0.041 | 0.005 | 0.005 |
|  | HBAC-NMI | 0.146 | 0.003 | 0.001 | 0.005 | 0.005 |
|  | Freq. | 0.99 | 0.508 | 0.018 | 0.005 | 0.005 |

**Table F.15:** Raw statistics of the six detectors in every MIMIC-CXR cell, attribute age 60+. Columns as in Table F.11.

| Encoder | Detector | *s* | Null mean | Null SD | *p* | *q* |
|---|---|---|---|---|---|---|
| RAD-DINO | Probe | 0.89 | 0.508 | 0.021 | 0.002 | – |
|  | Stat. | 0.312 | 6.5×10<sup>−5</sup> | 3.1×10<sup>−4</sup> | 0.005 | 0.005 |
|  | Geom. | 0.18 | 0.067 | 0.003 | 0.005 | 0.005 |
|  | BiasDir | 2.17 | 0.971 | 0.050 | 0.005 | 0.005 |
|  | HBAC-NMI | 0.073 | 0.003 | 0.002 | 0.005 | 0.005 |
|  | Freq. | 0.88 | 0.522 | 0.020 | 0.005 | 0.005 |
| ViT-B/16 | Probe | 0.80 | 0.510 | 0.020 | 0.002 | – |
|  | Stat. | 0.587 | 6.5×10<sup>−5</sup> | 4.3×10<sup>−4</sup> | 0.005 | 0.005 |
|  | Geom. | 0.23 | 0.067 | 0.006 | 0.005 | 0.005 |
|  | BiasDir | 1.12 | 0.467 | 0.056 | 0.005 | 0.005 |
|  | HBAC-NMI | 0.024 | 0.003 | 0.001 | 0.005 | 0.005 |
|  | Freq. | 0.82 | 0.525 | 0.020 | 0.005 | 0.005 |
| ViT-B/32 | Probe | 0.83 | 0.511 | 0.019 | 0.002 | – |
|  | Stat. | 0.654 | 1.0×10<sup>−4</sup> | 0.001 | 0.005 | 0.005 |
|  | Geom. | 0.28 | 0.067 | 0.007 | 0.005 | 0.005 |
|  | BiasDir | 1.26 | 0.404 | 0.046 | 0.005 | 0.005 |
|  | HBAC-NMI | 0.025 | 0.003 | 0.002 | 0.005 | 0.005 |
|  | Freq. | 0.83 | 0.524 | 0.019 | 0.005 | 0.005 |
| MedSigLIP | Probe | 0.89 | 0.512 | 0.020 | 0.002 | – |
|  | Stat. | 0.779 | 1.8×10<sup>−4</sup> | 0.001 | 0.005 | 0.005 |
|  | Geom. | 0.39 | 0.066 | 0.009 | 0.005 | 0.005 |
|  | BiasDir | 1.29 | 0.315 | 0.044 | 0.005 | 0.005 |
|  | HBAC-NMI | 0.068 | 0.003 | 0.002 | 0.005 | 0.005 |
|  | Freq. | 0.89 | 0.524 | 0.019 | 0.005 | 0.005 |

**Table F.16:**
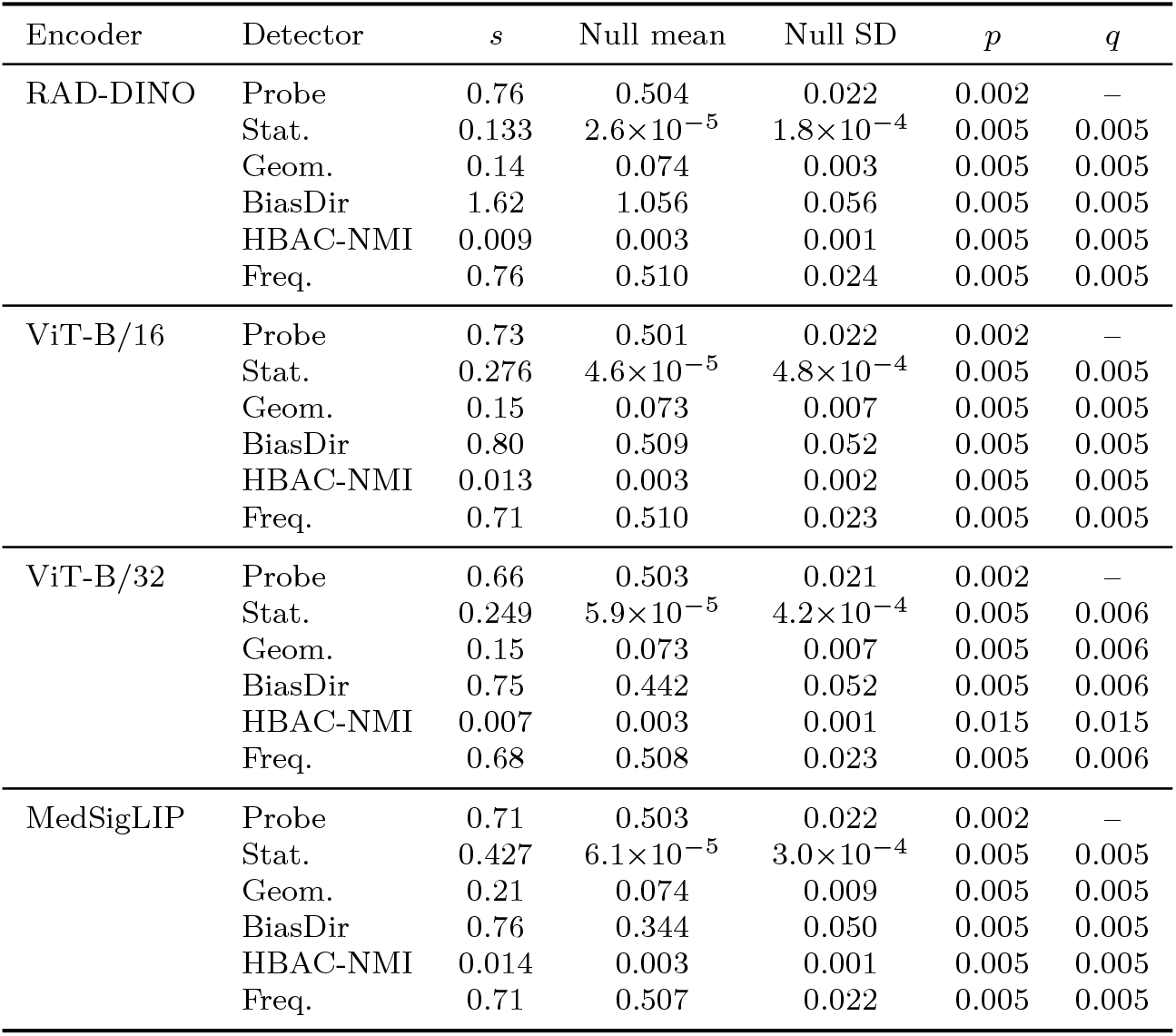
Raw statistics of the six detectors in every MIMIC-CXR cell, attribute White versus non-White. Columns as in Table F.11.

| Encoder | Detector | *s* | Null mean | Null SD | *p* | *q* |
|---|---|---|---|---|---|---|
| RAD-DINO | Probe | 0.76 | 0.504 | 0.022 | 0.002 | – |
|  | Stat. | 0.133 | 2.6×10<sup>−5</sup> | 1.8×10<sup>−4</sup> | 0.005 | 0.005 |
|  | Geom. | 0.14 | 0.074 | 0.003 | 0.005 | 0.005 |
|  | BiasDir | 1.62 | 1.056 | 0.056 | 0.005 | 0.005 |
|  | HBAC-NMI | 0.009 | 0.003 | 0.001 | 0.005 | 0.005 |
|  | Freq. | 0.76 | 0.510 | 0.024 | 0.005 | 0.005 |
| ViT-B/16 | Probe | 0.73 | 0.501 | 0.022 | 0.002 | – |
|  | Stat. | 0.276 | 4.6×10<sup>−5</sup> | 4.8×10<sup>−4</sup> | 0.005 | 0.005 |
|  | Geom. | 0.15 | 0.073 | 0.007 | 0.005 | 0.005 |
|  | BiasDir | 0.80 | 0.509 | 0.052 | 0.005 | 0.005 |
|  | HBAC-NMI | 0.013 | 0.003 | 0.002 | 0.005 | 0.005 |
|  | Freq. | 0.71 | 0.510 | 0.023 | 0.005 | 0.005 |
| ViT-B/32 | Probe | 0.66 | 0.503 | 0.021 | 0.002 | – |
|  | Stat. | 0.249 | 5.9×10<sup>−5</sup> | 4.2×10<sup>−4</sup> | 0.005 | 0.006 |
|  | Geom. | 0.15 | 0.073 | 0.007 | 0.005 | 0.006 |
|  | BiasDir | 0.75 | 0.442 | 0.052 | 0.005 | 0.006 |
|  | HBAC-NMI | 0.007 | 0.003 | 0.001 | 0.015 | 0.015 |
|  | Freq. | 0.68 | 0.508 | 0.023 | 0.005 | 0.006 |
| MedSigLIP | Probe | 0.71 | 0.503 | 0.022 | 0.002 | – |
|  | Stat. | 0.427 | 6.1×10<sup>−5</sup> | 3.0×10<sup>−4</sup> | 0.005 | 0.005 |
|  | Geom. | 0.21 | 0.074 | 0.009 | 0.005 | 0.005 |
|  | BiasDir | 0.76 | 0.344 | 0.050 | 0.005 | 0.005 |
|  | HBAC-NMI | 0.014 | 0.003 | 0.001 | 0.005 | 0.005 |
|  | Freq. | 0.71 | 0.507 | 0.022 | 0.005 | 0.005 |

## Appendix G. Embedding visualisations

Figure G.1 shows a one-dimensional linear discriminant analysis (LDA) projection for sex on MIMIC-CXR RAD-DINO embeddings, fitted on training folds and shown for held-out images. An LDA fitted and plotted on the same images would separate the groups in 768 dimensions even without signal. The held-out AUROC of the projection was 0.998. Figures G.2 and G.3 show UMAP projections. These figures are illustrations, and the tests are in the tables.

**Figure G.1.**
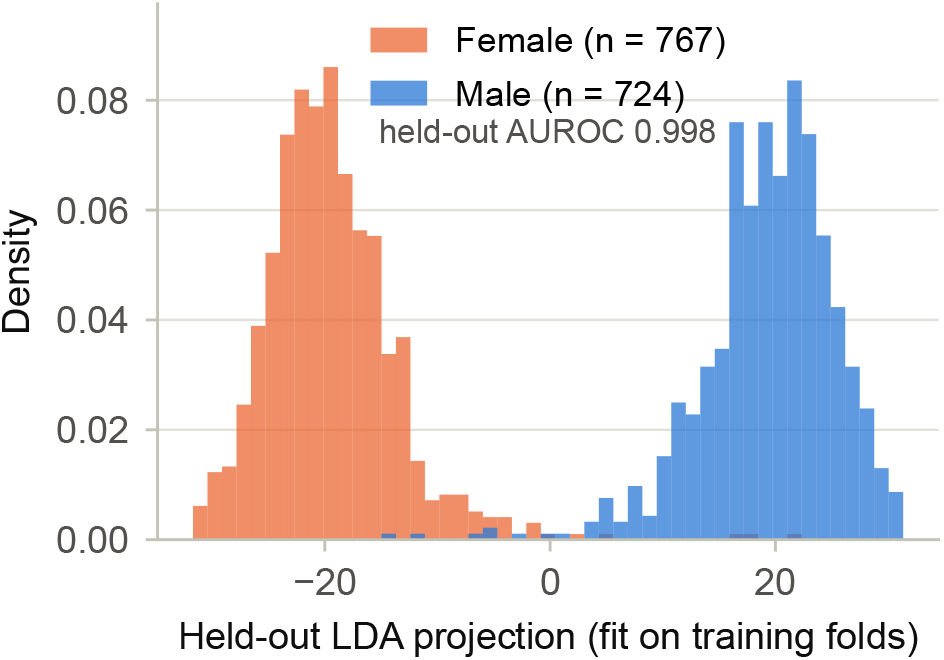
Linear discriminant analysis (LDA) projection for sex, MIMIC-CXR RAD-DINO (768-dim), fitted on training folds, held-out images shown (1,491 in total over folds).

**Figure G.2.**
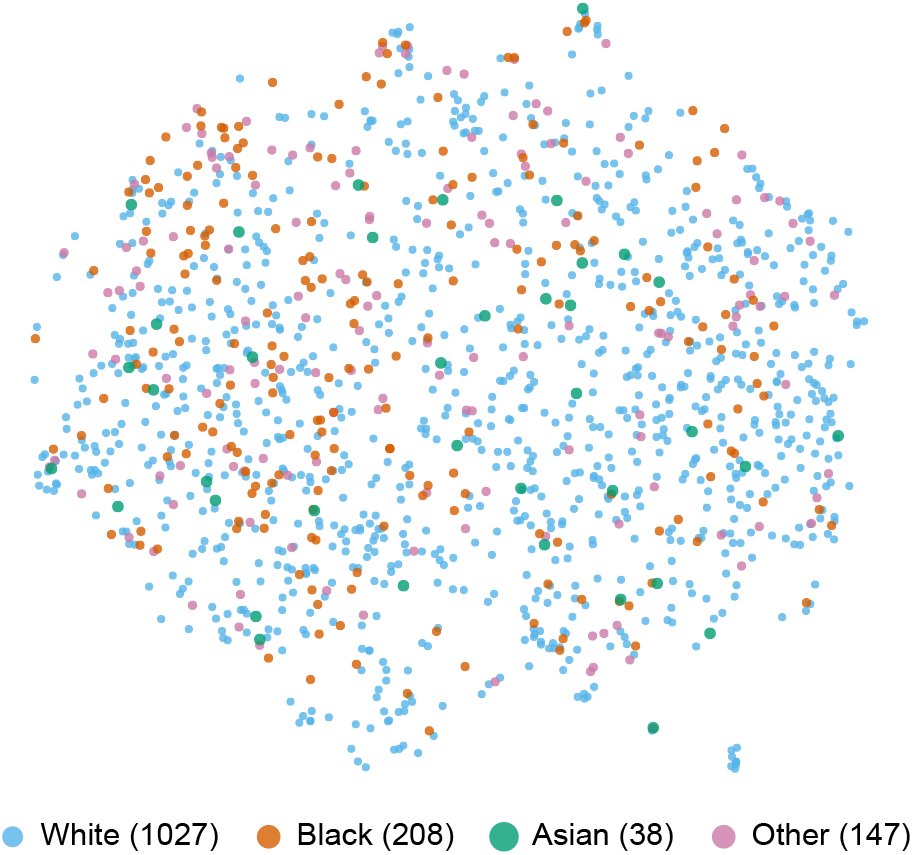
UMAP projection of MIMIC-CXR RAD-DINO embeddings (768-dim, standardised) coloured by race, 1,420 subjects. The 71 subjects without a recorded race are left out, as in the audit.

**Figure G.3.**
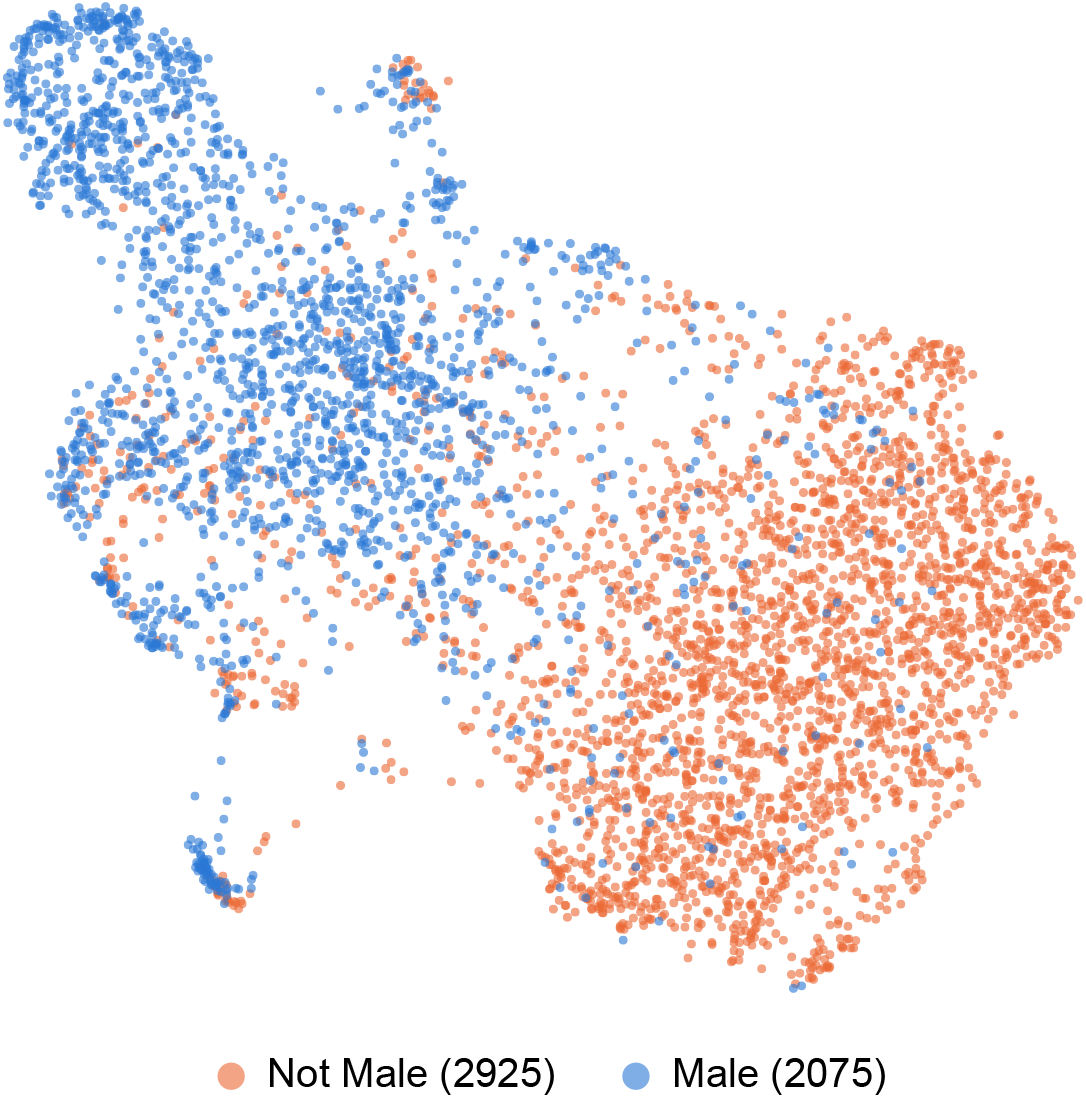
UMAP projection of CelebA ResNet-50 embeddings (2,048-dim, standardised, random 5,000-image subsample) coloured by the Male attribute.

## References

[1] A. Esteva, B. Kuprel, R. A. Novoa, J. Ko, S. M. Swetter, H. M. Blau, S. Thrun, Dermatologist-level classification of skin cancer with deep neural networks, Nature 542 (7639) (2017) 115–118. doi:10.1038/nature21056.

[2] P. Rajpurkar, J. Irvin, K. Zhu, B. Yang, H. Mehta, T. Duan, D. Ding, A. Bagul, C. Langlotz, K. Shpanskaya, M. P. Lungren, A. Y. Ng, CheXNet: Radiologist-level pneumonia detection on chest X-rays with deep learning, arXiv preprint arXiv:1711.05225 (2017).

[3] S. M. McKinney, M. Sieniek, V. Godbole, J. Godber, R. Patel, R. Padilla, D. Shanmugam, D. Phung, Q. V. Le, et al., International evaluation of an AI system for breast cancer screening, Nature 577 (7788) (2020) 89–94. doi:10.1038/s41586-019-1799-6.

[4] R. Geirhos, J.-H. Jacobsen, C. Michaelis, R. Zemel, W. Brendel, M. Bethge, F. A. Wichmann, Shortcut learning in deep neural networks, Nature Machine Intelligence 2 (11) (2020) 665–673. doi:10.1038/s42256-020-00257-z.

[5] L. Oakden-Rayner, J. Dunnmon, G. Carneiro, C. Ré, Hidden stratification causes clinically meaningful failures in machine learning for medical imaging, in: Proceedings of the ACM Conference on Health, Inference, and Learning (CHIL), 2020, pp. 151–159. doi:10.1145/3368555.3384468.

[6] J. R. Zech, M. A. Badgeley, M. Liu, A. B. Costa, J. J. Titano, E. K. Oermann, Variable generalization performance of a deep learning model to detect pneumonia in chest radiographs: A cross-sectional study, PLoS Medicine 15 (11) (2018) e1002683. doi:10.1371/journal.pmed.1002683.

[7] M. A. Badgeley, J. R. Zech, L. Oakden-Rayner, B. S. Glicksberg, M. Liu, W. Gale, M. V. McConnell, B. Percha, T. M. Snyder, J. T. Dudley, Deep learning predicts hip fracture using confounding patient and healthcare variables, npj Digital Medicine 2 (2019) 31. doi: 10.1038/s41746-019-0105-1.

[8] J. W. Gichoya, I. Banerjee, A. R. Bhimireddy, J. L. Burns, L. A. Celi, L.-C. Chen, R. Correa, N. Davila-Gavilan, J. M. Dueñas-Dominguez, et al., AI recognition of patient race in medical imaging: a modelling study, The Lancet Digital Health 4 (6) (2022) e406–e414. doi:10.1016/S2589-7500(22)00063-2.

[9] K. Čevora, B. Glocker, W. Bai, Quantifying the impact of population shift across age and sex for abdominal organ segmentation, in: Ethics and Fairness in Medical Imaging (FAIMI/EPIMI 2024, MICCAI Work-shops), Vol. 15198 of Lecture Notes in Computer Science, Springer, 2024, pp. 88–97. doi:10.1007/978-3-031-72787-0_9.

[10] A. Brown, N. Tomasev, J. Freyberg, Y. Liu, A. Karthikesalingam, J. Schrouff, Detecting shortcut learning for fair medical AI using short-cut testing, Nature Communications 14 (2023) 4314. doi:10.1038/s41467-023-39902-7.

[11] B. Glocker, C. Jones, M. Bernhardt, S. Winzeck, Algorithmic encoding of protected characteristics in chest X-ray disease detection models, eBioMedicine 89 (2023) 104467. doi:10.1016/j.ebiom.2023.104467.

[12] B. G. Hill, F. L. Koback, P. L. Schilling, The risk of shortcutting in deep learning algorithms for medical imaging research, Scientific Reports 14 (1) (2024) 29224. doi:10.1038/s41598-024-79838-6.

[13] E. Petersen, E. Ferrante, M. Ganz, A. Feragen, Are demographically invariant models and representations in medical imaging fair?, arXiv preprint, v3 dated 3 Jul 2024. No journal version found (2023). arXiv: 2305.01397, doi:10.48550/arXiv.2305.01397.

[14] M. A. Kina, E. Petersen, Prevalence calibration as shortcut mitigation, accepted at MLMI 2026 (MICCAI workshop) (2026). arXiv: 2609.07922, doi:10.48550/arXiv.2609.07922.

[15] S. Bird, M. Dudík, R. Edgar, B. Horn, R. Lutz, V. Milan, M. Sameki, H. Wallach, K. Walker, Fairlearn: A toolkit for assessing and improving fairness in AI, in: Microsoft Research, 2020.

[16] R. K. E. Bellamy, K. Dey, M. Hind, S. C. Hoffman, S. Houde, K. Kannan, P. Lohia, J. Martino, S. Mehta, et al., AI Fairness 360: An extensible toolkit for detecting and mitigating algorithmic bias, IBM Journal of Research and Development 63 (4/5) (2019) 4:1–4:15. doi:10.1147/JRD.2019.2942287.

[17] G. Alain, Y. Bengio, Understanding intermediate layers using linear classifier probes, arXiv preprint arXiv:1610.01644 (2016).

[18] Y. Belinkov, Probing classifiers: Promises, shortcomings, and advances, Computational Linguistics 48 (1) (2022) 207–219. doi:10.1162/coli_a_00422.

[19] L. Fay, E. Cobos, B. Yang, S. Gatidis, T. Küstner, Avoiding shortcut-learning by mutual information minimization in deep learning-based image processing, IEEE Access 11 (2023) 64070–64086. doi:10.1109/ACCESS.2023.3289397.

[20] S. Müller, L. Fay, L. M. Koch, S. Gatidis, T. Küstner, P. Berens, Benchmarking dependence measures to prevent shortcut learning in medical imaging, in: Machine Learning in Medical Imaging (MLMI 2024), Lecture Notes in Computer Science, Springer Nature Switzerland, Cham, 2024, pp. 53–62, arXiv:2407.18792. doi:10.1007/978-3-031-73290-4_6.

[21] R. Geirhos, P. Rubisch, C. Michaelis, M. Bethge, F. A. Wichmann, W. Brendel, ImageNet-trained CNNs are biased towards texture; in-creasing shape bias improves accuracy and robustness, in: Proceedings of the International Conference on Learning Representations (ICLR), 2019.

[22] R. T. McCoy, E. Pavlick, T. Linzen, Right for the wrong reasons: Diagnosing syntactic heuristics in natural language inference, in: Proceedings of the 57th Annual Meeting of the Association for Computational Linguistics (ACL), 2019, pp. 3428–3448. doi:10.18653/v1/P19-1334.

[23] D. Steinmann, et al., Navigating shortcuts, spurious correlations, and confounders: From origins via detection to mitigation, arXiv preprint arXiv:2412.05152 (2024).

[24] W. Ye, et al., The Clever Hans mirage: A comprehensive survey on spurious correlations in machine learning, arXiv preprint arXiv:2402.12715 (2024).

[25] M. Schmitt, R. C. Maron, A. Hekler, A. Stenzinger, A. Hauschild, M. Weichenthal, M. Tiemann, D. Krahl, H. Kutzner, J. S. Utikal, S. Haferkamp, J. N. Kather, F. Klauschen, E. Krieghoff-Henning, S. Fröhling, C. von Kalle, T. J. Brinker, Hidden variables in deep learning digital pathology and their potential to cause batch effects: Prediction model study, Journal of Medical Internet Research 23 (2) (2021) e23436. doi:10.2196/23436.

[26] A. Jiménez-Sánchez, D. Juodelyte, B. Chamberlain, V. Cheplygina, Detecting shortcuts in medical images – a case study in chest X-rays, in: Proceedings of the IEEE International Symposium on Biomedical Imaging (ISBI), 2023, pp. 1–5. doi:10.1109/ISBI53787.2023.10230572.

[27] A. J. DeGrave, J. D. Janizek, S.-I. Lee, AI for radiographic COVID-19 detection selects shortcuts over signal, Nature Machine Intelligence 3 (7) (2021) 610–619. doi:10.1038/s42256-021-00338-7.

[28] A. Bissoto, E. Valle, S. Avila, Debiasing skin lesion datasets and models? not so fast, in: Proceedings of the IEEE/CVF Conference on Computer Vision and Pattern Recognition Workshops (CVPRW), 2020, pp. 3192–3201. doi:10.1109/CVPRW50498.2020.00378.

[29] I. Banerjee, K. Bhattacharjee, J. L. Burns, H. Trivedi, S. Purkayastha, L. Seyyed-Kalantari, B. N. Patel, R. Shiradkar, J. Gichoya, “shortcuts” causing bias in radiology artificial intelligence: Causes, evaluation, and mitigation, Journal of the American College of Radiology 20 (9) (2023) 842–851. doi:10.1016/j.jacr.2023.06.025.

[30] C. Ong Ly, B. Unnikrishnan, T. Tadic, T. Patel, J. Duhamel, S. Kandel, Y. Moayedi, M. Brudno, A. Hope, H. Ross, C. McIntosh, Shortcut learning in medical AI hinders generalization: method for estimating AI model generalization without external data, npj Digital Medicine 7 (2024) 124. doi:10.1038/s41746-024-01118-4.

[31] B. Glocker, C. Jones, M. Roschewitz, S. Winzeck, Risk of bias in chest radiography deep learning foundation models, Radiology: Artificial Intelligence 5 (6) (2023) e230060. doi:10.1148/ryai.230060.

[32] C. Jones, M. Roschewitz, B. Glocker, The role of subgroup separability in group-fair medical image classification, in: Medical Image Computing and Computer Assisted Intervention (MICCAI 2023), Lecture Notes in Computer Science, Springer Nature Switzerland, Cham, 2023, pp. 179–188. doi:10.1007/978-3-031-43898-1_18.

[33] C. Jones, D. C. Castro, F. De Sousa Ribeiro, O. Oktay, M. McCradden, B. Glocker, A causal perspective on dataset bias in machine learning for medical imaging, Nature Machine Intelligence 6 (2) (2024) 138–146. doi:10.1038/s42256-024-00797-8.

[34] Y. Yang, H. Zhang, J. W. Gichoya, D. Katabi, M. Ghassemi, The limits of fair medical imaging AI in real-world generalization, Nature Medicine 30 (10) (2024) 2838–2848. doi:10.1038/s41591-024-03113-4.

[35] T. Bolukbasi, K.-W. Chang, J. Zou, V. Saligrama, A. Kalai, Man is to computer programmer as woman is to homemaker? Debiasing word embeddings, in: Advances in Neural Information Processing Systems (NeurIPS), 2016, pp. 4349–4357, arXiv:1607.06520.

[36] H. Gonen, Y. Goldberg, Lipstick on a pig: Debiasing methods cover up systematic gender biases in word embeddings but do not remove them, in: Proceedings of the 2019 Conference of the North American Chapter of the Association for Computational Linguistics: Human Language Technologies (NAACL-HLT), 2019, pp. 609–614.

[37] S. Ravfogel, Y. Elazar, H. Gonen, M. Twiton, Y. Goldberg, Null it out: Guarding protected attributes by iterative nullspace projection, in: Proceedings of the 58th Annual Meeting of the Association for Computational Linguistics (ACL), 2020, pp. 7237–7256. doi:10.18653/v1/2020.acl-main.647.

[38] R. Steed, A. Caliskan, Image representations learned with unsupervised pre-training contain human-like biases, in: Proceedings of the ACM Conference on Fairness, Accountability, and Transparency (FAccT), 2021, pp. 701–713.

[39] T. B. Berrett, Y. Wang, R. F. Barber, R. J. Samworth, The conditional permutation test for independence while controlling for confounders, Journal of the Royal Statistical Society Series B: Statistical Methodology 82 (1) (2020) 175–197. doi:10.1111/rssb.12340.

[40] A. Gretton, K. M. Borgwardt, M. J. Rasch, B. Schölkopf, A. Smola, A kernel two-sample test, Journal of Machine Learning Research 13 (25) (2012) 723–773.

[41] C. DiCiccio, S. Vasudevan, K. Basu, J. Frankle, K. Batmanghelich, Evaluating fairness using permutation tests, in: Proceedings of the 26th ACM SIGKDD International Conference on Knowledge Discovery & Data Mining (KDD), 2020. doi:10.1145/3394486.3403199.

[42] N. S. Sohoni, J. A. Dunnmon, G. Angus, A. Gu, C. Ré, No sub-class left behind: Fine-grained robustness in coarse-grained classification problems, in: Advances in Neural Information Processing Systems (NeurIPS), 2020, pp. 19339–19352.

[43] S. Eyuboglu, M. Varma, K. Saab, J.-B. Delbrouck, C. Lee-Messer, J. Dunnmon, J. Zou, C. Ré, Domino: Discovering systematic errors with cross-modal embeddings, in: Proceedings of the International Conference on Learning Representations (ICLR), 2022.

[44] A. Krishnakumar, V. Prabhu, S. Sudhakar, J. Hoffman, UDIS: Unsupervised discovery of bias in deep visual recognition models, in: Proceedings of the British Machine Vision Conference (BMVC), 2021, arXiv:2110.15499. doi:10.5244/C.35.102.

[45] P. Saleiro, B. Kuester, L. Hinkson, J. London, A. Stevens, A. Anisfeld, K. T. Rodolfa, R. Ghani, Aequitas: A bias and fairness audit toolkit, arXiv preprint arXiv:1811.05577 (2018).

[46] Y. Zong, Y. Yang, T. Hospedales, MEDFAIR: Benchmarking fairness for medical imaging, in: Proceedings of the International Conference on Learning Representations (ICLR), 2023, arXiv:2210.01725.

[47] R. Jin, et al., FairMedFM: Fairness benchmarking for medical imaging foundation models, in: Advances in Neural Information Processing Systems (NeurIPS) Datasets and Benchmarks, 2024.

[48] J. Wexler, M. Pushkarna, T. Bolukbasi, M. Wattenberg, F. Viégas, J. Wilson, The What-If Tool: Interactive probing of machine learning models, IEEE Transactions on Visualization and Computer Graphics 26 (1) (2020) 56–65. doi:10.1109/TVCG.2019.2934619.

[49] A. Caliskan, J. J. Bryson, A. Narayanan, Semantics derived automatically from language corpora contain human-like biases, Science 356 (6334) (2017) 183–186. doi:10.1126/science.aal4230.

[50] B. Phipson, G. K. Smyth, Permutation P-values should never be zero: calculating exact P-values when permutations are randomly drawn, Statistical Applications in Genetics and Molecular Biology 9 (1) (2010) Article 39. doi:10.2202/1544-6115.1585.

[51] Y. Benjamini, Y. Hochberg, Controlling the false discovery rate: A practical and powerful approach to multiple testing, Journal of the Royal Statistical Society: Series B (Methodological) 57 (1) (1995) 289–300. doi:10.1111/j.2517-6161.1995.tb02031.x.

[52] J. Misztal-Radecka, B. Indurkhya, Bias-aware hierarchical clustering for detecting the discriminated groups of users in recommendation systems, Information Processing & Management 58 (3) (2021) 102519. doi:10.1016/j.ipm.2021.102519.

[53] S. Wang, R. Veldhuis, C. Brune, N. Strisciuglio, What do neural networks learn in image classification? A frequency shortcut perspective, in: Proceedings of the IEEE/CVF International Conference on Computer Vision (ICCV), 2023, pp. 1433–1442. doi:10.1109/ICCV51070.2023.00138.

[54] P. Kirichenko, P. Izmailov, A. G. Wilson, Last layer re-training is sufficient for robustness to spurious correlations, in: Proceedings of the International Conference on Learning Representations (ICLR), 2023.

[55] D. R. Nyholt, A simple correction for multiple testing for single-nucleotide polymorphisms in linkage disequilibrium with each other, The American Journal of Human Genetics 74 (4) (2004) 765–769. doi:10.1086/383251.

[56] J. Li, L. Ji, Adjusting multiple testing in multilocus analyses using the eigenvalues of a correlation matrix, Heredity 95 (3) (2005) 221–227. doi:10.1038/sj.hdy.6800717.

[57] J. Irvin, P. Rajpurkar, M. Ko, Y. Yu, S. Ciurea-Ilcus, C. Chute, H. Marklund, B. Haghgoo, R. Ball, K. Shpanskaya, et al., CheXpert: A large chest radiograph dataset with uncertainty labels and expert comparison, in: Proceedings of the AAAI Conference on Artificial Intelligence, Vol. 33, 2019, pp. 590–597. doi:10.1609/aaai.v33i01.3301590.

[58] A. E. W. Johnson, T. J. Pollard, S. J. Berkowitz, N. R. Greenbaum, M. P. Lungren, C.-y. Deng, R. G. Mark, S. Horng, MIMIC-CXR, a de-identified publicly available database of chest radiographs with free-text reports, Scientific Data 6 (2019) 317. doi:10.1038/s41597-019-0322-0.

[59] S. Cajas Ordóñez, F. Ocampo Osorio, D. E. Koh, R. Al Attrach, A. Marzullo, A. Guerra-Adames, J. A. Andrade, S. T. Goh, C.-Y. Chen, R. Gorijavolu, X. Yang, N. D. Hebdon, L. A. Celi, Quantum kernel advantage over classical collapse in medical foundation model embeddings, arXiv:2604.24597 (2026). arXiv:2604.24597, doi:10.48550/arXiv.2604.24597.

[60] A. E. W. Johnson, L. Bulgarelli, L. Shen, A. Gayles, A. Shammout, S. Horng, T. J. Pollard, S. Hao, B. Moody, B. Gow, L.-w. H. Lehman, L. A. Celi, R. G. Mark, MIMIC-IV, a freely accessible electronic health record dataset, Scientific Data 10 (2023) 1. doi:10.1038/s41597-022-01899-x.

[61] F. Pérez-García, H. Sharma, S. Bond-Taylor, K. Bouzid, V. Salvatelli, M. Ilse, S. Bannur, D. C. Castro, A. Schwaighofer, M. P. Lungren, M. T. Wetscherek, N. Codella, S. L. Hyland, J. Alvarez-Valle, O. Oktay, Exploring scalable medical image encoders beyond text supervision, Nature Machine Intelligence 7 (1) (2025) 119–130. doi:10.1038/s42256-024-00965-w.

[62] Z. Liu, P. Luo, X. Wang, X. Tang, Deep learning face attributes in the wild, in: Proceedings of the IEEE International Conference on Computer Vision (ICCV), 2015, pp. 3730–3738.

[63] T. G. Dietterich, Ensemble methods in machine learning, in: Multiple Classifier Systems (MCS 2000), Vol. 1857 of Lecture Notes in Computer Science, Springer, 2000, pp. 1–15. doi:10.1007/3-540-45014-9_1.

[64] R. A. Jacobs, M. I. Jordan, S. J. Nowlan, G. E. Hinton, Adaptive mixtures of local experts, Neural Computation 3 (1) (1991) 79–87. doi:10.1162/neco.1991.3.1.79.

[65] A. Q. Xiang, T. Tohyama, A. C. Bank, Q. Bui, R. Gorijavolu, J. A. Garcia Henao, N. Jaiswal, A. Kelshiker, K. Madapati, S. Cajas Or-dóñez, M. Patel, N. Prakash, L. A. Celi, Distributed open justice oversight (DOJO): A community-driven, modality-agnostic platform for adversarial evaluation of health AI, sSRN preprint 6676818 (2026). URL https://papers.ssrn.com/sol3/papers.cfm?abstract_id=6676818

[66] S. Sagawa, P. W. Koh, T. B. Hashimoto, P. Liang, Distributionally robust neural networks for group shifts: On the importance of regularization for worst-case generalization, in: Proceedings of the International Conference on Learning Representations (ICLR), 2020.

[67] J. Nam, H. Cha, S. Ahn, J. Lee, J. Shin, Learning from failure: De-biasing classifier from biased classifier, in: Advances in Neural Information Processing Systems (NeurIPS), Vol. 33, 2020, pp. 20673–20684.

[68] S. Lapuschkin, S. Wäldchen, A. Binder, G. Montavon, W. Samek, K.-R. Müller, Unmasking Clever Hans predictors and assessing what machines really learn, Nature Communications 10 (2019) 1096. doi:10.1038/s41467-019-08987-4.

[69] R. R. Selvaraju, M. Cogswell, A. Das, R. Vedantam, D. Parikh, D. Batra, Grad-CAM: Visual explanations from deep networks via gradient-based localization, in: Proceedings of the IEEE International Conference on Computer Vision (ICCV), 2017, pp. 618–626. doi:10.1109/ICCV.2017.74.

[70] M. Arjovsky, L. Bottou, I. Gulrajani, D. Lopez-Paz, Invariant risk minimization, arXiv preprint arXiv:1907.02893 (2019).

[71] D. C. Castro, I. Walker, B. Glocker, Causality matters in medical imaging, Nature Communications 11 (2020) 3673. doi:10.1038/s41467-020-17478-w.

[72] Y. Elazar, Y. Goldberg, Adversarial removal of demographic attributes from text data, in: Proceedings of the 2018 Conference on Empirical Methods in Natural Language Processing (EMNLP), 2018, pp. 11–21. doi:10.18653/v1/D18-1002.

[73] D. Madras, E. Creager, T. Pitassi, R. Zemel, Learning adversarially fair and transferable representations, in: Proceedings of the International Conference on Machine Learning (ICML), Vol. 80 of PMLR, 2018, pp. 3384–3393.

[74] R. Zemel, Y. Wu, K. Swersky, T. Pitassi, C. Dwork, Learning fair representations, in: Proceedings of the International Conference on Machine Learning (ICML), Vol. 28 of PMLR, 2013, pp. 325–333.

